# Eye-brain connections revealed by multimodal retinal and brain imaging genetics in the UK Biobank

**DOI:** 10.1101/2023.02.16.23286035

**Authors:** Bingxin Zhao, Yujue Li, Zirui Fan, Zhenyi Wu, Juan Shu, Xiaochen Yang, Yilin Yang, Xifeng Wang, Bingxuan Li, Xiyao Wang, Carlos Copana, Yue Yang, Jinjie Lin, Yun Li, Jason L. Stein, Joan M. O’Brien, Tengfei Li, Hongtu Zhu

## Abstract

As an anatomical extension of the brain, the retina of the eye is synaptically connected to the visual cortex, establishing physiological connections between the eye and the brain. Despite the unique opportunity retinal structures offer for assessing brain disorders, less is known about their relationship to brain structure and function. Here we present a systematic cross-organ genetic architecture analysis of eye-brain connections using retina and brain imaging endophenotypes. Novel phenotypic and genetic links were identified between retinal imaging biomarkers and brain structure and function measures derived from multimodal magnetic resonance imaging (MRI), many of which were involved in the visual pathways, including the primary visual cortex. In 65 genomic regions, retinal imaging biomarkers shared genetic influences with brain diseases and complex traits, 18 showing more genetic overlaps with brain MRI traits. Mendelian randomization suggests that retinal structures have bidirectional genetic causal links with neurological and neuropsychiatric disorders, such as Alzheimer’s disease. Overall, cross-organ imaging genetics reveals a genetic basis for eye-brain connections, suggesting that the retinal images can elucidate genetic risk factors for brain disorders and disease-related changes in intracranial structure and function.

---

The retina of the eye is the only part of the central nervous system that can be visualized without surgical intervention. As a key component of the visual pathway, the retina is synaptically connected to the visual cortex through the optic nerve, thalamus, and optic radiations. There are anatomical, physiological, and embryological similarities between the retina and the brain, in terms of cell types, vasculature, and immune responses^1^. The eye develops from the forebrain during the third week of gestation^2^. The retina is embryonically formed as part of the diencephalon, which later becomes the thalamus, thus being developmentally related to specific brain regions. Consequently, the retina has been considered a unique window into altered brain structure/function^1, 3^ and brain disorders^4^, such as Alzheimer’s disease^5–8^, Parkinson’s disease^9^, stroke^10, 11^, cerebral small vessel disease^12^, schizophrenia^13^, cognitive decline^11, 14, 15^, and many others. For example, it has been extensively studied that retinal neurodegeneration can be used as an easily accessible biomarker to identify individuals at high risk of developing Alzheimer’s disease or those with preclinical Alzheimer’s disease^6, 8, 16–18^. Retinal abnormalities have also been frequently reported in Parkinson’s disease, and animal models have demonstrated that similar molecular mechanisms underlie Parkinson’s disease pathology and neurodegeneration in parkinsonian eyes^9^. However, except for a few pairs of diseases, such as primary open-angle glaucoma and Alzheimer’s disease^19^, little is known about the shared genetic effects underlying eye-brain relationships and parallel pathological changes between the two organs.

The retina and brain images provide well-defined clinical endophenotypes for disorders of the eye and brain. Both color fundus photography and optical coherence tomography (OCT) are popular retinal imaging modalities. Retinal images serve as the gold standard screening for age-related macular degeneration^20^, diabetic retinopathy^21^, and all other pathologies involving the retina. These images offer a color picture of the back of the eye, including the retina, optic nerve head, and retinal vasculature. Retinal OCT imaging shows a high-resolution view of the cross-sectional structure of the retina^22^. In neurologic conditions, OCT imaging allows the assessment of the retinal layers’ thickness and structural changes caused by the modification of neuronal and retinal glial cells^23^. In addition, magnetic resonance imaging (MRI) captures both structural and functional characteristics of the brain, resulting in a wide range of clinical applications in neurological and neuropsychiatric disorders^24^. Recent large-scale genome-wide association studies (GWAS) have shown that both retinal imaging biomarkers^25–33^ and brain MRI traits^34–41^ are heritable, with the genetic influences of common genetic variants being identified in hundreds of genomic regions. As expected, genetic overlaps were identified between retinal imaging traits and eye disorders (such as the cupping of the optic nerve head and glaucoma^30^) as well as between brain MRI traits and brain disorders (such as functional connectivity of the visual network and Alzheimer’s disease^39^). However, few studies have used imaging genetics to study brain health from a retinal perspective. A large-scale cross-organ analysis of retinal and brain imaging traits may provide an opportunity to identify retinal imaging biomarkers for brain disorders and to uncover the genetic basis for eye-brain connections.

Using multimodal retinal and brain imaging traits from the UK Biobank (UKB) study^42^, we investigated the cross-organ genetic architecture of the eye and brain. A total of 156 retinal imaging traits were examined, of which 46 were derived from OCT images and 110 from fundus photographs. The 46 OCT derived measurements were already available in the UKB database, such as retinal thickness across layers^43, 44^ and vertical cup-to-disc ratio^29^. For the fundus images, we used 11 different pre-trained transfer learning^31^ models built from the ImageNet^45^ database to extract imaging features of the retinal structure. In each deep transfer learning model, the top 10 principal components (PCs) were considered, explaining an average of 70.71% variance (range = [50.58%, 95.84%]) in the last layer, resulting in 110 fundus image features (11 x 10). These deep-learning-based image embeddings and low-dimensional representations contain eye-specific biological information, which may not be present in standard eye measurements^31^. We conducted GWAS for these 156 (46 + 110) retinal imaging traits and then evaluated their genetic connections with 458 imaging traits from three primary brain MRI modalities, including 1) 101 regional brain volumes^35^ and 63 cortical thickness traits^46^ from structural MRI; 2) 110 diffusion tensor imaging (DTI) parameters from diffusion MRI^37^; and 3) 92 functional connectivity and activity (or amplitude) traits from resting state and task-based functional MRI (fMRI)^39^, respectively. The **Methods** section and **Table S1** provide more information on these retinal and brain imaging data. An overview of the study design and data analysis is provided in **Figure 1**. GWAS summary statistics for retinal imaging traits and our data analysis results will be made available through the eye imaging genetics knowledge portal (Eye-KP) at https://www.eyekp.org/.

**Fig. 1.**
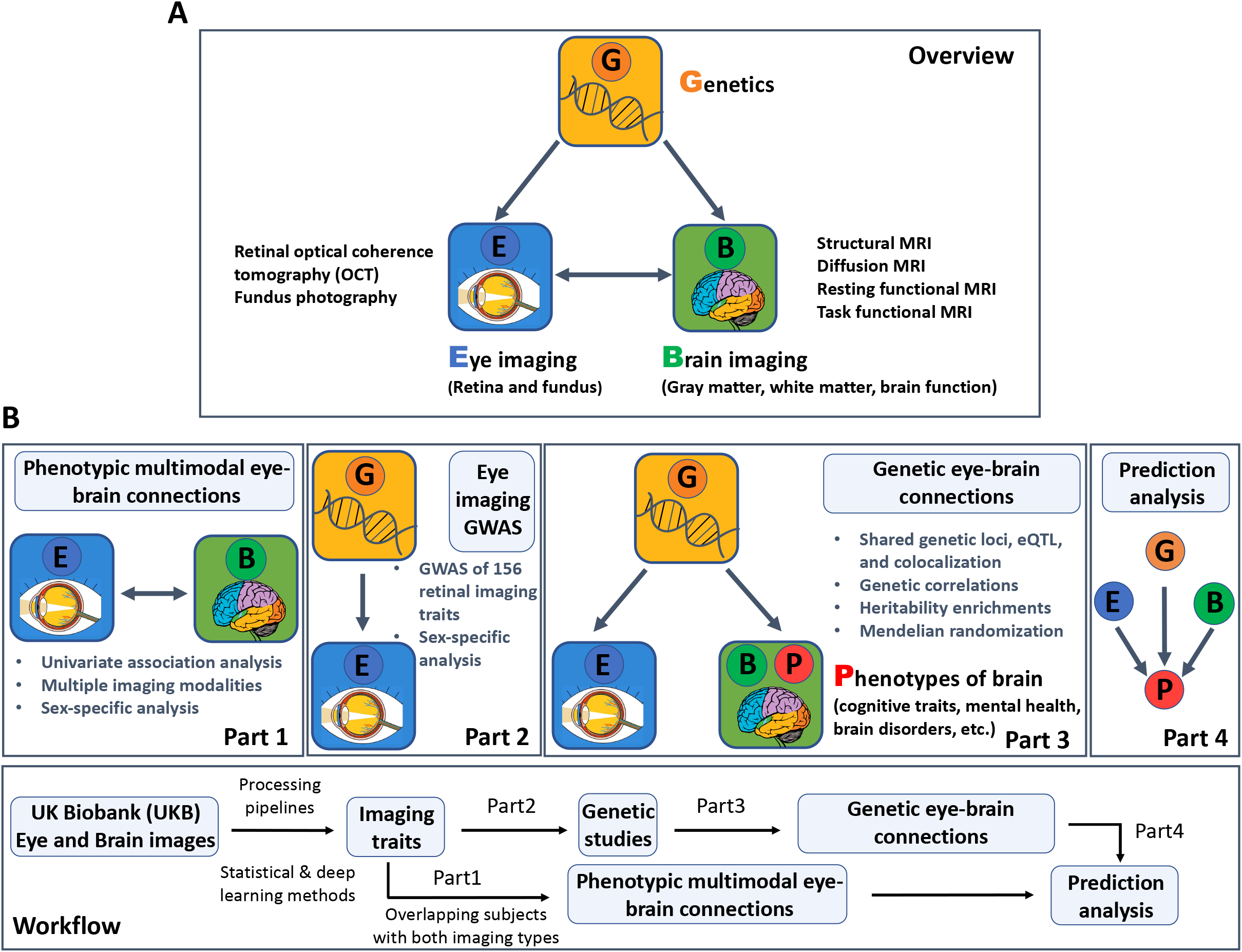
Study overview and workflow. **(A)** An overview of the study design. We used multimodal retinal and brain imaging data to understand the phenotypic and genetic connections between the brain and the eye. We considered multiple brain magnetic resonance imaging (MRI) modalities, including structural MRI, diffusion MRI, resting-state functional MRI (fMRI), and task-based fMRI. For the eye, we used traits derived from retinal optical coherence tomography (OCT) and extracted from fundus retinal images using pre-trained transfer learning models. **(B)** A brief description of the overall workflow and major analyses in each part.

## RESULTS

### Phenotypic multimodal eye-brain connections

We examined phenotypic associations between 156 retinal imaging traits and 458 brain MRI traits after adjusting for a wide variety of vascular risk factors^3^ and imaging confounders^34^, as well as body size, age, and sex effects (see the **Methods** section for the complete list of adjusted covariates). For discovery, we analyzed data of UKB white British individuals (average *n* = 6,454 across different modalities). At the false discovery rate (FDR) level of 5% (by the Benjamini-Hochberg procedure, *P* < 4.37 × 10^-4^, 156 × 458 tests), we identified 625 associations (**Figs. 2A** and **S1**), 135 of which were replicated in a hold-out independent validation dataset (average *n* = 959) with concordant association signs (**Fig. S2**). Among the 625 associations, 121 further survived the conservative Bonferroni significance level (*P* < 6.99 × 10^-7^), and 66 can be replicated in the same hold-out independent dataset. These significant results were mainly related to multiple brain structural modalities, including regional brain volumes, cortical thickness, and DTI parameters. They were broadly related to both OCT measures and fundus image features (**Table S2**). Below we summarized the patterns of associations that have been replicated.

**Fig. 2.**
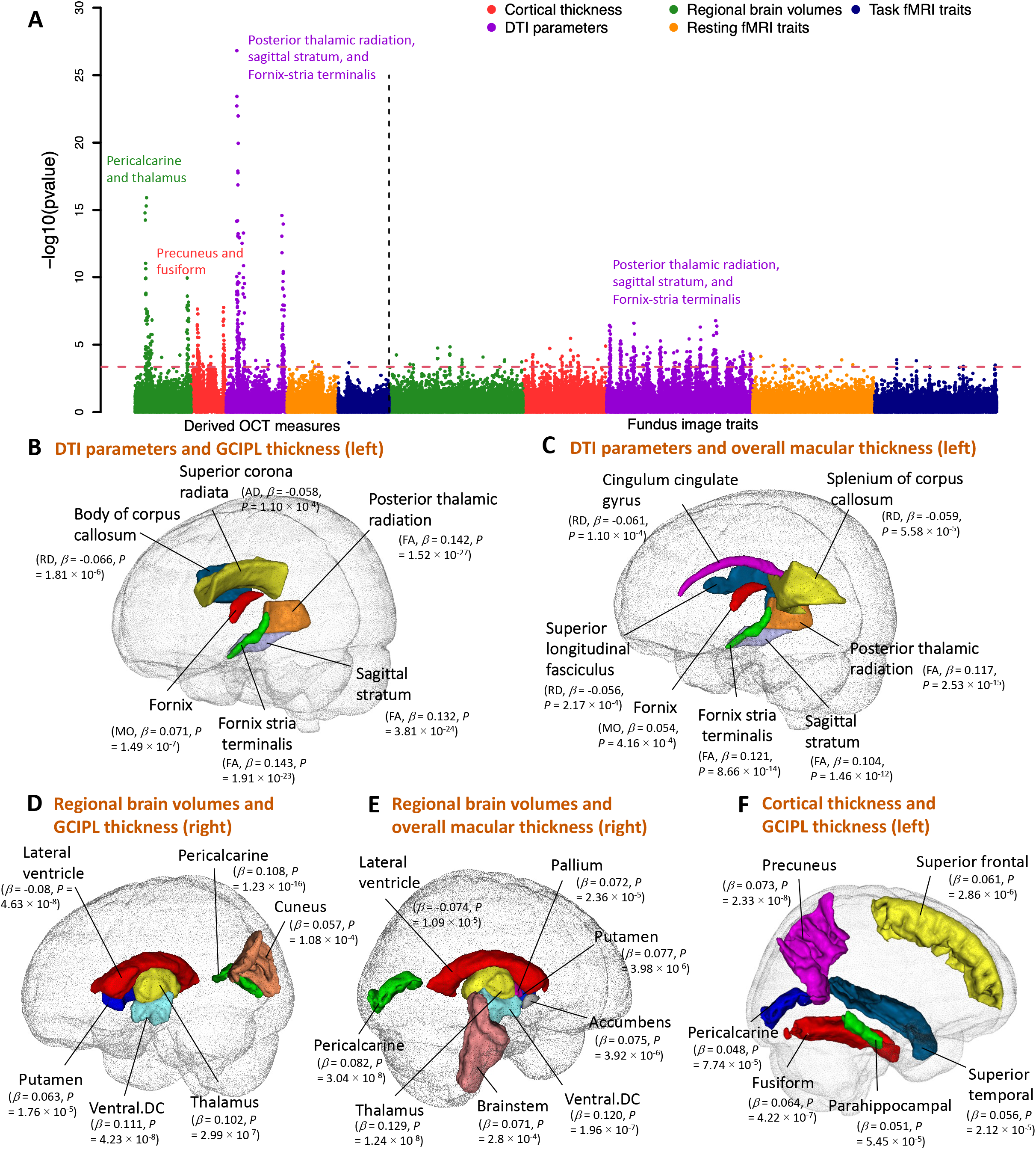
Phenotypic eye-brain imaging associations. **(A)** This figure shows the -log10(p-value) of testing the associations between 156 retinal imaging traits (46 derived OCT measures and 110 fundus image traits) and 458 brain MRI traits, including 101 regional brain volumes, 63 cortical thickness traits, 110 diffusion tensor imaging (DTI) parameters, 92 resting fMRI traits, and 92 task fMRI traits. **Table S1** provides more information on these imaging traits. The red dashed horizontal line indicates the Benjamini-Hochberg FDR 5% significance level (raw *P* < 4.37 × 10^-4^). Each brain imaging modality is labeled with a different color. We have also labeled the brain structures that had the strongest associations in each modality. **(B-C)** Location of the white matter tracts whose DTI parameters were significantly associated with **(B)** the thickness of the ganglion cell and inner plexiform layer (GCIPL, left eye) and **(C)** the overall thickness of the macula (left eye). AD, axial diffusivity; RD, radial diffusivity; MO, mode of anisotropy; and FA, fractional anisotropy. **(D-E)** Location of the brain regions whose volumes were significantly associated with **(D)** the thickness of GCIPL (right eye) and **(E)** the overall thickness of the macula (right eye). **(F)** Location of the cortical brain regions whose thickness was significantly associated with the thickness of GCIPL (left eye).

Thicknesses of the macula^44^, the retinal nerve fiber layer (RNFL)^43^, and the ganglion cell and inner plexiform layer (GCIPL)^43^ consistently had positive associations with the fractional anisotropy (FA) of multiple white matter tracts, including those related to the visual pathway (**Figs. 2B-C** and **S3**). Various eye diseases are associated with retinal thinning^47^. Moreover, previous studies have consistently demonstrated the thinning of the RNFL and GCIPL to be associated with cerebrovascular diseases^48^ and early-stage Alzheimer’s disease^49^. These results suggest a parallel relationship between retinal and brain health, as well as changes in brain white matter that may be related to both. The strongest associations were observed between the GCIPL thickness of the left eye and the mean FA of the posterior thalamic radiation, sagittal stratum, and fornix-stria terminalis tracts (*β* > 0.142, *P* < 1.91 × 10^-23^). The posterior thalamic radiation overlaps with the optic radiation in the visual pathway, which links the lateral geniculate nucleus to the primary visual cortex, transmitting visual input from the eye. There were also similar associations between fundus image features and DTI parameters, although the associations were weaker than those for retinal thickness traits (**Fig. S3**). Moreover, thickness measures of the RNFL, GCIPL, macula, and inner nuclear layer (INL)^43^ were positively associated with volumes of multiple brain cortical and subcortical structures, including the pericalcarine, thalamus, pallidum, and putamen (**Figs. 2D-E** and **S4**). The pericalcarine is the location where the primary visual cortex (V1) concentrates, and we found that the regional brain volumes of the pericalcarine had consistent positive associations with the RNFL, GCIPL, and macular thickness (*β* > 0.052, *P* < 5.90 × 10^-5^). We also found positive associations with brain structures in the dorsal and ventral visual pathways that extended from the primary visual cortex, such as the cuneus (*β* > 0.057, *P* < 1.08 × 10^-4^). The thalamus and macular are both derived from the diencephalon. Positive associations between regional brain volumes of the thalamus and macular thickness were found (*β* > 0.120, *P* < 1.24 × 10^-8^), emphasizing their developmental origins. Negative associations between retinal layer thickness and enlargement of the lateral ventricles were also detected. The left and right hemispheres of the brain demonstrated consistent associations with retinal imaging traits. For example, the left and right brain thalamus volumes were significantly correlated with the thicknesses of the macular and GCIPL in both eyes (*β* > 0.120, *P* < 1.24 × 10^-8^). The GCIPL thickness was also positively associated with global and regional brain cortical thickness measures, including the primary visual cortex (the pericalcarine, *β* = 0.048, *P* = 7.74 × 10^-5^). The top two regions with the strongest links were the precuneus, which is in the dorsal visual pathway (*β* = 0.073, *P* = 2.33 × 10^-8^), and the fusiform, which is in the ventral visual pathway (*β* = 0.064, *P* = 4.22 × 10^-7^, **Figs. 2F** and **S4**),

We repeated the above analyses separately for females and males to examine the sex-specific patterns (average *n* = 3,338 and 3,150, respectively). At the FDR 5% level (*P* < 4.37 × 10^-4^), 53 associations were identified in both females and males, the female sample identified 191 additional associations, and 62 more were only found in males. The additional associations found in analyses that included only females or males were primarily related to fundus image traits. Specifically, the female analysis showed more significant associations with DTI parameters, while the male analysis revealed more significant associations with cortical thickness measures (**Figs. S5-S7**). For OCT measures, males and females demonstrated similar eye-brain association patterns, although the number of significant pairs that survived multiple testing adjustments varied between the two samples. For example, the mean FA of the fornix-stria terminalis, posterior thalamic radiation, and sagittal stratum tracts was associated with the thickness of RNFL, GCIPL, and macula in both males and females, with more significant pairs being identified in females (**Fig. S8**). These retinal thickness traits were also consistently associated with volumes of the pericalcarine, thalamus, and accumbens regions in sex-specific analysis (**Fig. S9**). In summary, although only a relatively small percentage of subjects had both brain and retinal imaging data in the UKB study, we uncovered that the retinal imaging biomarkers, such as the thickness of different retinal layers, were associated with smaller brain volumes, reduced cortical thickness, and weaker white matter structural connections in the brain. Many retina-related brain structural variations were observed in the primary visual cortex and other structures in the visual pathways. Our results are consistent with previous studies that have found parallel changes in eye-brain structures during pathological progression^1, 3^, as well as providing further information on the most relevant brain MRI modalities and biomarkers for clinical applications and future research.

### GWAS for 156 retinal imaging traits

Based on UKB individuals of white British ancestry^50^, we estimated the proportion of phenotypic variance explained by single nucleotide polymorphisms (SNPs) for the 156 retinal imaging traits (average *n* = 60,748). The average SNP-based heritability (*h^2^*) was 42.21% for the 46 OCT measures (*h^2^* range = (19.28%, 68.28%)), all of which were significant at FDR 5% level (**Fig. S10** and **Table S3**). Of the 110 fundus image traits, 90 were significant at FDR 5% level, with the mean *h^2^* being 19.27% (*h^2^* range = (4.06%, 42.75%))). In each of the 11 transfer learning models, at least seven of the 10 PCs had significant *h^2^*. Additionally, we estimated *h^2^* separately for females and males, and the results were highly consistent between the two sexes (mean *h^2^* = 24.83% among females vs. 23.33% among males, correlation = 0.972, *P* = 0.457, **Fig. S11**).

We conducted GWAS based on the same white British cohort to uncover the genetic architecture of the 156 retinal imaging traits (average *n* = 60,748). QQ and Manhattan plots can be viewed on our server (http://165.227.78.169:443/) developed via PheWeb^51^. Linkage disequilibrium score regression (LDSC) intercepts^52^ were all near one, indicating that no confounding factor resulted in the genomic inflation of test statistics (average = 1.004, range = (0.974, 1.031)). At a stringent GWAS significance level 3.20 × 10^-10^ (5 × 10^-8^/156, that is, the standard GWAS significance level after further considering Bonferroni-type adjustment for 156 retinal imaging traits), we identified independent (linkage disequilibrium [LD] *r^2^* < 0.1) significant genetic associations in 258 genomic regions (cytogenetic bands). Significant associations were found for all 46 OCT measures and 91 of the 110 fundus image traits (**Fig. S12** and **Table S4**). We also estimated these significant genetic effects separately for males and females in a sex-specific analysis. We found that the genetic effects were highly consistent in both sexes (correlation = 0.975, *P* = 0.76, **Fig. S13**).

We replicated our GWAS results using independent European and non-European datasets (**Methods**). First, we performed GWAS of the 156 retinal imaging traits using the UKB European ancestry but non-British subjects (average *n* = 5,320). For the 4,329 identified independent (LD *r^2^* < 0.1) image-variant associations in 258 genomic regions, 1,630 (37.65%, in 162 genomic regions) passed the FDR 5% significance level in this European validation GWAS, and 2,210 (51.05%, in 189 regions) were significant at the nominal significance level (0.05) (**Fig. S14** and **Table S5**). Most of the significant genetic effects (2,207/2,210, in 188 regions) had concordant directions in the two independent GWAS, with the correlation of their genetic effects being 0.958 (**Fig. S15**). Among the 188 replicated genomic regions, 146 were associated with OCT measures, and 103 were associated with fundus image traits. These results suggest the high generalizability of our GWAS findings in European samples. Next, we repeated the validation GWAS on the non-European UKB subjects (average *n* = 6,490) and found that 25.18% (1,090/4,329, in 142 regions) associations were significant at the nominal significance level, most of which (1,068/1,090, in 140 regions) had the same genetic effect directions as the discovery GWAS (**Fig. S16**). Overall, 107 replicated regions were observed for OCT measures and 51 for fundus image traits in non-European validation analysis.

We have also developed polygenic risk scores (PRS) via PRS-CS^53^ to evaluate the out-of-sample prediction performance of our discovery GWAS results (**Methods**). The PRS for 133 of the 156 retinal imaging traits were significant at FDR 5% level (*P* range = (6.90 × 10^-98^, 3.82 × 10^-2^), **Table S6** and **Fig. S17**), with the mean incremental R-squared being 2.51% (S.E. = 2.29%). A total of 22 traits had R-squared greater than 5%. The highest prediction accuracy was observed on a set of traits related to the INL^43^, such as the thickness from the INL to the retinal pigment epithelium (RPE) (R-squared = 9.46% and 8.43% for right and left eyes, respectively) and the thickness between the INL to the external limiting membrane (ELM) (R-squared = 7.68% and 7.20% for right and left eyes, respectively). To evaluate the transferability of GWAS findings, we also examined the PRS performance on non-European UKB subjects. We found that 101 retinal imaging PRS had significant prediction performance in the non-European UKB dataset at FDR 5% level (*P* range = (2.77 × 10^-65^, 2.88 × 10^-2^)). The average incremental R-squared of these significant PRS was 1.39% (S.E. = 1.45%), which was significantly lower than their corresponding performance in European dataset (*P* = 7.89 × 10^-9^). These results demonstrate the capability of our GWAS summary statistics in out-of-sample analyses and also illustrate the challenge of cross-population genetic prediction.

### Genetic underpinnings of eye-brain connections in 65 genomic loci

We examined eye-brain genetic pleiotropy in 188 replicated genomic regions of retinal imaging traits that had concordant genetic effect directions in the discovery and validation GWAS. First, for the retinal imaging-significant genetic variants and those in LD with them (*r*^2^ ≥ 0.6), we symmetrically searched for GWAS signals that have been identified to be associated with brain MRI traits^35, 37, 39, 46^. Second, we performed association lookups in the NHGRI-EBI GWAS catalog^54^ to identify shared genetic influences between retinal imaging traits and brain-related complex traits and diseases (**Methods**). In 65 of these 188 genomic regions, we found genetic overlaps between the retinal imaging traits and brain phenotypes, 47 of which had also been linked to various eye traits and conditions, such as glaucoma^55^, refractive error^56^, advanced age-related macular degeneration^57^, and cataracts^58^. Specifically, we found genetic pleiotropy for a wide range of brain traits and disorders, including stroke, Parkinson’s disease, Alzheimer’s Disease, glioma/glioblastoma, neuropsychiatric disorders, migraine, mental health, and cognitive traits (**Fig. 3** and **Table S7**). Shared genetic influences were also identified in 18 regions with different brain MRI modalities, including 10 regions with regional brain volumes^35^, 9 regions with DTI parameters^37^, 3 regions with cortical thickness traits^46^, and 2 regions with resting fMRI traits^39^ (**Fig. S18**). Using Bayesian colocalization analysis^59^, we examined whether there were common causal genetic variants underlying the overlapping genetic signals between retinal structures and brain phenotypes (posterior probability of the shared causal variant hypothesis [PPH4] > 0.8^59, 60^). In addition, we found that many retinal imaging-significant genetic variants were expression quantitative trait loci (eQTLs) reported in large-scale eQTL studies of brain tissues^61^. Our results are summarized in **Table S8**, with selected eye-brain trait pairs being displayed in **Figures 4-5** and **S19-S79.** Below we have provided more details for each brain MRI modality and major brain phenotype category.

**Fig. 3.**
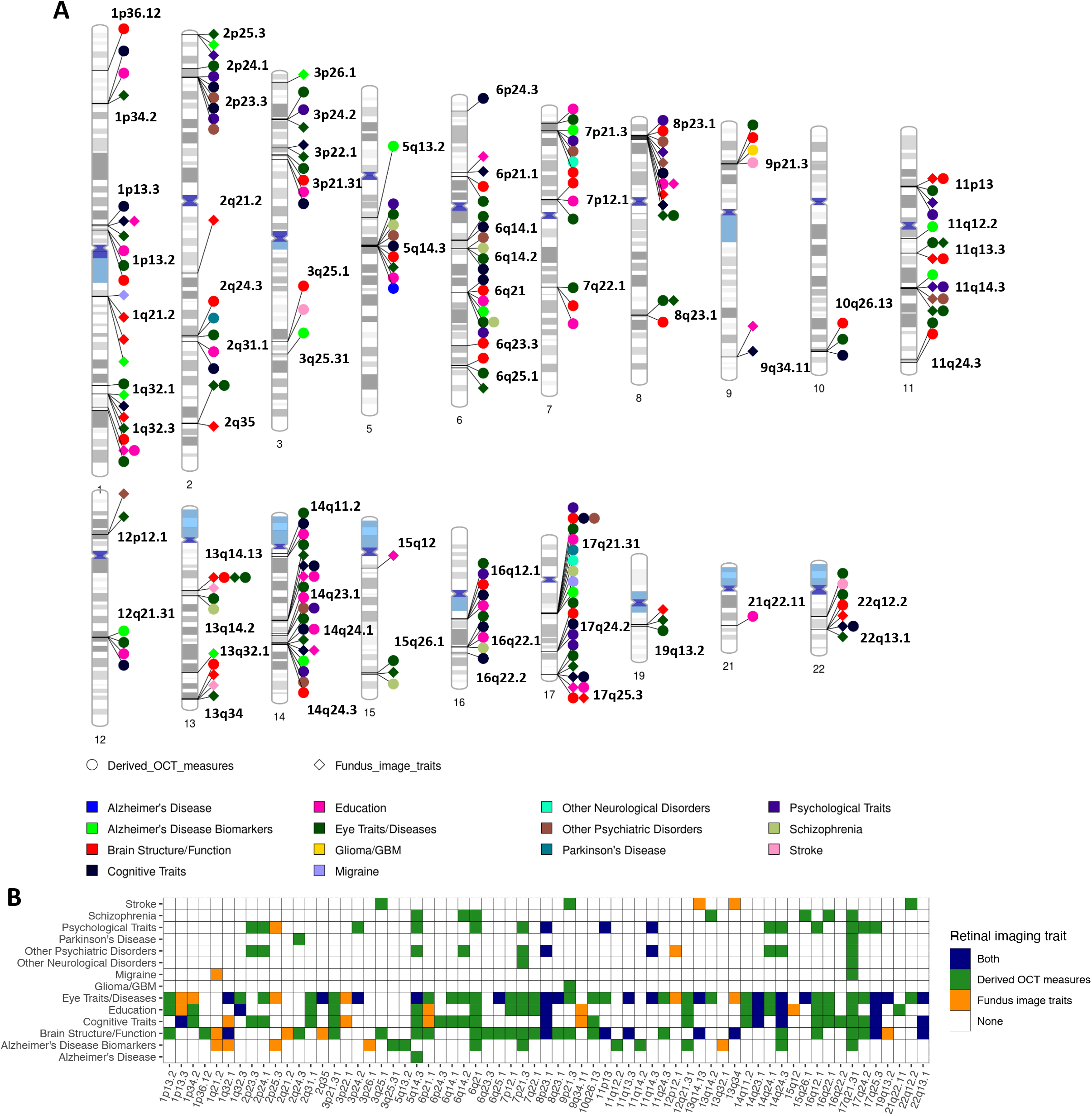
Genomic loci associated with both eye imaging traits and brain-related complex traits and diseases. **(A)** Ideogram of genomic regions (names are in black) influencing both retinal imaging traits and brain-related complex traits and diseases, including the phenotypes reported on the NHGRI-EBI GWAS catalog (https://www.ebi.ac.uk/gwas/) and the brain MRI traits available on BIG-KP (https://bigkp.org/). Each category of brain phenotypes is labeled with a different color, and we use different shapes for OCT measures and fundus image traits. **(B)** Table summary, where the *x*-axis represents the genomic regions and *y*-axis displays the category of brain phenotypes. Derived OCT measures and fundus image traits are labeled with different colors, and a third color is used when both are observed in the locus.

**Fig. 4.**
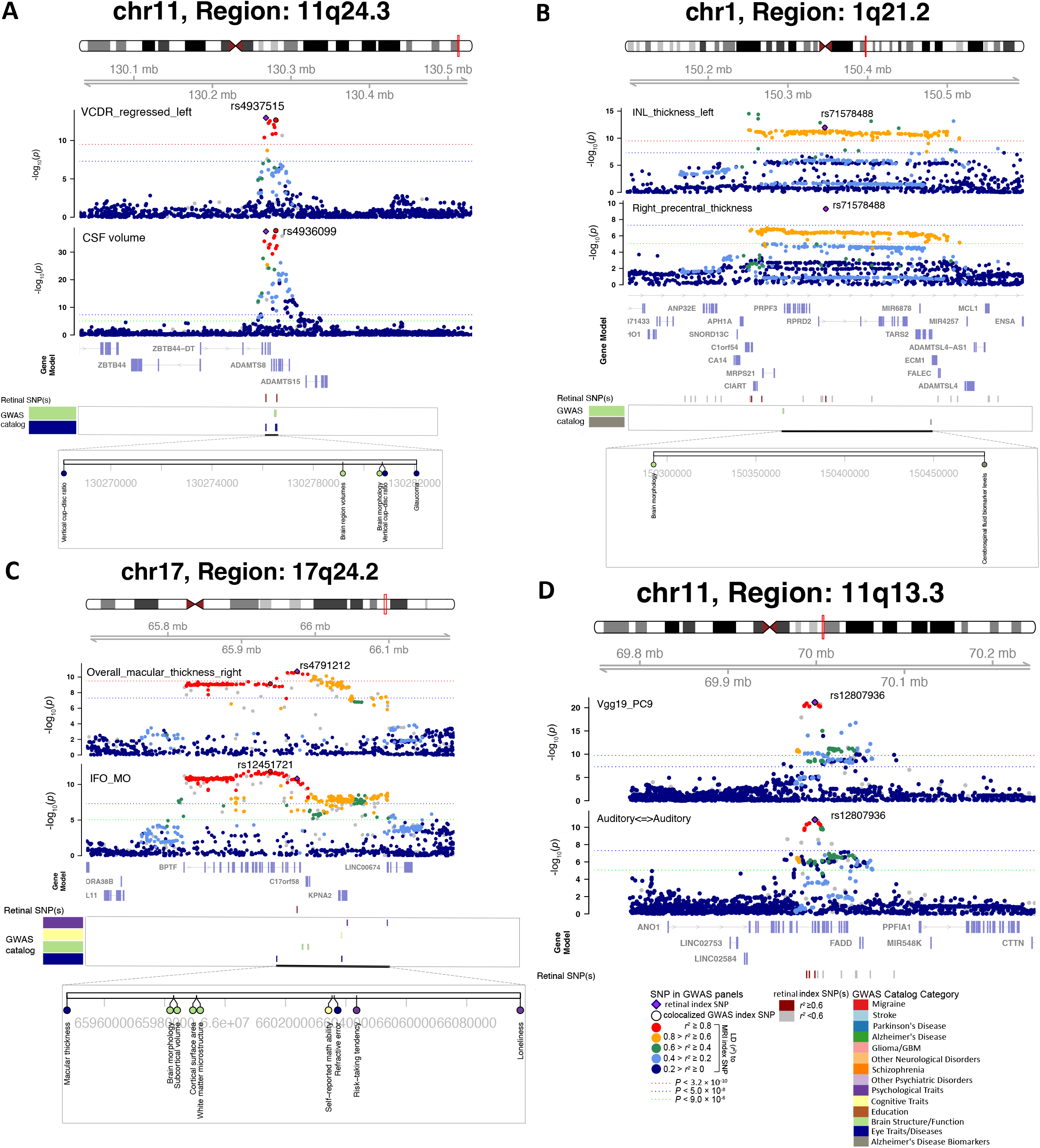
Selected genetic loci that were associated with both eye and brain imaging traits. **(A)** In 11q24.3, we observed shared genetic influences between the vertical cup-to-disc ratio (regressed on disc diameter, left eye, VCDR_regressed_left, index variant rs4937515) and the cerebrospinal fluid volume (CSF volume, index variant rs4936099). Bayesian colocalization analysis suggested the shared causal variant between the two traits (posterior probability PPH4 = 0.997). **(B)** In 1q21.2, we observed shared genetic influences between the inner nuclear layer (INL) thickness (left eye, INL_thickness_left) and the cortical thickness of the right precentral brain region (Right_precentral_thickness, shared index variant rs71578488, PPH4 = 0.562). In this region, the INL_thickness_left was also in LD (*r*^2^ ≥ 0.6) with cerebrospinal fluid biomarker levels. **(C)** In 17q24.2, we observed shared genetic influences between the overall macular thickness (right eye, overall_macular_thickness_right, index variant rs4791212) and the mean MO of the inferior fronto-occipital fasciculus (IFO_MO, index variant rs12451721, PPH4 = 0.963). We also observed genetic overlaps (LD *r*^2^ ≥ 0.6) with self-reported math ability, risk-taking tendency, and loneliness. **(D)** In 11q13.3, we observed shared genetic influences between the ninth PC of the Vgg19 model on fundus image (Vgg19_PC9) and the functional connectivity within the auditory network (Auditory<=>Auditory, shared index variant rs12807936, PPH4 = 0.994).

**Fig. 5.**
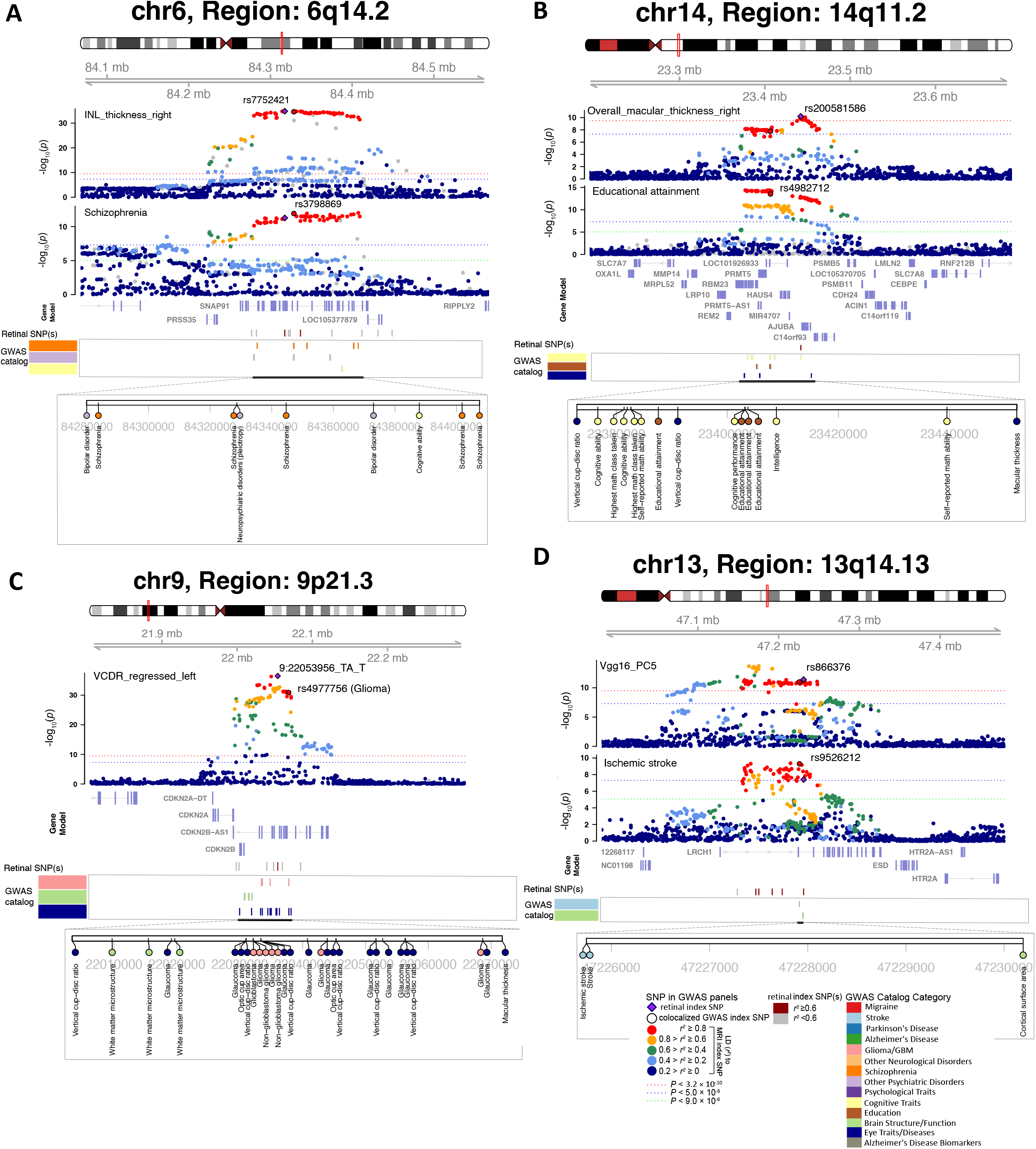
Selected genetic loci that were associated with both eye and brain-related complex traits and disorders. **(A)** In 6q14.2, we observed shared genetic influences between the inner nuclear layer (INL) thickness (right eye, INL_thickness_right, index variant rs7752421) and schizophrenia (index variant rs3798869). Bayesian colocalization analysis suggested the shared causal variant between the two traits (posterior probability PPH4 = 0.952). In this region, the thickness of INL was also in LD (*r*^2^ ≥ 0.6) with bipolar disorder and cognitive ability. **(B)** In 14q11.2, we observed shared genetic influences between the overall macular thickness (right eye, overall_macular_thickness_right, index variant rs200581586) and educational attainment (index variant rs4982712, PPH4 = 0.764). In this region, the overall macular thickness was also in LD (*r*^2^ ≥ 0.6) with intelligence and cognitive ability. **(C)** In 9p21.3, we observed shared genetic influences between the vertical cup-to-disc ratio (regressed on disc diameter, left eye, VCDR_regressed_left, index variant 9:22053956_TA_T) and Glioma (index variant rs4977756). We also observed genetic overlaps (LD *r*^2^ ≥ 0.6) with self-reported math ability, risk-taking tendency, and loneliness. **(D)** In 13q14.13, we observed shared genetic influences between the fifth PC of the Vgg16 model on fundus image (Vgg16_PC5, index variant rs866376) and ischemic stroke (index variant rs9526212, PPH4 = 0.994).

Brain volumetric measures had genetic overlaps with retinal structures in 10 genomic loci (LD *r*^2^ ≥ 0.6, **Figs. 4A** and **S19-27**). For example, shared genetic components between cerebrospinal fluid (CSF) volume and vertical cup-to-disc ratio^62^ were found in 11q24.3 (**Fig. 4A**). The retinal index variant rs4937515 was an eQTL of *ADAMTS8* in brain tissues^61^, and there was strong evidence of shared causal genetic variants between the two traits (PPH4 = 0.997). The rs4937515 was also in LD (*r*^2^ ≥ 0.6)^63^ with known genetic risk variants of glaucoma (index variant rs2875238)^55^. The CSF is the primary fluid within the central nervous system, and the biological role of CSF pressure has been well-established in glaucoma and other ophthalmic diseases^64, 65^. Our results provide further evidence of genetic links underlying the connections between CSF and eye disorders. Colocalizations between retinal imaging traits and brain volumes were also observed in 8q23.1 (e.g., right thalamus), 22q13.1 (e.g., left lateral ventricle), 17q24.2 (e.g., left caudal anterior cingulate), 6q25.1 (e.g., right hippocampus), and 7q22.1 (e.g., right accumbens area). In these regions, retinal imaging traits also tagged (LD *r*^2^ ≥ 0.6) schizophrenia, major depressive disorder, neuroticism, and cognitive traits. In addition, genetic overlaps between retinal structures and cortical thickness traits were found in 3 loci (17q21.31, 8p23.1, and 1q21.3), where several brain structures in the visual pathways were involved, including the precentral, supramarginal, fusiform, and precuneus (**Fig. 4B**). In summary, we identify locus-specific genetic overlaps between the thickness of different retinal layers and the morphometry of multiple brain regions, which play essential roles in cognitive functions and are affected in various brain disorders.

Retinal structures also had widespread genetic pleiotropy with brain structural and functional connectivity. Retinal imaging traits and DTI parameters had shared genetic influences in 9 genomic regions, 7 of which had strong evidence of colocalization (**Figs. 4C** and **S28-35**). For example, the overall macular thickness^44^ and the mode of anisotropy (MO)^66^ of the inferior fronto-occipital fasciculus tract had common causal genetic variants in 17q24.2 (PPH4 = 0.816, **Fig. 4C**). The inferior fronto-occipital fasciculus is the longest associative white matter tract that connects various brain areas and involves multiple functions. Thinner retinal layers had close relationships with the reduced volume and worse microstructural integrity of the brain’s white matter^67^. These results provide genetic insights into retina-white matter connections. Both retinal imaging traits and DTI parameters also overlapped genetically with cognitive traits (such as intelligence in 22q13.1), psychiatric disorders (such as in 14q24.3), and eye disorders (such as advanced age-related macular degeneration in 17q25.3). We also found shared genetic influences between functional connectivity of resting fMRI and retinal imaging traits in 11q13.3 and 17q21.31 (**Figs. 4D** and **S36**).

In addition, many genomic regions associated with retinal imaging traits have been linked to brain-related complex traits and diseases in previous GWAS. In 6q14.2, 6q21, 13q14.2, 15q26.1, and 16q22.1 regions, the thickness of different retinal layers was in LD (*r*^2^ ≥ 0.6) with schizophrenia^68–70^ (**Figs. 5A** and **S37-S40**). For example, the INL thickness had shared causal genetic variants with schizophrenia (PPH4 = 0.952). The retinal index variant (rs7752421) was an eQTL of *SNAP91* in human brain tissues^61^, affecting gene expression levels in the brain. In excitatory neurons, synaptic defects are increasingly associated with schizophrenia, and altered expression of *SNAP91* has been observed to impact synaptic development^71^. As schizophrenia patients often report visual perception changes, OCT measures of the retinal structure have received increasing attention in schizophrenia research^72^. The identified genetic links in our analysis support the use of retinal layer assessments as potential biomarkers for schizophrenia. Retinal structures were also in LD (*r*^2^ ≥ 0.6) with other neuropsychiatric disorders and mental health traits, such as bipolar disorder^73^, anxiety^74^, depressive symptoms^75^, neuroticism^76^, subjective well-being^77^, and risk-taking tendency^78^ (**Figs. S41-S49**). For example, the thickness of various retinal layers had shared genetic influences with neuroticism in multiple regions (**Figs. S41-S47**).

There were 23 genomic regions associated with cognitive traits (such as intelligence^79^, cognitive performance^80^, general cognitive ability^81^, and reaction time^82^, **Figs. S50-S57**) and/or educational attainment^83^ (**Figs. 5B** and **S58-S72**). Several studies have reported that retinal layer thickness may be a prognostic biomarker of cognitive impairment and long-term cognitive decline in older individuals^84, 85^. Retinal imaging traits were in LD (*r*^2^ ≥ 0.6) with multiple neurodegenerative disorders, such as in 2q24.3, 17q21.31, 8p23.1, and 15q12 with Parkinson’s disease^86^ (**Fig. S73**); in 17q21.31 with corticobasal degeneration^87^; in 7p21.3 with frontotemporal dementia^88^; in 5q14.3, 17q21.31, 6p12.1 with Alzheimer’s disease^89^, and several more loci (such as 1q32.1, 2p25.3, and 11q14.2) with biomarkers of Alzheimer’s disease^90^ (**Figs. S74-76**). Genetic overlaps with other brain diseases were also observed. For example, vertical cup-to-disc ratio^29^ was in LD (*r*^2^ ≥ 0.6) with glioma/ glioblastoma^91^ and white matter microstructure in 9p21.3 (**Fig. 5C**). Glioma may affect the optic nerve (optic nerve glioma), which is the most common primary neoplasm of the optic nerve^92^. Retinal imaging traits also had shared genetic effects with migraine/headache^93^ in 5 regions and cerebrovascular diseases in 9 regions, including stroke^94^, Moyamoya disease^95^, intracranial aneurysm^96^, and cerebral aneurysm^97^ (**Figs. 5D** and **S77-79**). In summary, our findings indicate close genetic connections between the eye and the brain. Abnormalities in retinal structure may provide insight into the genetic risk of neurodegenerative diseases and neuropsychiatric disorders.

### Genetic correlation and heritability enrichment patterns

We examined genetic correlations (GC) between 156 retinal imaging traits and 39 sets of publicly available GWAS summary statistics of brain-related complex traits and diseases using cross-trait LDSC^98^ (**Table S9**). At FDR 5% level (*P* < 2.06×10^-3^), we observed 246 significant genetic correlation pairs between 69 retinal imaging traits and 21 brain phenotypes, including brain disorders, cognitive traits, and mental health traits (**Fig. S80**).

For example, multiple cognitive traits (such as cognitive function, numerical reasoning, intelligence, and cognitive performance) and education had consistent positive genetic correlations with the thickness of RNFL^43^ and the overall thickness between the ELM to the inner and outer photoreceptor segments (ISOS)^43^ as well as their subfields (**Fig. 6A**). Consistent with our results, previous clinical studies have identified RNFL thickness to be phenotypically related to global cognitive score, executive function, and verbal function^99–101^. These studies examined RNFL thickness as a possible early biomarker of cognitive decline, whose thinning suggests axonal loss during the neurodegenerative process of the brain^101, 102^. On the other hand, negative genetic correlations with cognitive traits were observed for the thickness of GCIPL, INL, and RPE^43^, as well as disc diameter^29^. The negative correlations between GCIPL thickness and cognitive traits were also in line with one recent study on patients with Alzheimer’s disease, where GCIPL thickening correlated with poor cognition in Alzheimer’s disease^103^. One hypothesis on the intrinsic mechanism for its thickening suggested that pathological amyloid β (Aβ) accumulation and neuroinflammation of retinal ganglion cells (RGCs) contributed to the thickening of GCIPL^103^, which was supported by a parallel study on RGCs in mouse model^104^. The thickness between the ISOS and RPE and their subfields were negatively associated with depression and neuroticism. There were also negative associations between depression symptoms and vertical cup-to-disc ratio^29^ (GC < -0.166, *P* < 7.72 × 10^-4^), between cross disorder (five major psychiatric disorders^105^) and disc diameter (GC = -0.116, *P* = 1.33 × 10^-3^), and between the RNFL thickness and cannabis use disorder (GC = -0.174, *P* = 1.29 × 10^-3^). In addition, we found positive genetic correlations between RPE thickness and attention-deficit/hyperactivity disorder (ADHD) as well as between the INL thickness and stroke (GC > 0.149, *P* < 1.88 × 10^-3^). For fundus imaging traits, there were also widespread genetic correlations with the above brain phenotypes identified by OCT measures, such as ADHD, cannabis use disorder, cognitive traits, and cross disorder. In addition, fundus imaging traits had higher correlations with schizophrenia (|GC| = 0.133, *P* = 1.10 × 10^-3^), major depressive disorder (|GC| = 0.316, *P* = 1.92 × 10^-3^), and risk tolerance (|GC| = 0.097, *P* = 1.92 × 10^-3^). These results demonstrated the genome-wide genetic similarity between retinal structures and brain disorders and traits.

**Fig. 6.**
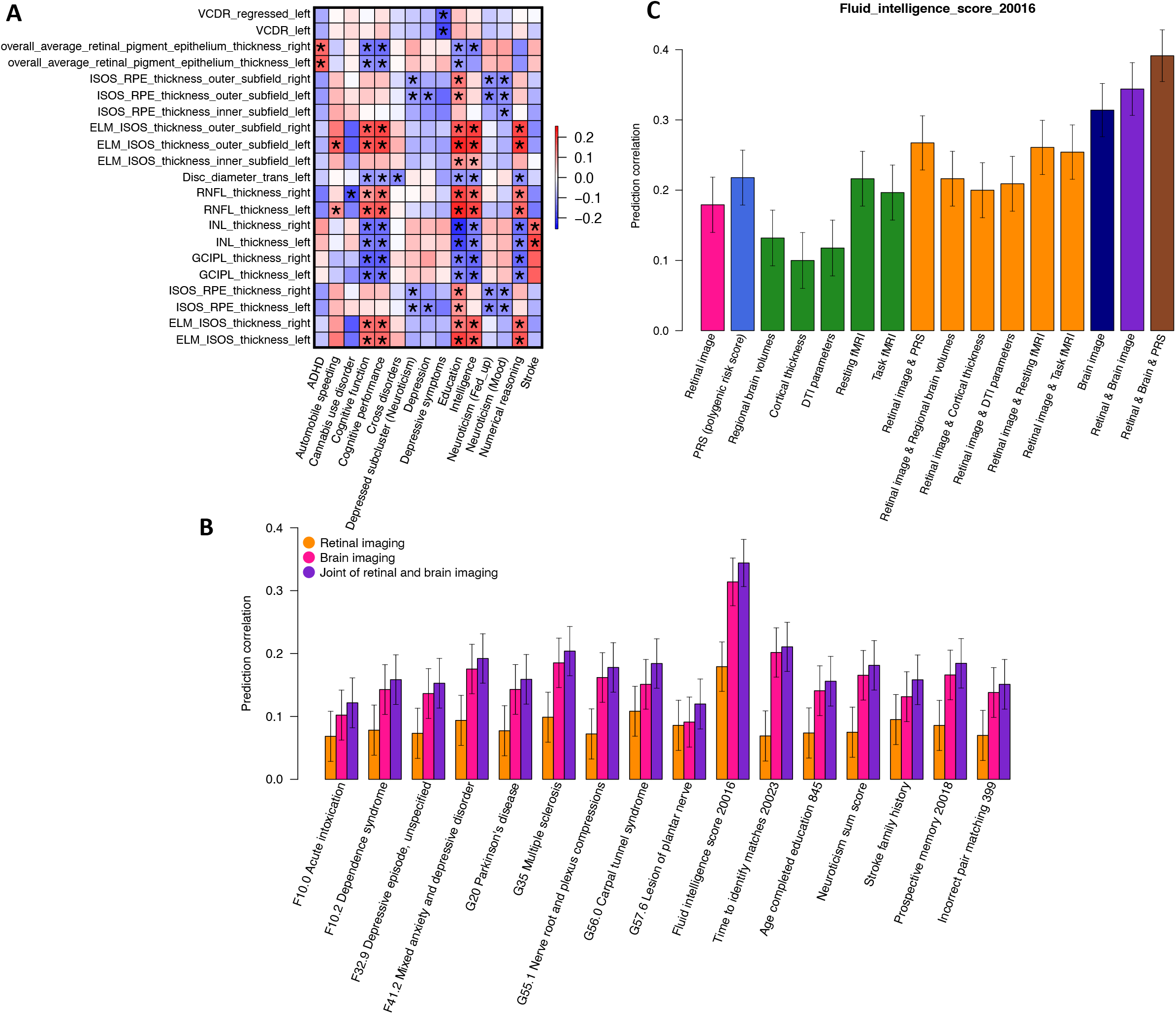
Genetic correlations and prediction analysis. **(A)** We show the selected genetic correlations between brain-related complex traits and diseases (*x*-axis) and retinal imaging traits (*y*-axis). The asterisks highlight significant genetic correlations after adjusting for multiple testing using the Benjamini-Hochberg procedure to control the FDR at 5% level. The colors represent genetic correlations. **Table S1** provides more information on these retinal imaging traits. ADHD, attention-deficit/hyperactivity disorder. **(B)** Predicting brain phenotypes using both retinal and brain imaging traits. **(C)** The accuracy of fluid intelligence prediction using multiple data types. Retinal image, including all retinal imaging traits. DTI parameters, diffusion tensor imaging parameters; Brain image, including all brain imaging modalities.

We also performed partitioned heritability analysis^106^ via LDSC to identify the tissues and cell types where genetic variation led to changes in retinal imaging traits. First, we examined a wide variety of tissue and cell type-specific regulatory elements from the Roadmap Epigenomics Consortium^107^. Among all tissue and cell types, the strongest heritability enrichments were observed in active gene regulatory regions of multiple brain tissues (**Fig. S81** and **Table S10**). Next, we repeated the partitioned heritability analysis using chromatin accessibility data from neurons (NeuN+) and glia (NeuN-) sampled from 14 cortical and subcortical brain regions^108^. We observed that the heritability of retinal structures had consistently stronger enrichment in brain glial regulatory elements than neuronal regulatory elements (**Fig. S82**). These heritability enrichments suggest that genetic variants associated with retinal structures may also alter the function of regulatory elements in brain tissues, especially glial cells, indicating the genomic links between the eye and the brain.

### Genetic causal links with brain disorders

We applied Mendelian randomization (MR) with GWAS summary statistics from the FinnGen database^109^ to examine the directional relationships between retinal structure and brain disorders. We used eight different MR methods^110–117^, and prioritized significant results that passed the Bonferroni adjustment of multiple testing in at least two methods (**Methods**). The results presented below have also passed several robustness tests, such as the MR-Egger intercept test for pleiotropy^118^.

Causal genetic effects were found between retinal imaging traits and brain disorders in both directions, and these results suggested close relationships between retinal structures and Alzheimer’s disease (**Table S11**). For example, causal genetic effects from Alzheimer’s disease to retinal structures were identified in multiple OCT measures and fundus imaging traits, including the thickness of INL (*β* > 0.025, *P* < 4.74 × 10^-5^) and the central subfield between the ISOS and RPE (*β* > 0.027, *P* < 1.12 × 10^-5^). We also observed causal effects from psychiatric diseases and other degenerative diseases of the nervous system to retinal structures, such as the INL thickness (*β* > 0.040, *P* < 3.56 × 10^-7^). These newly established positive causal effects between psychiatric diseases and INL thickness can be linked to the identified negative genetic correlations between INL thickness and cognitive traits in our previous section. There was a similar conclusion reached in previous studies regarding the thickness of the RNFL, whose thinning was indicative of cognitive decline^119, 120^. On the contrary, other recent studies have also noted correlations between INL thickening and brain-related diseases, such as Alzheimer’s disease and multiple sclerosis^121, 122^. These studies suggested that INL thickness was a response marker for inflammation during the early stages of diseases, which was further confirmed by another study, where effective disease treatment was associated with a reduction in INL thickness^123^. In addition, when we used retinal imaging traits as exposures and brain disorders as outcomes, we observed causal effects from retinal structural changes to dementia and Alzheimer’s disease. These causal links were all observed on fundus imaging traits generated from pre-trained transfer learning models. For OCT measures, causal links were identified between anxiety disorders and the thickness of the central subfield between the INL and RPE (*β* = 0.278, *P* = 7.92 × 10^-6^). Overall, MR analysis indicates that retinal imaging traits have genetic interactions with brain neurodegenerative and neuropsychiatric diseases, especially dementia and Alzheimer’s disease.

### Joint prediction of brain phenotypes using retinal and brain imaging

Using retinal and brain imaging traits, we examined whether they could be combined to better predict brain-related complex traits and diseases than using only one type of imaging data. We used a training, validation, and testing design, in which both retinal and brain images were available for the subjects in the validation and testing datasets. Model parameters were tuned based on the validation data, and prediction performance was evaluated in the independent testing dataset (**Methods**).

First, retinal imaging traits had significant prediction power on 16 brain phenotypes, including cognitive traits (such as fluid intelligence and prospective memory), neuroticism, family history of stroke, mental and behavioral disorders (ICD-10 Chapter F, such as depressive episode), and diseases of the nervous system (ICD-10 Chapter G, such as multiple sclerosis and carpal tunnel syndrome) (prediction correlation *β* range = [0.068, 0.179], *P* = [8.11×10^-19^, 7.88×10^-4^], **Fig. S83** and **Table S12**). The strongest prediction accuracy was observed on fluid intelligence (*β* = 0.179, *P* = 8.11×10^-19^). The top-ranking features for fluid intelligence prediction were from both OCT measures and fundus imaging traits, such as the thickness of RNFL, INL, and GCIPL (**Table S12**). Moreover, the prediction accuracy was improved by adding more retinal imaging traits, suggesting that various retinal structural variations captured by different retinal imaging modalities and pre-trained models can contribute to cognitive performance prediction (**Fig. S84** and **Table S12**). Multiple clinical studies have suggested that retinal imaging traits like retinal layer thickness show promising prediction power for pathological cognitive decline and dementia diagnoses^8, 102^. Similar additive effects were observed on other brain phenotypes, such as the family history of stroke (**Fig. S85**).

Next, we included brain MRI traits in the prediction model of these brain phenotypes. **Figure 6B** shows that multimodal brain imaging data can significantly predict all these brain phenotypes (*β* range = [0.091, 0.314], *P* range = [7.61×10^-6^, 3.25×10^-56^]), and using both retinal and brain imaging traits can further improve the performance (*β* range = [0.120, 0.344], *P* range = [3.72×10^-9^, 7.54×10^-68^]). For example, multiple categories of brain MRI traits can predict fluid intelligence, including DTI parameters (*β* = 0.118, *P* = 7.05 × 10^-9^), regional brain volumes (*β* = 0.132, *P* = 8.17 × 10^-11^), cortical thickness traits (*β* = 0.100, *P* = 9.01 × 10^-7^), resting fMRI (*β* = 0.216, *P* = 7.01 × 10^-27^), and task fMRI (*β* = 0.197, *P* = 2.11 × 10^-22^). Adding retinal imaging traits to each of these brain modalities improved the prediction performance over only using this single brain modality. The largest improvement was observed when we added all imaging data types together (*β* = 0.344, *P* = 7.54×10^-68^). The prediction accuracy further moved up to 0.391 (*P* = 9.56 × 10^-89^) by adding the genetic PRS of fluid intelligence (**Fig. 6C**). These results demonstrate that integrating retinal and brain imaging modalities may lead to better predictions of brain-related complex traits and diseases than using only one type of imaging data alone.

## DISCUSSION

Imaging of the eye is inexpensive and noninvasive, and it can provide rich information about the retina’s structure and function. Many brain diseases, such as neuropsychiatric and neurodegenerative disorders, are diagnosed and monitored primarily based on subjective reports of clinical symptoms^124^. The accuracy of these subjective reports is often complicated by the fact that patients with impaired mental capacity report inconsistent symptoms in varying degrees, which can bias the downstream data analysis and clinical prediction^125^. Also, patients presenting with acute mental symptomatology may have hard-to-define underlying ailments, which leads to imprecise medical management. Retinal imaging traits may serve as objective biomarkers for brain abnormalities and to assess the progression of neurological conditions^126^. In this paper, we identified novel eye-brain connections using multimodal imaging data from the two organs. The pericalcarine (primary visual cortex) and other structures within the visual pathway were associated with retinal features. Furthermore, we observed correlations between retinal features and thalamic volume, both of which are derived from the diencephalon. We then described the genetic co-architecture of the eye and the brain in 65 genomic regions, suggesting genetic associations that overlap among retinal features, brain MRI traits, and eye disorders (such as macular degeneration). We found genetic correlations and causal links between retinal imaging traits and various cognitive and mental health traits, as well as brain disorders. Additionally, we demonstrated that multi-organ images could be combined to improve the prediction of brain phenotypes. As neuroprotective treatments become more widely available, this ability to predict brain diseases could have major clinical benefits. Compared with previous clinical studies, our findings support hypotheses regarding underlying mechanisms of eye-brain connections from a novel cross-organ genetic perspective.

This study has a few limitations. Our analyses were based on the ongoing UKB brain imaging study, which currently covered only a small proportion of all UKB participants (about 10% by 2022) and consisted primarily of European ancestry individuals. We conducted phenotypic analyses on an even smaller sample of UKB subjects with both eye and brain imaging data. It is anticipated that more brain-related retinal imaging biomarkers can be discovered and replicated as the UKB brain imaging study collects data from more subjects^127^. Furthermore, it is challenging to infer phenotypic causality from our current cross-sectional analysis. Repeated UKB imaging scans in the future will allow us to study the causal relationships between eye and brain changes in a longitudinal study design. In addition, the eye-brain genetic links identified were European or UKB specific, and it will be important to examine whether these cross-organ genetic overlaps can be generalized to other populations or studies when more data are collected^128^. In summary, the massive genetic connections between the brain and the eye found in our UKB-based study support the use of retinal imaging to study and manage the risk of brain disorders.

The utility of these retinal imaging biomarkers needs to be verified in future clinical and research settings.

## Data Availability

GWAS summary statistics of brain MRI traits can be freely downloaded at BIG-KP (https://bigkp.org/). GWAS summary statistics of retinal imaging traits will be made publicly available at Eye-KP (https://www.eyekp.org/). The individual-level data used in this study can be obtained from https://www.ukbiobank.ac.uk/.

https://www.eyekp.org/

## METHODS

Methods are available in the ***Methods*** section.

*Note: One supplementary information pdf file and one supplementary table zip file are available*.

## ACKNOWLEDGEMENTS

We thank Mufeng Gao for her help with data management in the early stage of this project. The study has been partially supported by start-up funds from Purdue Statistics Department and funding from Analytics at Wharton. This research has been conducted using the UK Biobank resource (application number 22783), subject to a data transfer agreement. We would like to thank the individuals who represented themselves in the UK Biobank for their participation and the research teams for their efforts in collecting, processing, and disseminating these datasets. We would like to thank the research computing groups at the University of North Carolina at Chapel Hill, Purdue University, and the Wharton School of the University of Pennsylvania for providing computational resources and support that have contributed to these research results. We gratefully acknowledge all the studies and databases that made GWAS summary-level data publicly available. Y.L. is partially supported by R56 AG079291 and U01 HG011720.

## AUTHOR CONTRIBUTIONS

B.Z. designed the study. Y.J.L. and B.Z. processed the raw retinal imaging data. B.Z., Z.F., Z.W., J.S., X.Y., X.F.W., B.L., X.Y.W, and C.C. analyzed the data. Z.F., B.Z., Y.Y., and J.L. designed the website and developed online resources. Y.L.Y, Y.L., J.L.S., J.M.O., T.L., and H.Z. provided comments and helped interpret the results. B.Z. wrote the manuscript with feedback from all authors.

## CORRESPONDENCE AND REQUESTS FOR MATERIALS

should be addressed to B.Z and H.Z.

## Code availability

We made use of publicly available software and tools. The original codes to apply pre-trained transfer learning models to extract features from raw retinal fundus images are available at https://github.com/mkirchler/transferGWAS. Our code with more implemented pre-trained CNN models will be shared on Zenodo https://zenodo.org/.

## METHODS

### Eye and brain imaging data

Our study was based on data obtained from the UK Biobank (UKB) study, which recruited approximately half a million individuals between the ages of 40 and 69 between 2006 and 2010^42^ (https://www.ukbiobank.ac.uk/). The ethics approval of the UKB study was from the North West Multicentre Research Ethics Committee (approval number: 11/NW/0382). The optical coherence tomography (OCT) and retinal imaging scans were part of the eye measurements conducted during the participant’s visit to the UKB assessment center. We considered two sets of retinal imaging traits. First, we used the derived OCT measures in Category 100079 (https://biobank.ndph.ox.ac.uk/showcase/label.cgi?id=100079), which were generated and returned by previous studies^29, 43, 44^. These measures mainly provide the thickness of different retinal layers and their subfields, as well as the vertical cup-to-disc ratio and disc diameter. As suggested, we used the data in Data-Fields 28552 & 28553^43^ to perform quality control for these OCT measures by keeping images with an image quality score > 45. We further only keep the OCT measures with a sample size > 30,000, resulting in 46 measures with an average sample size of 62,425.

Second, we downloaded the raw fundus retinal eye images from Category 100016 (https://biobank.ndph.ox.ac.uk/showcase/label.cgi?id=100016) and performed GWAS on these whole images by extracting imaging biomarkers using transfer learning models. Briefly, we used multiple pre-trained deep convolutional neural networks (CNNs) trained from the ImageNet^45^ database. The ImageNet database contains more than 14 million images classified into more than 20,000 classes, which can be used to train models that extract various features from retinal fundus images. Many CNNs models have been trained on ImageNet and were widely used in the image processing field to learn complex patterns from images. In addition to the ResNet50^129^ model used by the transferGWAS^31^, we implemented 10 more pre-trained CNN models, including the AlexNet^130^, Vgg16^131^, Vgg19^131^, GoogLeNet (Inception V1)^132^, Inception (V3)^132^, ResNet18^129^, ResNet34^129^, SqueezeNet^133^, MobileNet^134^, and ShuffleNet^135^. These pre-trained models are available on Pytorch^136^ and represent different designs and architectures, such as layer depth, size of kernels, and hyperparameters. For example, the ResNet50 has 50 layers with kernel size 1 × 1, 3 × 3, and 7 × 7, while AlexNet has 8 layers with kernel size 3 × 3, 5 × 5, and 11 × 11. All pre-trained models use the rectified linear unit (ReLU) as the activation function. We began by combining the original left and right retinal fundus images and the rotated images with 90°, 180°, and 270°, each with and without horizontal mirroring. Next, we input these eight retinal fundus images into each pre-trained model and averaged the outputs from the last layer of convolutional networks. Then we generated the top-10 ranked principal components (PCs) from each of the 11 models as retinal imaging biomarkers in downstream GWAS analyses. The average sample size across all these 110 (10 × 11) fundus imaging traits is 78,513. In all the OCT measures and fundus image traits, the values greater than five times the median absolute deviation from the median were treated as outliers and removed.

The UKB brain MRI data were generated from raw images downloaded from Category 100003 (https://biobank.ndph.ox.ac.uk/showcase/label.cgi?id=100003). The multimodal brain imaging traits used in the present paper have been extracted in previous papers by our research group^35, 37, 39, 46^. First, we had 101 regional brain volumes^35^ and 63 cortical thickness traits^46^ generated from T1-weighted structural MRI images. These structural MRI traits were produced by the advanced normalization tools^137^ (ANTs). For the 101 volumetric traits, we had brain volumes for 98 pre-defined cortical and subcortical areas and three global brain volume measures (total gray matter volume, total white matter volume, and total brain volume). We also examined the thickness of 62 cortical areas and the global thickness. Second, the ENIGMA-DTI pipeline^138, 139^ was used to generate 110 tract-averaged DTI parameters based on diffusion MRI, including fractional anisotropy, mean diffusivity, axial diffusivity, radial diffusivity, and mode of anisotropy, for 21 predefined major white matter tracts and the whole brain (5 × 22). For resting fMRI, we applied the Glasser360 atlas^140^ to partition the cerebral cortex into 360 regions for 12 functional networks^141^, including the primary visual, secondary visual, auditory, somatomotor, cingulo-opercular, default mode, dorsal attention, frontoparietal, language, posterior multimodal, ventral multimodal, and orbito-affective networks. We generated 92 functional activity (amplitude) and functional connectivity traits, including the average activity for each network and the average connectivity for each pair of networks (including within the same network), as well as the global activity and connectivity of the whole cortex. Similarly, 92 functional activity and connectivity traits were generated from task fMRI^39^. In summary, we considered 458 brain MRI traits of brain structure and function. See **Table S1** for the complete ID list of both retinal imaging and brain imaging traits.

### Phenotypic eye-brain imaging analyses

In our phenotypic analysis, we examined pairwise associations between 156 retinal imaging traits and 458 brain MRI traits. We used the UKB subjects with both two imaging types and adjusted a wide range of covariates, including age, sex, standing height, assessment center, body mass index, weight, waist-to-hip ratio, smoking status, mean arterial blood pressure, age-squared, age-sex-interaction, age-squared-sex-interaction, top 40 genetic PCs^142^, volumetric scaling, head motion, head motion-squared, brain position, brain position-squared^34, 36^, diabetes, ICD-10 disease code staring with R73 (“elevated blood glucose level”, such as hyperglycemia), I70 (“atherosclerosis”, such as atherosclerosis of aorta), I10 (Essential (primary) hypertension), and E78 (“disorders of lipoprotein metabolism and other lipidaemias”, such as hyperlipidemia). For regional brain volumes, we additionally corrected for total brain volume to remove global effects. We fitted linear models for each pair of imaging traits (R version 3.6.0) and used a discovery-validation design, in which the UKB individuals of white British ancestry (average *n* = 6,454 across different modalities) were used to discover eye-brain imaging associations, which were verified by a hold-out independent validation dataset (average *n* = 959, relatives^142^ of the discovery sample were removed). The Benjamini-Hochberg procedure was used to adjust for multiple testing, and we reported significant associations at the false discovery rate (FDR) of 5%. Validation criteria included a *P* value less than 0.05 in the hold-out independent dataset with concordant association signs between the discovery and validation datasets. We also considered the conservative Bonferroni multiple testing correction and highlighted these top-ranking significant findings in the paper. In addition, we repeated the above analysis separately for females and males (average *n* = 3,338 and 3,150, respectively) and reported the sex-specific association patterns.

### Genetic analysis of 156 retinal imaging traits

We performed GWAS for the 156 retinal imaging traits using the imputed genotyping data from the UKB study. For the set of subjects with both retinal imaging traits and genetic data, we performed the following quality controls^39^: 1) removed individuals with missing genotype rate > 0.1; 2) removed variants with missing genotype rate > 0.1; 3) removed variants with minor allele frequency (MAF) < 0.01; 4) removed variants that failed the Hardy-Weinberg equilibrium test at 1 × 10^-7^ level; and 5) removed variants with imputation INFO score < 0.8. The SNP-based heritability of white British samples was estimated based on all autosomal SNPs using GCTA^50^ (average *n* = 60,748). We adjusted for the effects of age, sex, assessment center, age-squared, age-sex interaction, age-squared-sex interaction, and top 40 genetic PCs^142^.

Using the same set of subjects and covariates data, we performed GWAS using linear mixed effect models via fastGWA^143^. SNP heritability and GWAS were also conducted separately for males and females. We defined the independent (LD *r*^2^ < 0.1) significant genetic associations and loci using FUMA^144^ (version v1.3.8). The details of FUMA annotations can be found at https://fuma.ctglab.nl/tutorial. Briefly, FUMA identified variants whose *P* values passed our stringent GWAS significance level 3.20 × 10^-10^ (the standard GWAS significance level after further Bonferroni-adjusted for the 156 retinal imaging traits) and were independent of other significant variants (LD *r*^2^ < 0.1). Based on these independently significant variants, FUMA constructed LD blocks by considering all variants (MAF ≥ 0.0005, including variants from the 1000 Genomes reference panel) in LD (*r*^2^ ≥ 0.6) with at least one independent significant variant. For independently significant associations defined by FUMA, we performed validations using 1) the UKB European but non-British subjects (average *n* = 5,320) and 2) UKB non-European subjects (average *n* = 6,490). Relatives of the discovery GWAS sample were removed, and we adjusted for the top 10 genetic PCs instead of the top 40. Other adjusted covariates remained the same. We also developed polygenic risk scores (PRS) using summary statistics from discovery GWAS and examined their prediction accuracy on the two validation datasets. We constructed PRS based on PRS-CS^53^ with all default parameters. The validation genotype data were randomly selected from 1,500 UKB European subjects without retinal imaging data.

### Genetic eye-brain imaging analyses

For the independently significant variants and all variants in their LD blocks, we used FUMA to look them up on the NHGRI-EBI GWAS catalog (version e104_2021-09-15) to search for any previously GWAS results reported on these variants (*P* < 9 × 10^-6^). We focused on the existing GWAS results of brain and eye-related complex traits and diseases and manually categorized them into 14 groups, including stroke (and other cerebrovascular disorders, such as Moyamoya disease, intracranial aneurysm, and cerebral aneurysm), Parkinson’s disease, Alzheimer’s disease, glioma/glioblastoma (GBM), other neurological disorders (such as amyotrophic lateral sclerosis, progressive supranuclear palsy, corticobasal degeneration, and frontotemporal dementia), schizophrenia, other psychiatric disorders (such as bipolar disorder, depression, major depressive disorder, and autism spectrum disorder), psychological traits (such as neuroticism, anxiety, subjective well-being, and risk tolerance), cognitive traits (such as general cognitive ability, the highest math class taken, intelligence, and reaction time), education, brain structure/function, migraine, Alzheimer’s disease biomarkers (such as cerebrospinal fluid biomarker levels, rate of cognitive decline in Alzheimer’s disease, and plasma t-tau levels), and eye traits/diseases (such as macular thickness, refractive error, spherical equivalent, and glaucoma). In addition, we systematically examined genetic overlaps with the GWAS results of brain MRI traits reported in previous studies, including 101 regional brain volumes^35^, 215 DTI parameters^37^ (including the 110 tract-average values used in our phenotypic analysis and 105 additional PCs of fractional anisotropy), 63 cortical thickness traits^46^, 92 resting fMRI traits, and 92 task fMRI traits^39^. For the index variants of retinal imaging traits defined by FUMA, we looked up the MetaBrain database^61^ (https://www.metabrain.nl/) to see if they were reported eQTLs in large-scale gene expression meta-analysis of brain tissues. For each locus with shared genetic influences, we tested for common causal genetic variants between the retinal imaging trait and the brain phenotype using Bayesian colocalization analysis^59^. The colocalization was established if the posterior probability of the shared causal variant hypothesis (PPH4) was greater than 0.8^59, 60^.

Cross-trait LDSC^98^ (https://github.com/bulik/ldsc/, version 1.0.1) was used to examine the pairwise genetic correlation between 156 retinal imaging traits and 39 sets of publicly available GWAS summary statistics of brain phenotypes. The default European LD scores provided by the LDSC software were used, which were based on the 1000 Genomes European data. We used the HapMap3 variants, and variants in the major histocompatibility complex region were excluded. For the 46 OCT measures, we also used LDSC to perform the heritability enrichment analysis^106^ with genetic variant annotations of tissue type and cell type-specific regulatory elements. The heritability explained by the annotated genome regions was estimated and tested with percentages and enrichment scores. Baseline annotation models were included in the analysis when we analyzed additional annotations. We tested for the annotations of regulatory elements from multiple adult and fetal tissues from the Roadmap Epigenomics Consortium^107^ and two major brain cell types (neurons and glia) sampled from various brain cortical and subcortical brain regions^108^.

Bi-directional Mendelian randomization (MR) analysis was used to discover the causal effect between 156 retinal imaging traits and 25 brain-related clinical endpoints. Eight MR methods^110–117^ were implemented, including MR Egger, simple median, simple mode, fixed effect inverse variance weighted (IVW), multiplicative random effect IVW, DIVW, MR-RAPS, and GRAPPLE. The 25 brain-related clinical endpoints were all from the latest release (R7) of FinnGen database (https://www.finngen.fi/en/access_results), where 12 of them were mental and behavior disorders, and the remaining 13 phenotypes were diseases of the nervous system. Most of the diseases we selected have a number of cases greater than 10,000, except for a few important brain diseases, including Alzheimer’s disease (*n* > 6,000), other neurological diseases (*n* = 7288), and epilepsy (*n* = 8523). **Table S11** provides more information on the MR methods and FinnGen data. Exposure GWAS summary statistics were first clumped with Plink^145^ to guarantee that the instrumental variables used in MR models are independent. The *P* value significance threshold (p1) and the secondary significance threshold (p2) in clumping were set to 5 × 10^-8^, and the 1000 Genomes European reference panel was applied. Besides, the threshold over the squared correlation between two genetic variants was set to be *r*^2^ = 0.01 and window size = 1Mb. After clumping, the selected SNPs from exposure GWAS data were extracted from outcome GWAS summary statistics with function *extract_outcome_data()* in the two-sample MR package (https://mrcieu.github.io/TwoSampleMR/). To ensure that the effect of a genetic variant on the exposure and outcome corresponded to the same allele, data harmonization was performed using the *harmonise_data function()* with the default settings. The estimated causal pairs of retinal imaging trait and brain disease were further screened with several rules. The first step was to discard pairs with fewer than six genetic variants. Second, we dropped the pairs whose estimated MR Egger intercept differed significantly from zero^118^. Bonferroni correction was then performed on the MR results of each method separately. Finally, we reported the causal pairs that were significant for either of the IVW methods and at least one of the robust MR methods (DIVW, simple mode, simple median, MR-RAPS, and GRAPPLE).

### Prediction of brain phenotypes using retinal and brain imaging data

We examined the prediction power of 156 retinal imaging traits on 32 brain-related complex traits and diseases, including cognitive traits, neuroticism sum score, family history of brain disorders, mental and behavioral disorders (ICD-10 Chapter F), and diseases of the nervous system (ICD-10 Chapter G). We focused on unrelated white British subjects and randomly selected 50,944 subjects as the training dataset, 2,464 subjects as the validation dataset, and 2,464 subjects as our testing dataset. The subjects in the validation and testing datasets also had brain MRI data, enabling testing the prediction performance with both two imaging types in later steps. For each of the 32 traits, we used ridge regression for prediction and the effect sizes of retinal imaging traits were estimated on the training dataset via the glmnet^146^ package (R version 3.6.0). All model parameters were tuned based on validation data, and prediction performance was examined based on the correlation between the predicted values and the observed ones in the independent testing data. In all the training, validation, and testing datasets, we removed the effects of age, sex, age-sex interaction, age-squared, age-squared-sex interaction, assessment center, and top 40 genetic PCs. For brain phenotypes where retinal imaging traits had significant predictive power after Bonferroni correction for multiple testing, we further examined the predictive power of multiple brain MRI modalities and the joint performance of using retinal and brain imaging traits. In brain imaging prediction, we used the same validation and testing datasets as the retinal imaging analysis and all other unrelated white British subjects (average *n* = 37,239) as training data. Finally, we examined the prediction accuracy of genetic PRS for fluid intelligence. We used the unrelated white British subjects without retinal or brain imaging data as training GWAS (*n* = 71,406) and developed the PRS with PRS-CS^53^. The same set of covariates as the imaging prediction analysis was removed.

