## Supplementary material for "Eye-brain connections revealed by multimodal retinal and brain imaging genetics in the UK Biobank": supp_figures

**This PDF file includes:**

Figs. S1 to S85
Legends for Tables S1 to S12

**Other Supplementary Materials for this manuscript include the following:**

Tables S1 to S12 (.xlsx) (available in a zip file)

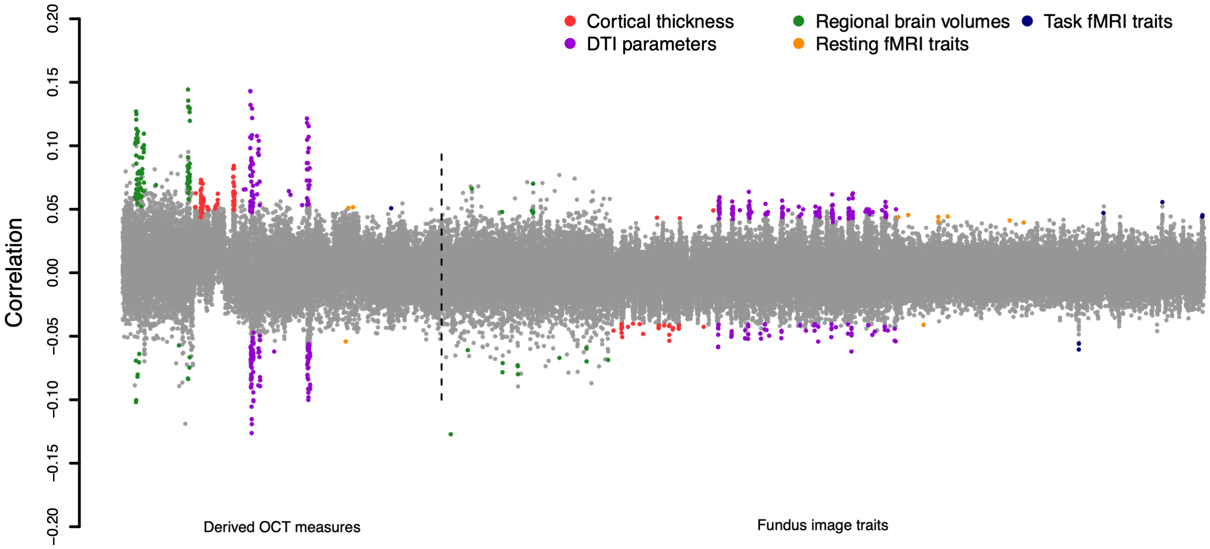


**Fig. S1 Phenotypic eye-brain associations.** The correlation coefficients between 156 retinal imaging traits (46 derived OCT measures and 110 fundus image traits) and 5 groups of brain MRI traits, including 101 regional brain volumes, 63 cortical thickness traits, 110 DTI parameters, 92 resting fMRI traits, and 92 task fMRI traits. The coefficients whose *P* values were significant after controlling for multiple testing at the false discovery rate (FDR) level of 5% level ($P < 4.37 \times 10^{-4}$) are highlighted in colors. We label the categories of brain imaging traits with different colors.

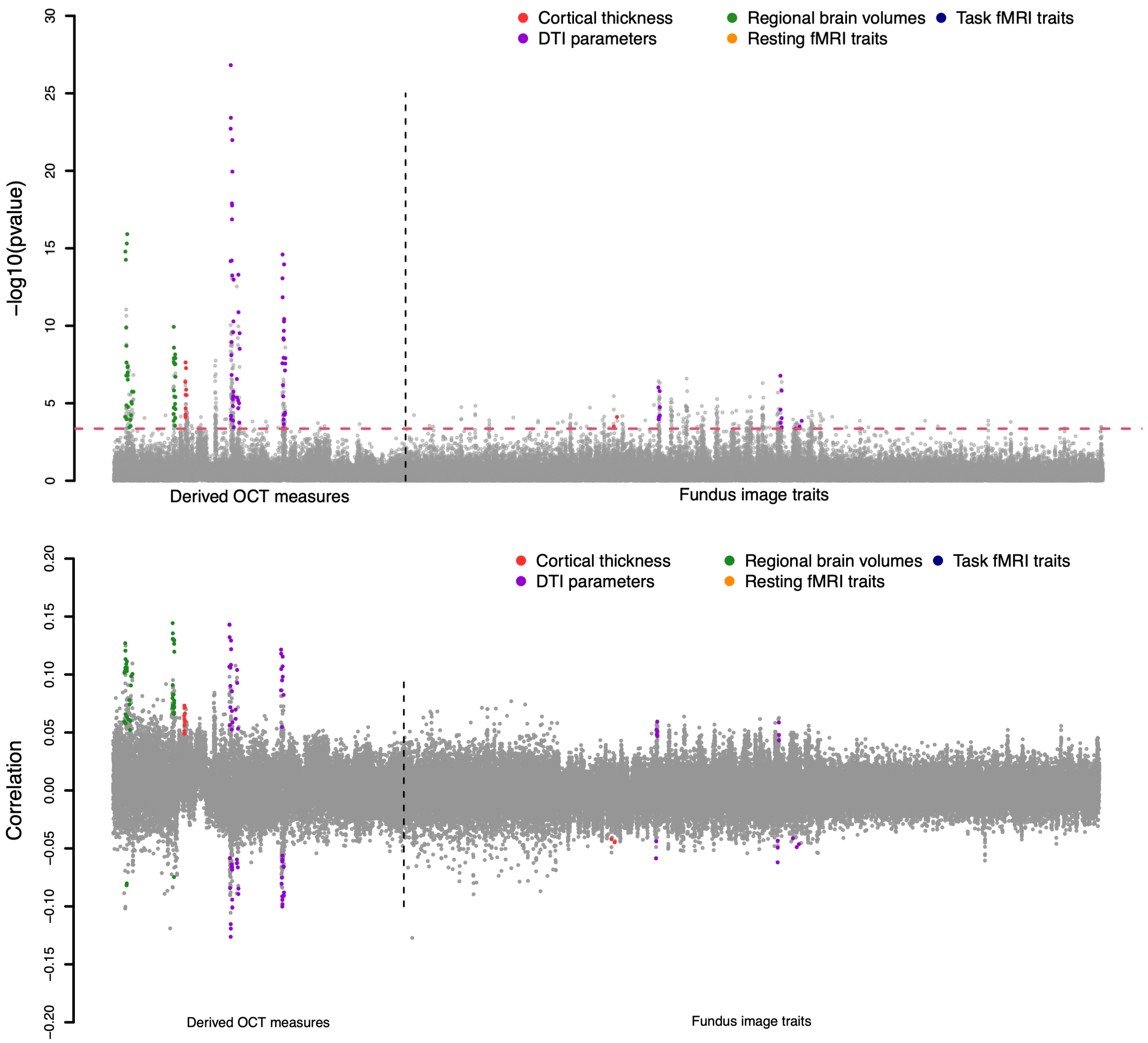


**Fig. S2 Phenotypic eye-brain associations that can be validated in an independent hold-out sample.** The *P* values (upper panel) and correlation coefficients (lower panel) between 156 retinal imaging traits (46 derived OCT measures and 110 fundus image traits) and 5 groups of brain MRI traits, including 101 regional brain volumes, 63 cortical thickness traits, 110 DTI parameters, 92 resting fMRI traits, and 92 task fMRI traits. The FDR-significance level ($P < 4.37 \times 10^{-4}$, horizontal line) coefficients validated in the independent hold-out sample are highlighted in colors. We label the categories of brain imaging traits with different colors.

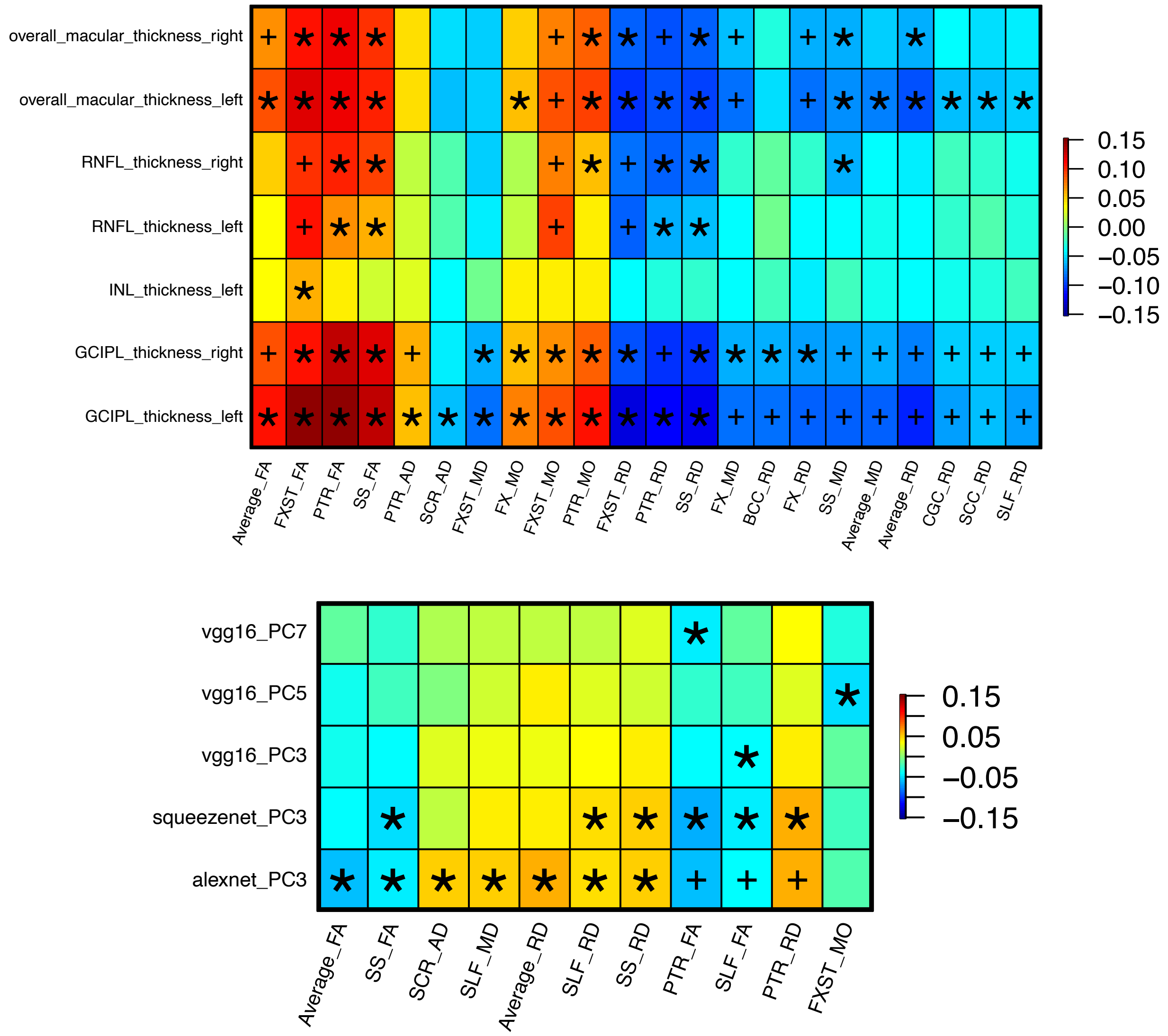


**Fig. S3 Phenotypic associations between retinal imaging traits and DTI parameters.** We illustrate correlations between retinal imaging traits (*y*-axis) and DTI parameters (*x*-axis). The color represents correlation estimates. The plus signs indicate significant correlations at the FDR-significance level ($P < 4.37 \times 10^{-4}$), and the asterisks highlight those were further validated in an independent hold-out sample.

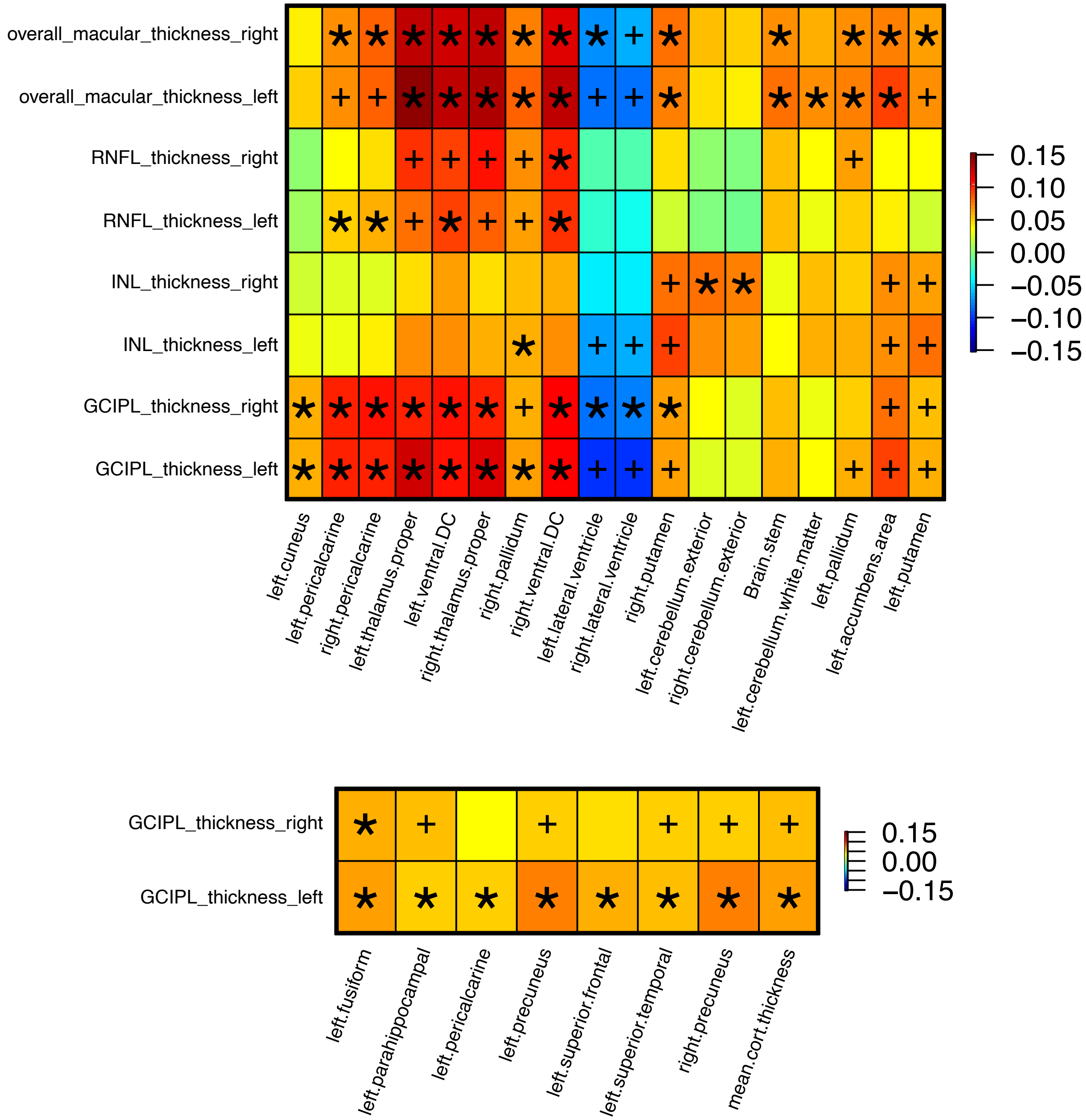


**Fig. S4 Phenotypic associations between derived OCT measures and brain structural MRI traits.** We illustrate correlations between derived OCT measures (*y*-axis) and regional brain volumes (*x*-axis, upper panel) or cortical thickness traits (*x*-axis, lower panel). The color represents correlation estimates. The plus signs indicate significant correlations at the FDR-significance level ($P < 4.37 \times 10^{-4}$), and the asterisks highlight those were further validated in an independent hold-out sample.

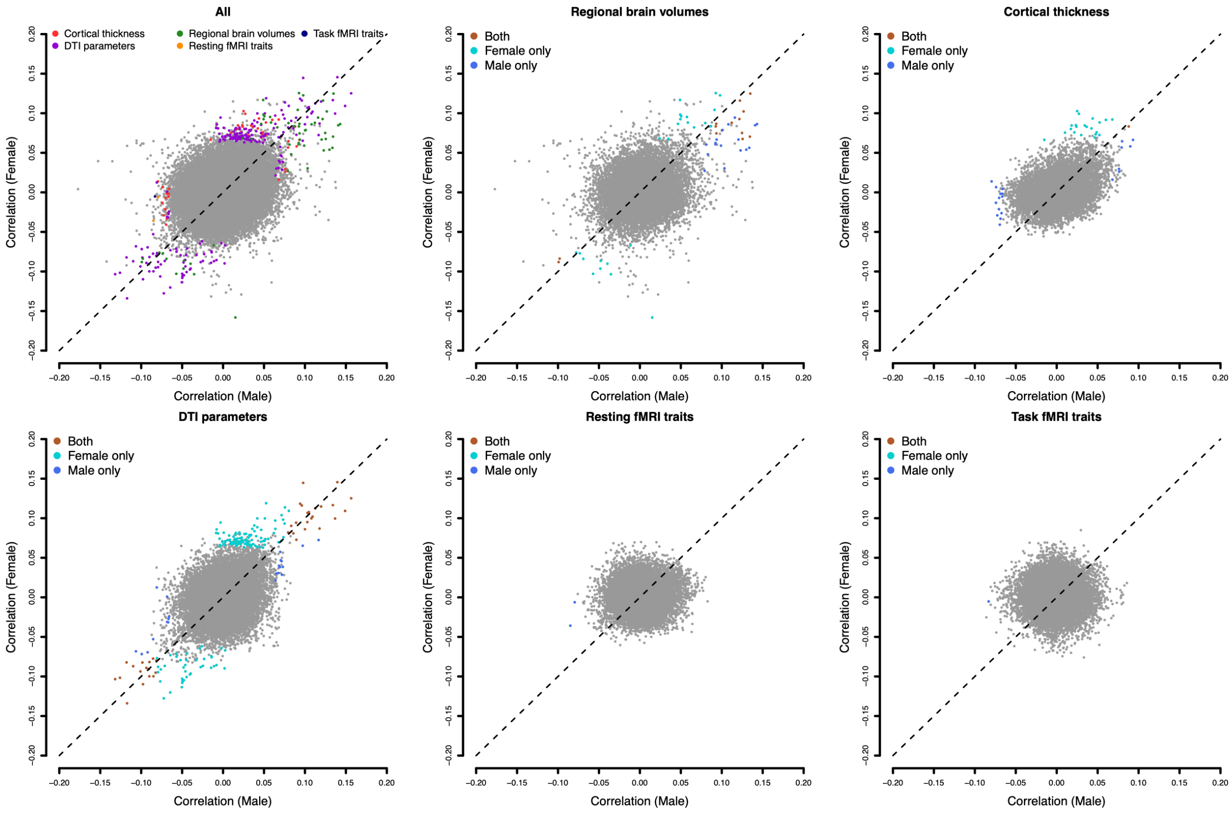


**Fig. S5 Comparison of phenotypic eye-brain associations in sex-specific analysis.** The correlation coefficients between retinal imaging traits and brain MRI traits were separately estimated for males (*x*-axis) and females (*y*-axis). There are 5 groups of brain MRI traits, including 101 regional brain volumes, 63 cortical thickness traits, 110 DTI parameters, 92 resting fMRI traits, and 92 task fMRI traits. The coefficients whose *P* values pass the FDR-significance level ($P < 4.37 \times 10^{-4}$) in either male or female analyses are highlighted in colors.

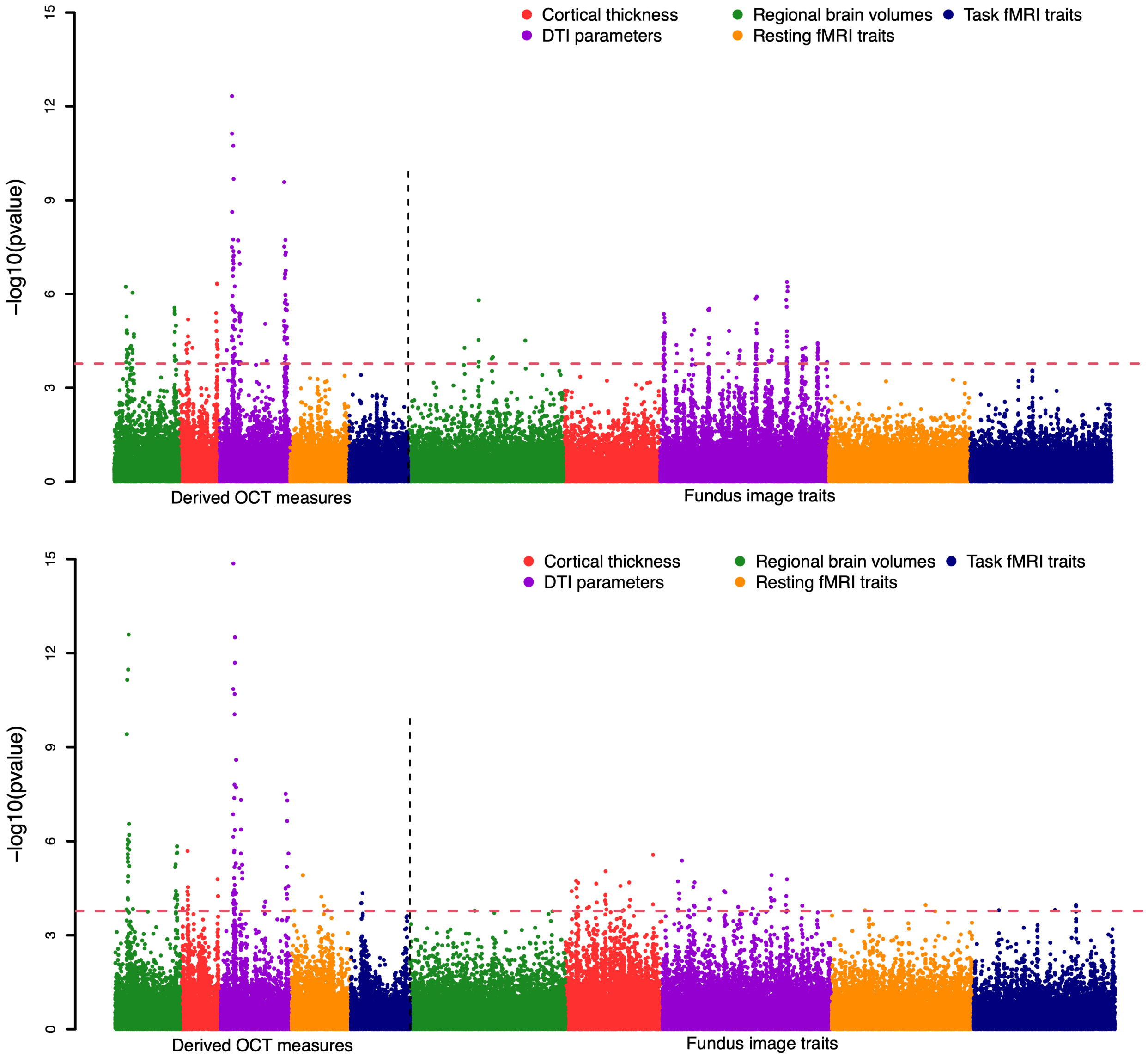


**Fig. S6 Phenotypic eye-brain associations in sex-specific analyses.** The -log10(p-value) between retinal imaging traits and brain MRI traits are separately displayed for females (upper panel) and males (lower panel). There are 5 groups of brain MRI traits, including 101 regional brain volumes, 63 cortical thickness traits, 110 DTI parameters, 92 resting fMRI traits, and 92 task fMRI traits. The dashed lines indicate the FDR-significance level ($P < 4.37 \times 10^{-4}$). We label the categories of brain imaging traits with different colors.

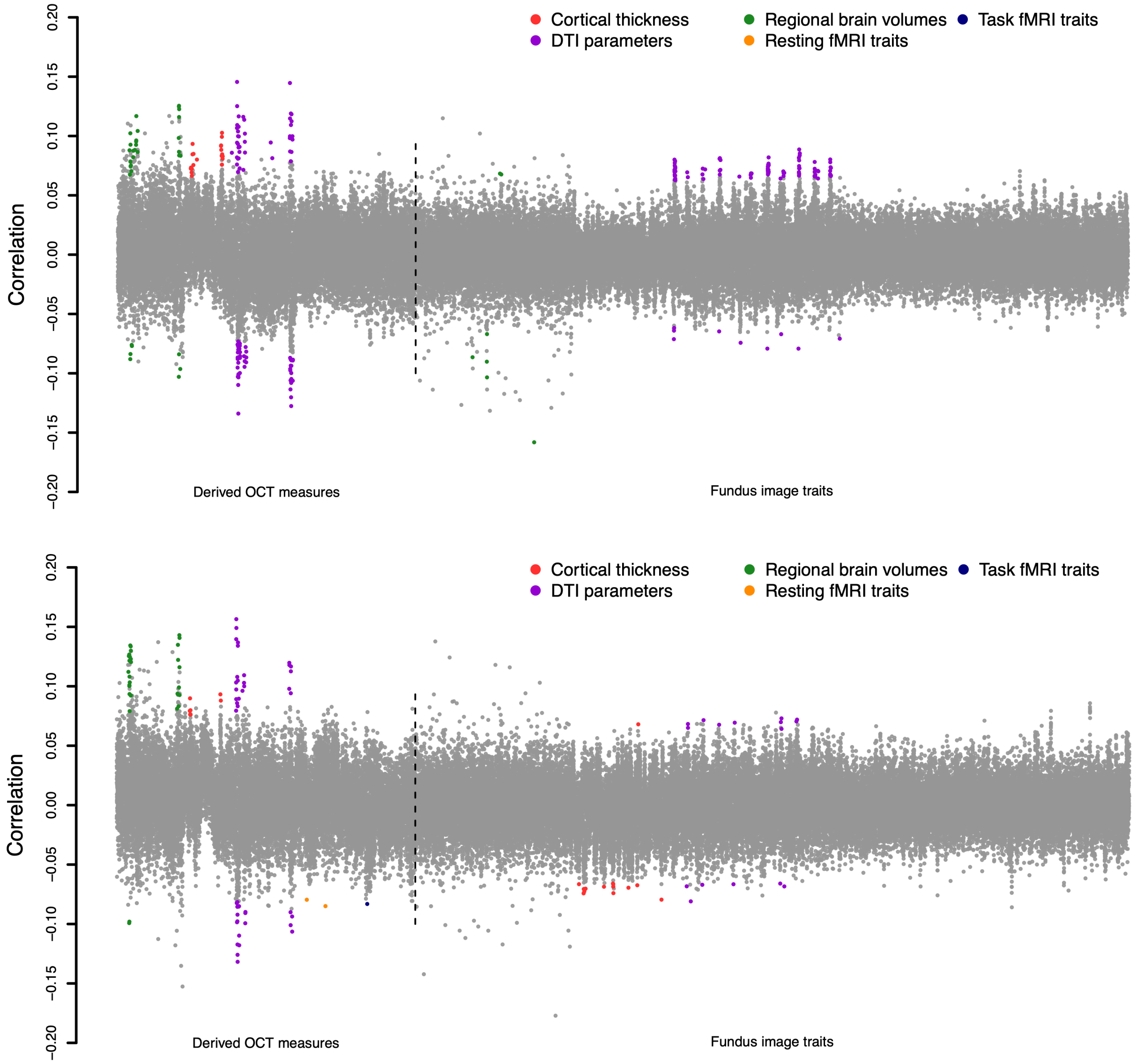


**Fig. S7 Phenotypic eye-brain associations in sex-specific analyses.** The correlation coefficients between retinal imaging traits and brain MRI traits are separately displayed for females (upper panel) and males (lower panel). There are 5 groups of brain MRI traits, including 101 regional brain volumes, 63 cortical thickness traits, 110 DTI parameters, 92 resting fMRI traits, and 92 task fMRI traits. The coefficients whose *P* values passing the FDR-significance level ($P < 4.37 \times 10^{-4}$) are highlighted in colors. We label the categories of brain imaging traits with different colors.

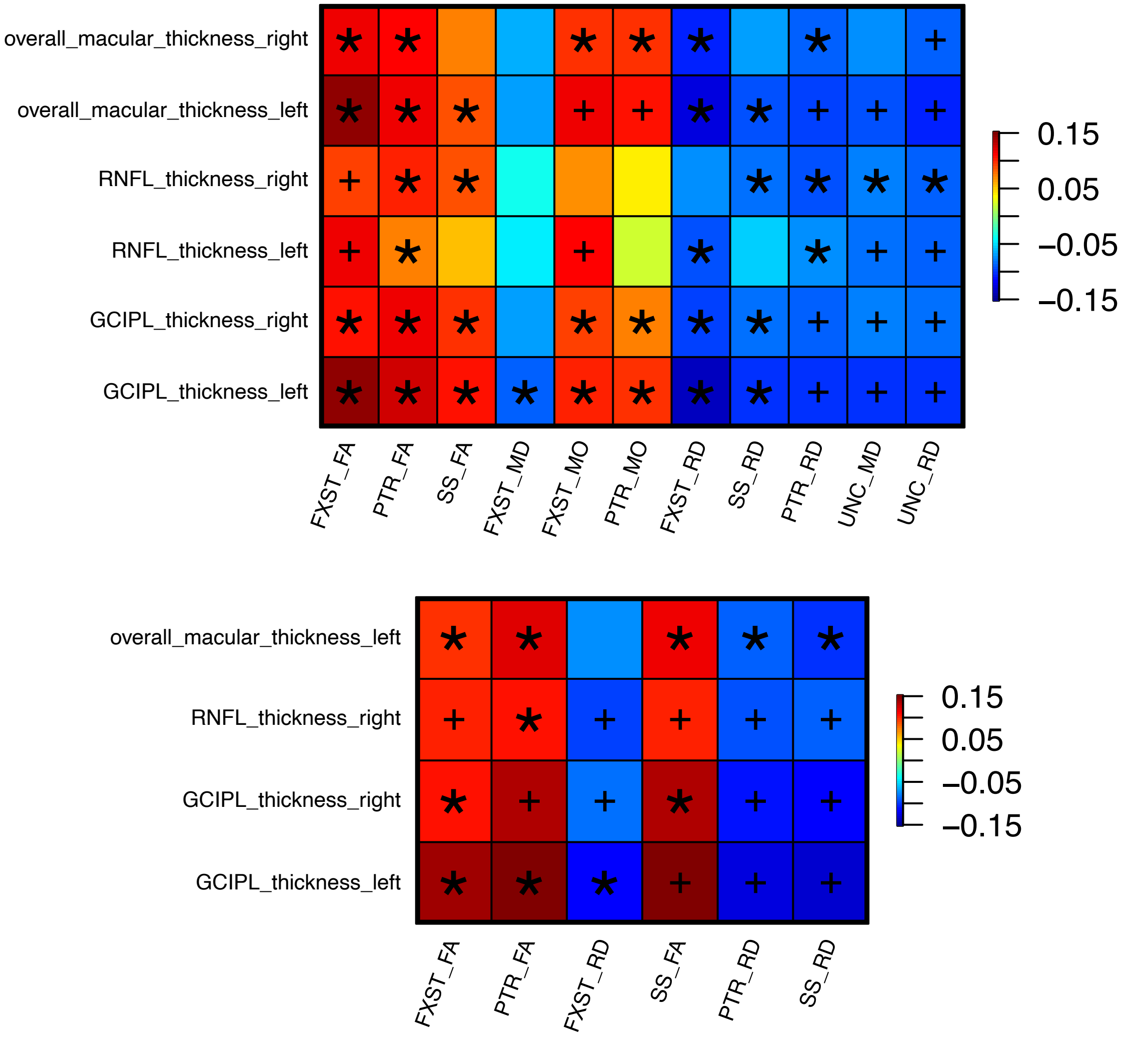


**Fig. S8 Phenotypic associations between retinal imaging traits and DTI parameters in sex-specific analyses.** We illustrate correlations between retinal imaging traits (*y*-axis) and DTI parameters (*x*-axis) for females (upper panel) and males (lower panel). The color represents correlation estimates. The plus signs indicate significant correlations at the FDR-significance level ($P < 4.37 \times 10^{-4}$), and the asterisks highlight those were further validated in an independent hold-out sample.

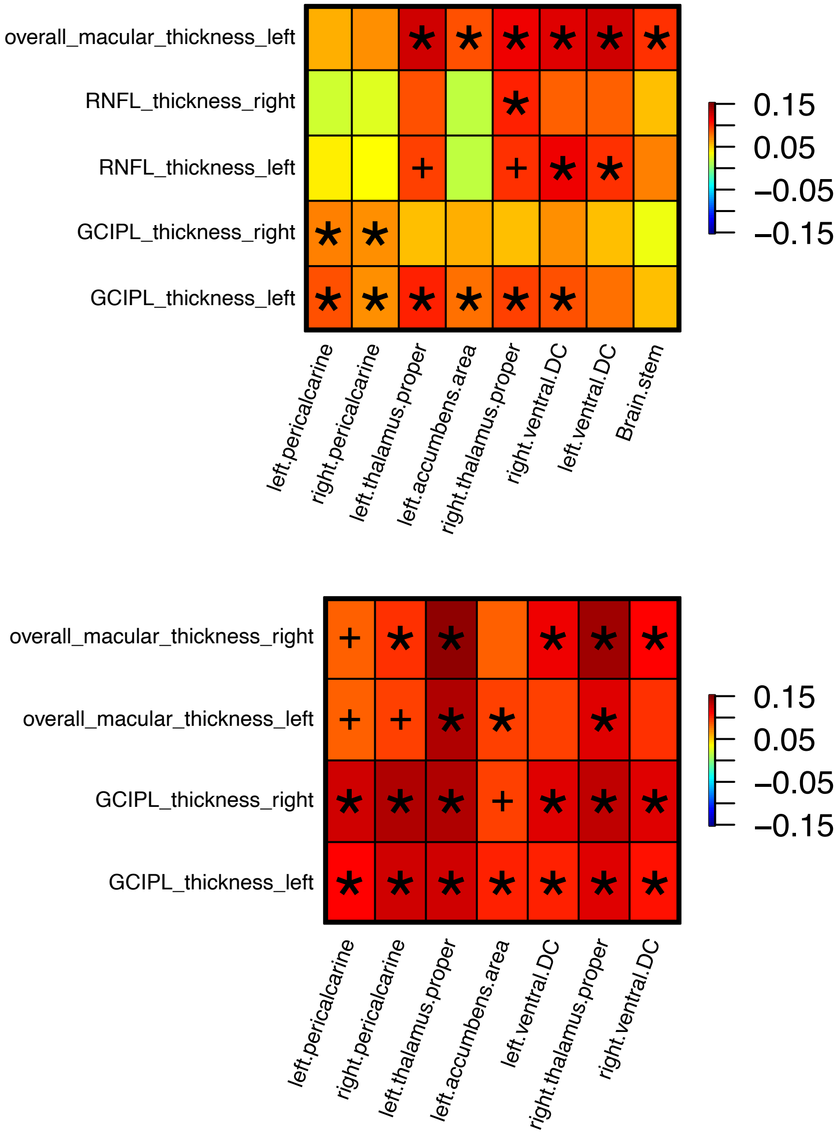


**Fig. S9 Phenotypic associations between retinal imaging traits and regional brain volumes in sex-specific analyses.** We illustrate correlations between retinal imaging traits (*y*-axis) and DTI parameters (*x*-axis) for females (upper panel) and males (lower panel). The color represents correlation estimates. The plus signs indicate significant correlations at the FDR-significance level ($P < 4.37 \times 10^{-4}$), and the asterisks highlight those were further validated in an independent hold-out sample.

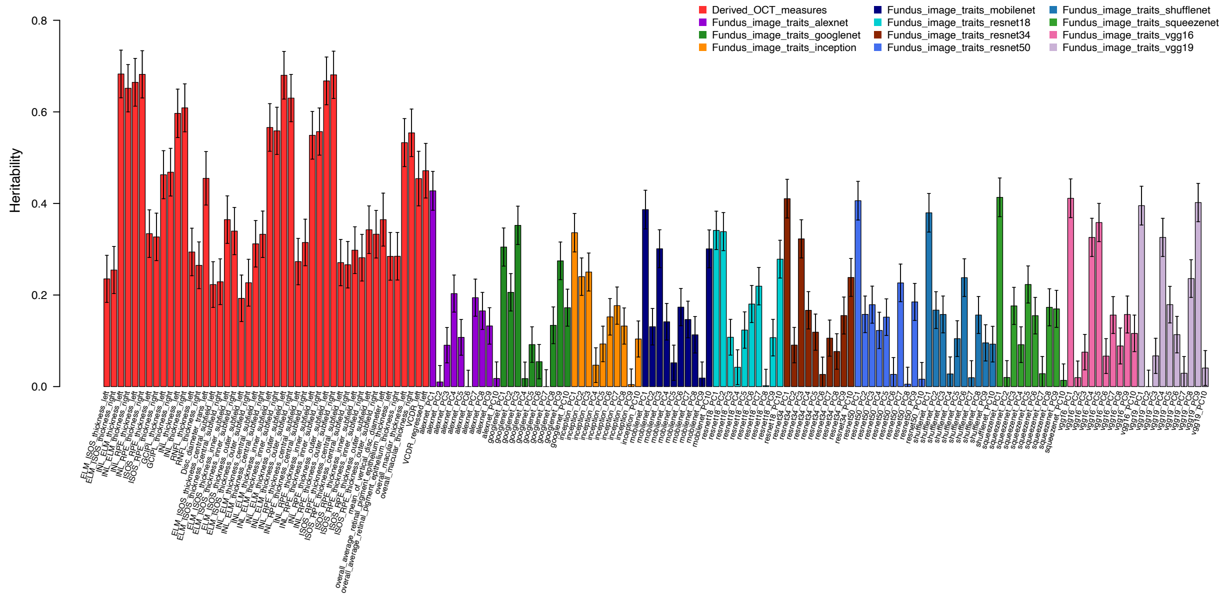


**Fig. S10 SNP heritability of the 156 retinal imaging traits.** The *x*-axis displays the ID of retinal imaging traits, see Table S1 for more information on these traits.

5

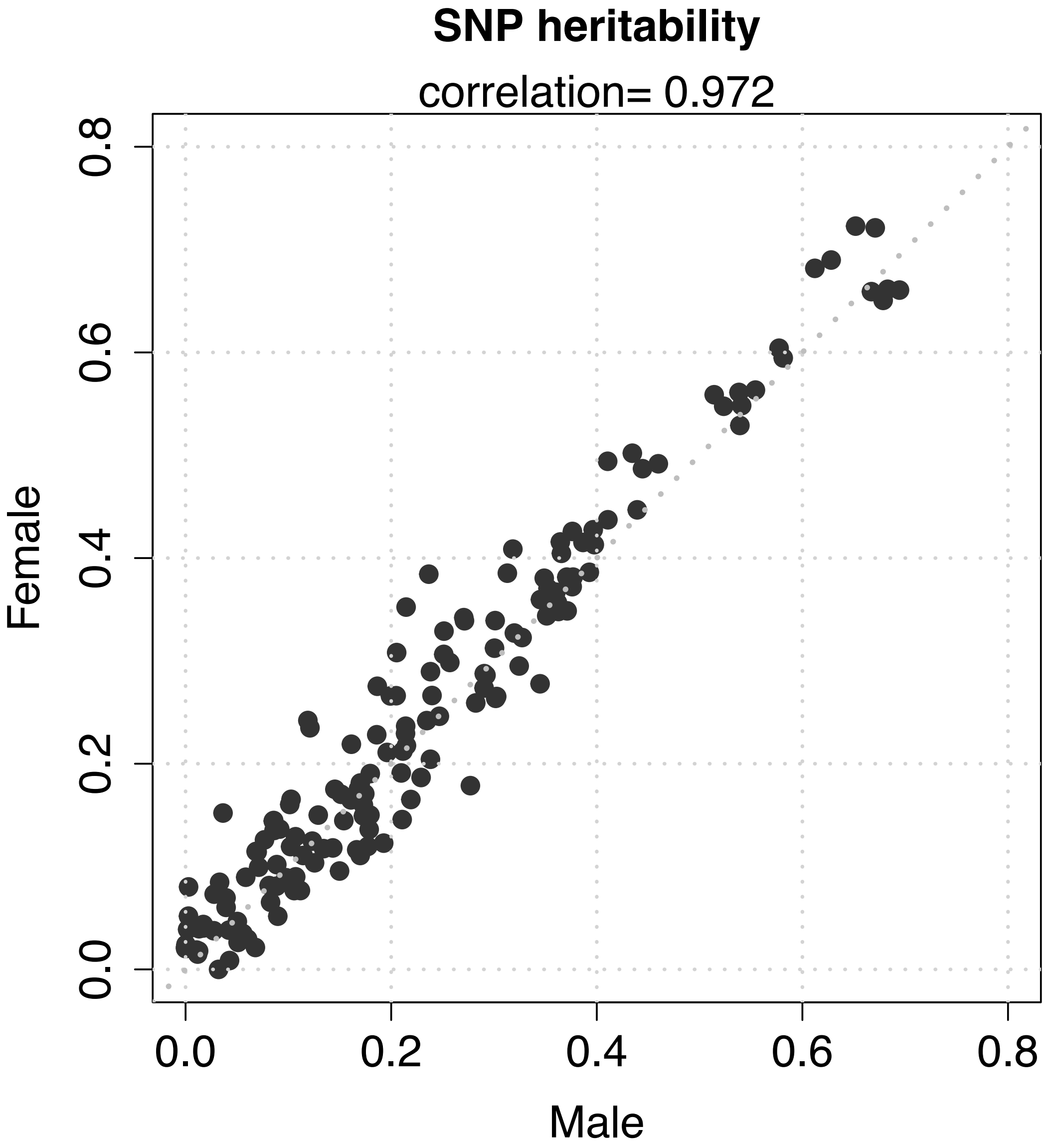


**Fig. S11 Comparison of heritability estimates of in retinal imaging traits sex-specific analysis.** The heritability was separately estimated for males (*x*-axis) and females (*y*-axis).

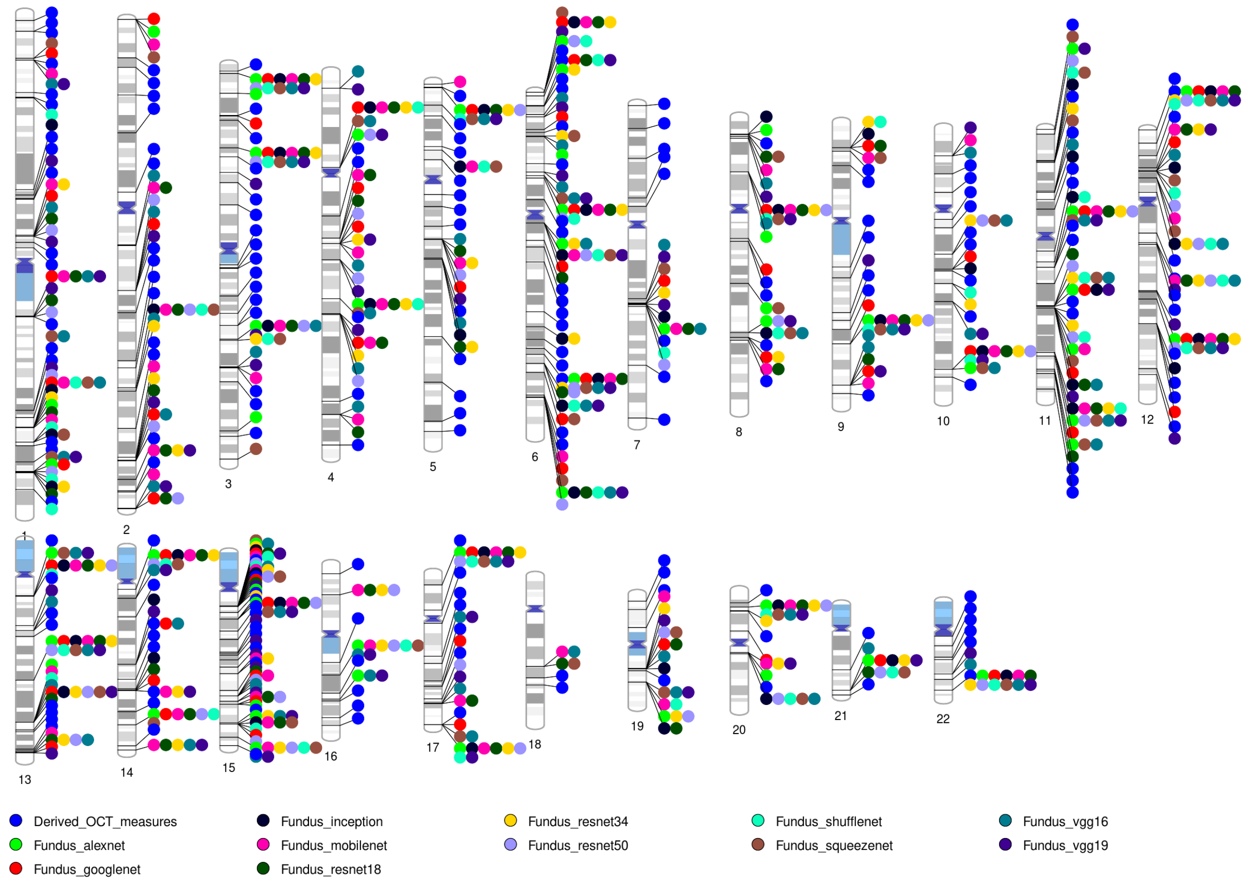


**Fig. S12 Ideogram of the 258 genomic regions associated with retinal imaging traits ($P < 3.20 \times 10^{-10}$).** We label the categories of retinal imaging traits with different colors.

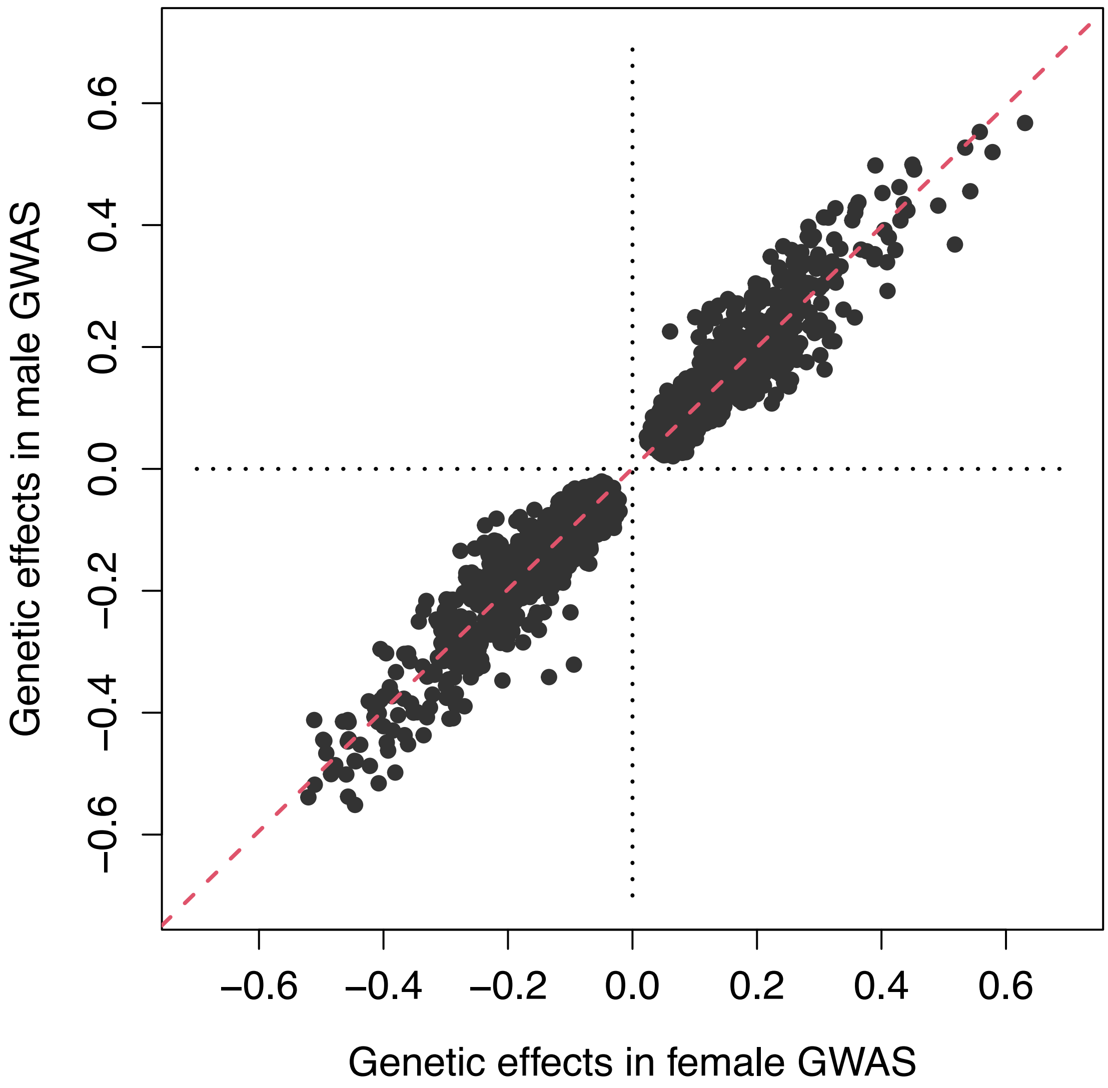


**Fig. S13 Genetic effects in sex-specific GWAS.** GWAS was conducted separately for female and male subjects, and we illustrate the significant genetic effects identified in our discovery GWAS ($P < 3.20 \times 10^{-10}$).

5

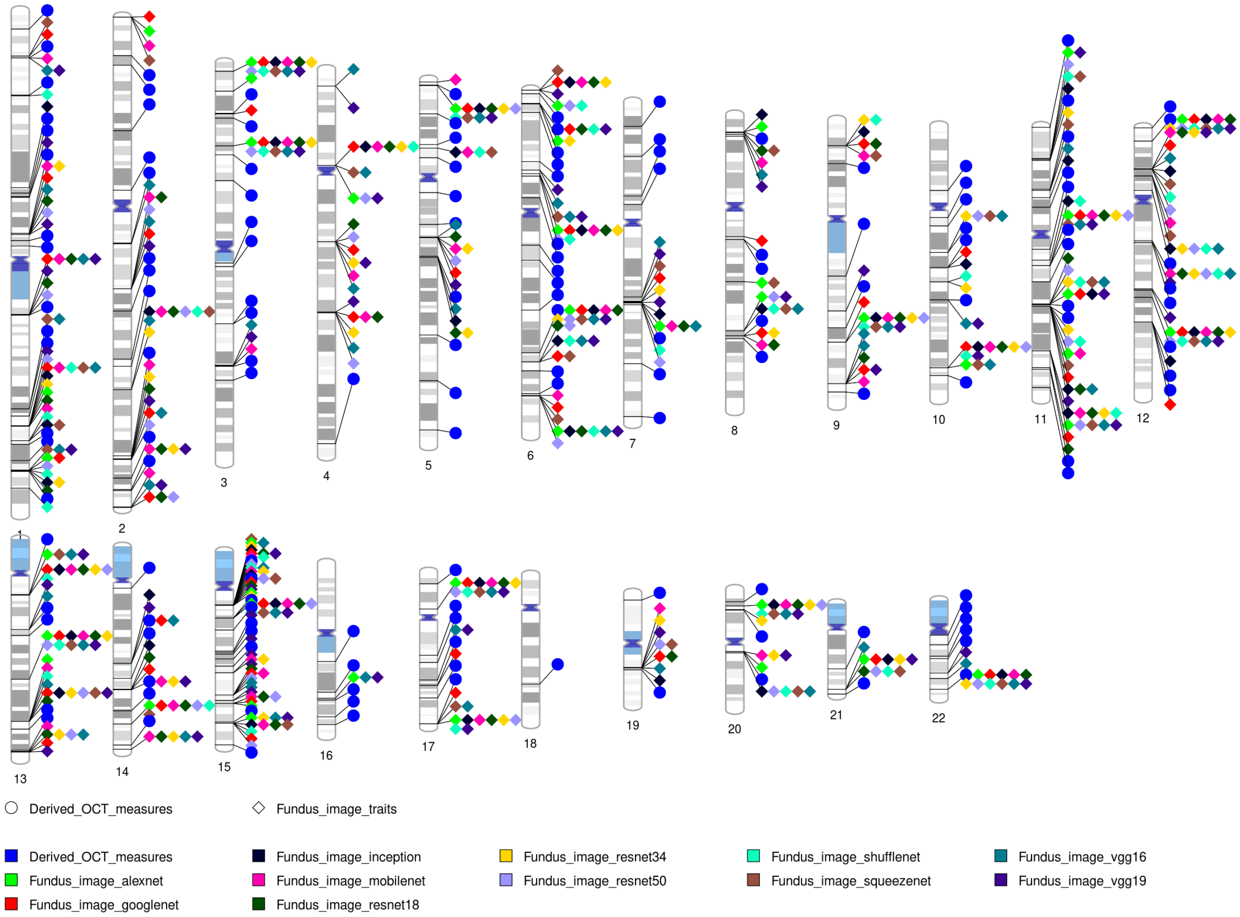


**Fig. S14 Ideogram of the 189 validated genomic regions associated with retinal imaging traits ($P < 3.20 \times 10^{-10}$).** We label the categories of retinal imaging traits with different colors.

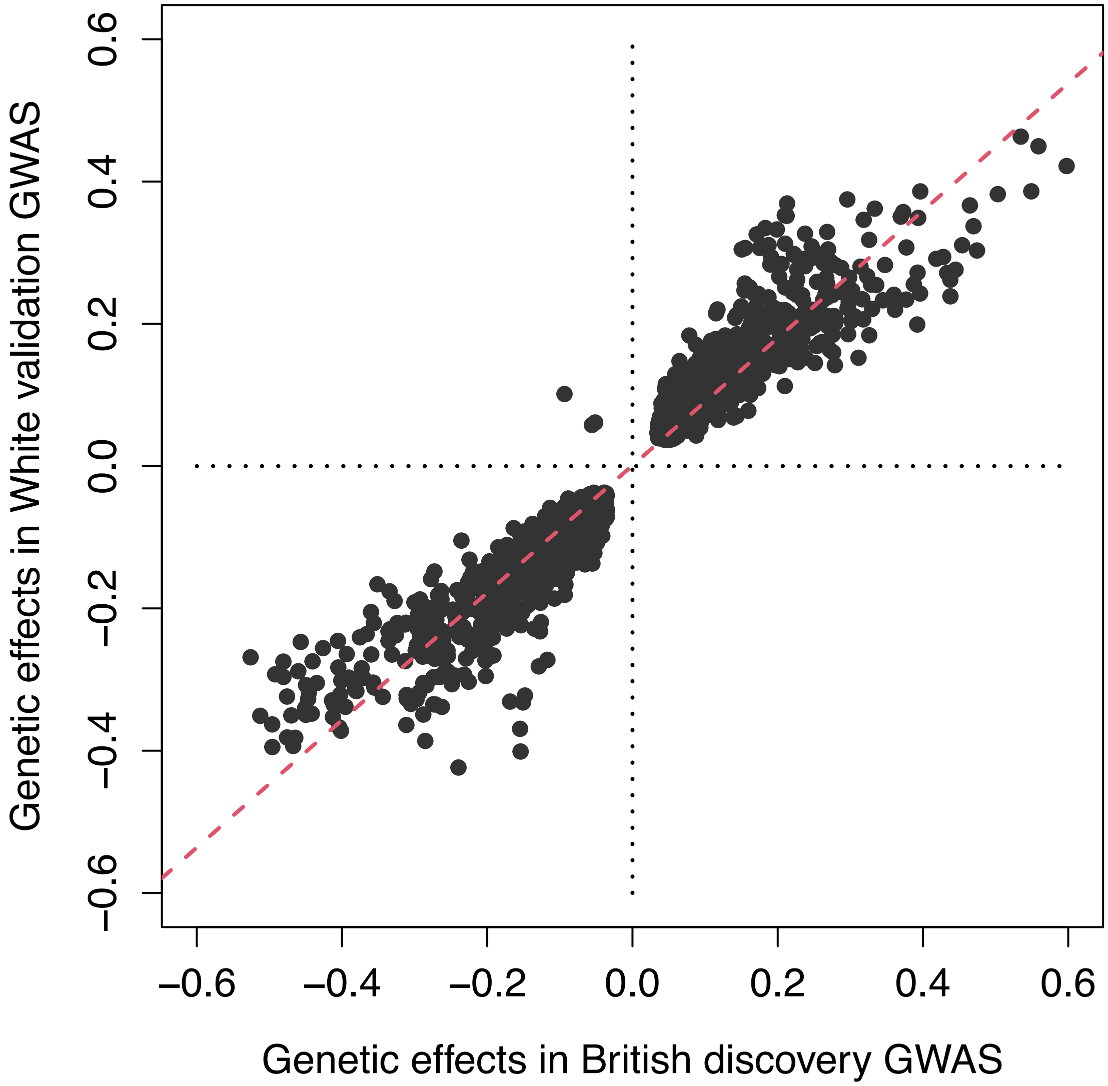


**Fig. S15 Relationship between significant genetic effects estimated in discovery GWAS and a European validation GWAS dataset.** For the significant ($P < 3.20 \times 10^{-10}$) imaging-variant associations identified in the discovery GWAS, we compared the genetic effects estimated in the discovery GWAS and those in an independent European validation GWAS. We illustrate the associations that were also significant in the validation GWAS at the nominal significance level (0.05).

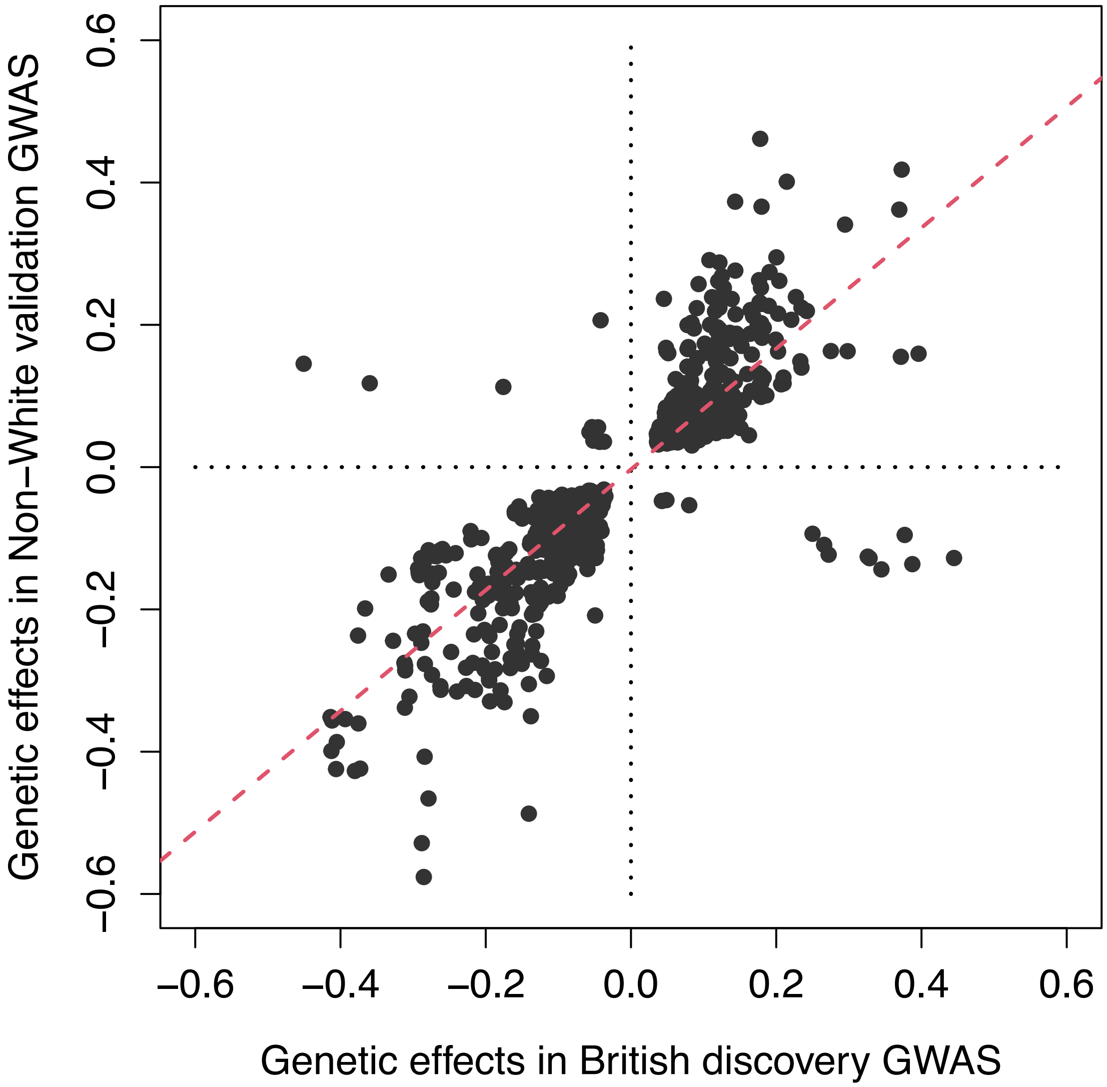


**Fig. S16 Relationship between significant genetic effects estimated in discovery GWAS and a non-European validation GWAS dataset.** For the significant ($P < 3.20 \times 10^{-10}$) imaging-variant associations identified in the discovery GWAS, we compared the genetic effects estimated in the discovery GWAS and those in an independent non-European validation GWAS. We illustrate the associations that were also significant in the validation GWAS at the nominal significance level (0.05).

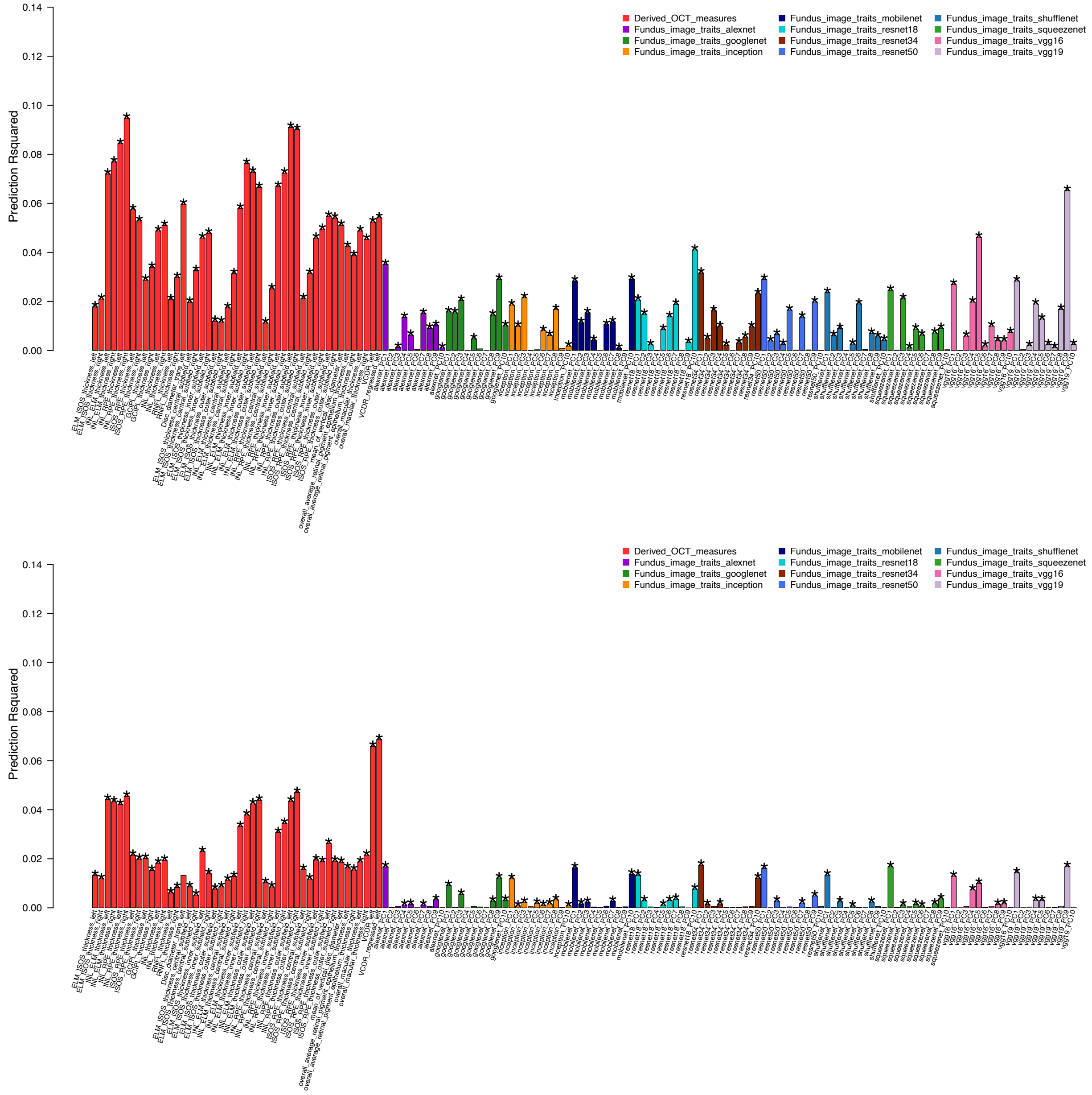


**Fig. S17 Performance of polygenic risk scores (PRS) of the 156 retinal imaging traits.** We developed the PRS using the summary statistics from the British GWAS and calculated the prediction R-squared in independent European (upper panel) and non-European (lower panel) validation datasets. The asterisks highlight significant predictions after FDR adjustment for multiple testing at a 5% level.

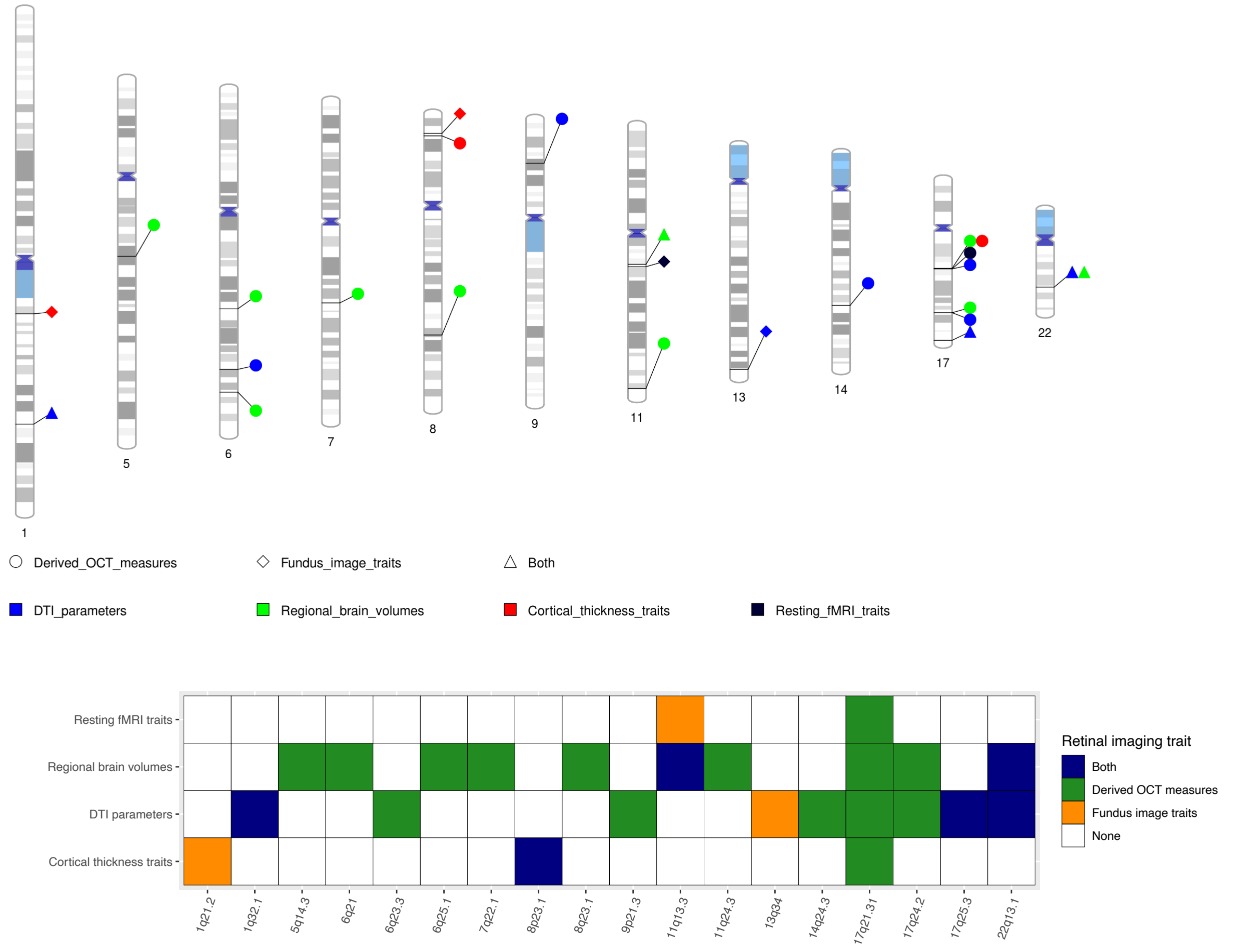


**Fig. S18 Ideogram of the 18 genomic regions associated with retinal and brain imaging traits.** In the upper panel, we label the categories of retinal imaging traits with different shapes and categories of brain imaging traits with different colors. In the lower panel, we further summarize the results into a table, where the *x*-axis represents the genomic regions and *y*-axis displays the categories of brain imaging traits. Each retinal imaging modality is labeled with a different color and the blue color is used when more than one retinal imaging modalities are observed in the locus.

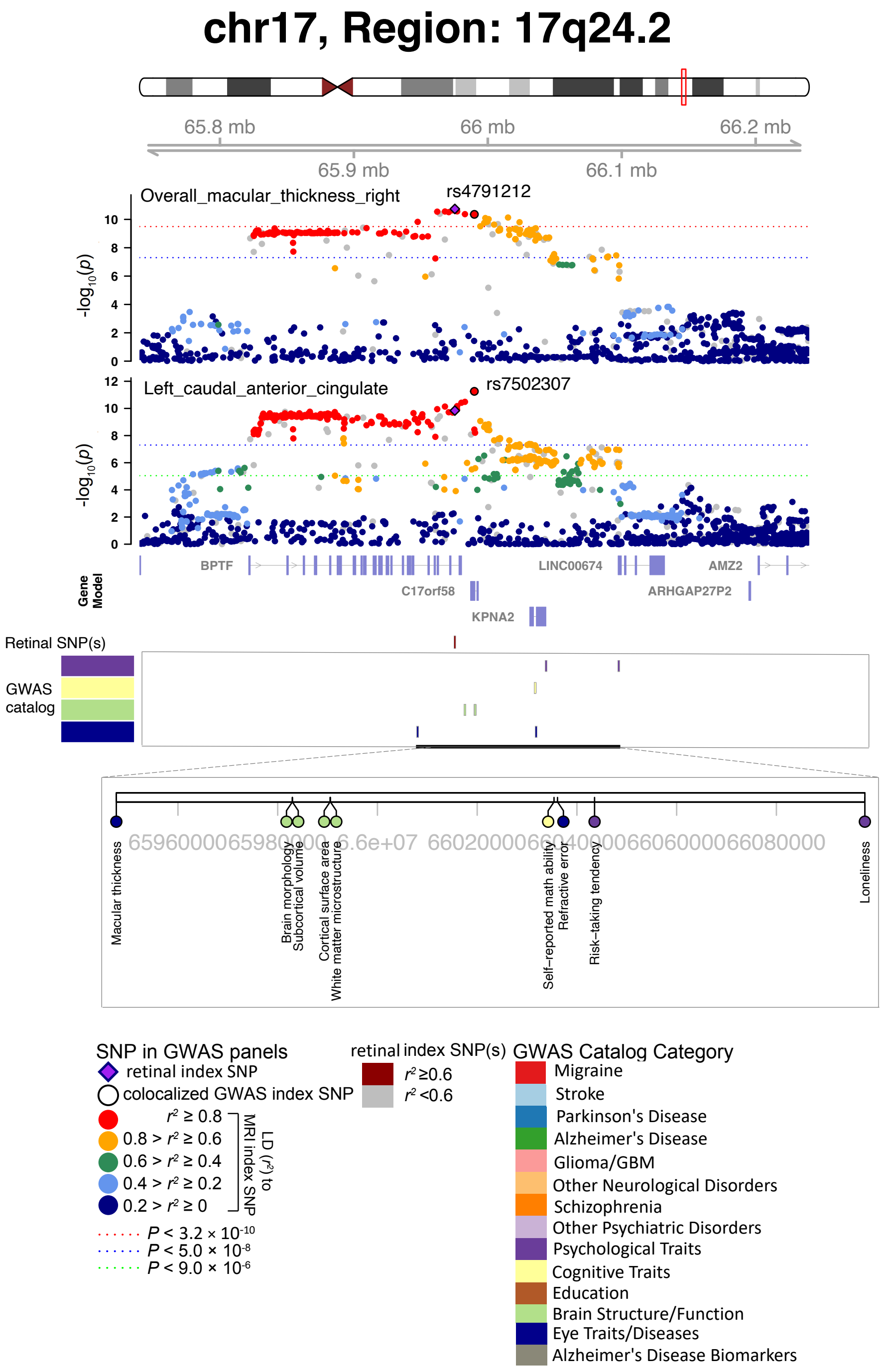


**Fig. S19 Selected genetic loci associated with retinal imaging traits and regional brain volumes.** In 17q24.2, we observed the shared association (LD $r^2 \geq 0.6$) between the retinal imaging trait (overall_macular_thickness_right, index variant rs4791212) and volume of the left caudal anterior cingulate (index variant rs7502307). We also observed shared associations with cognitive traits and psychological traits.

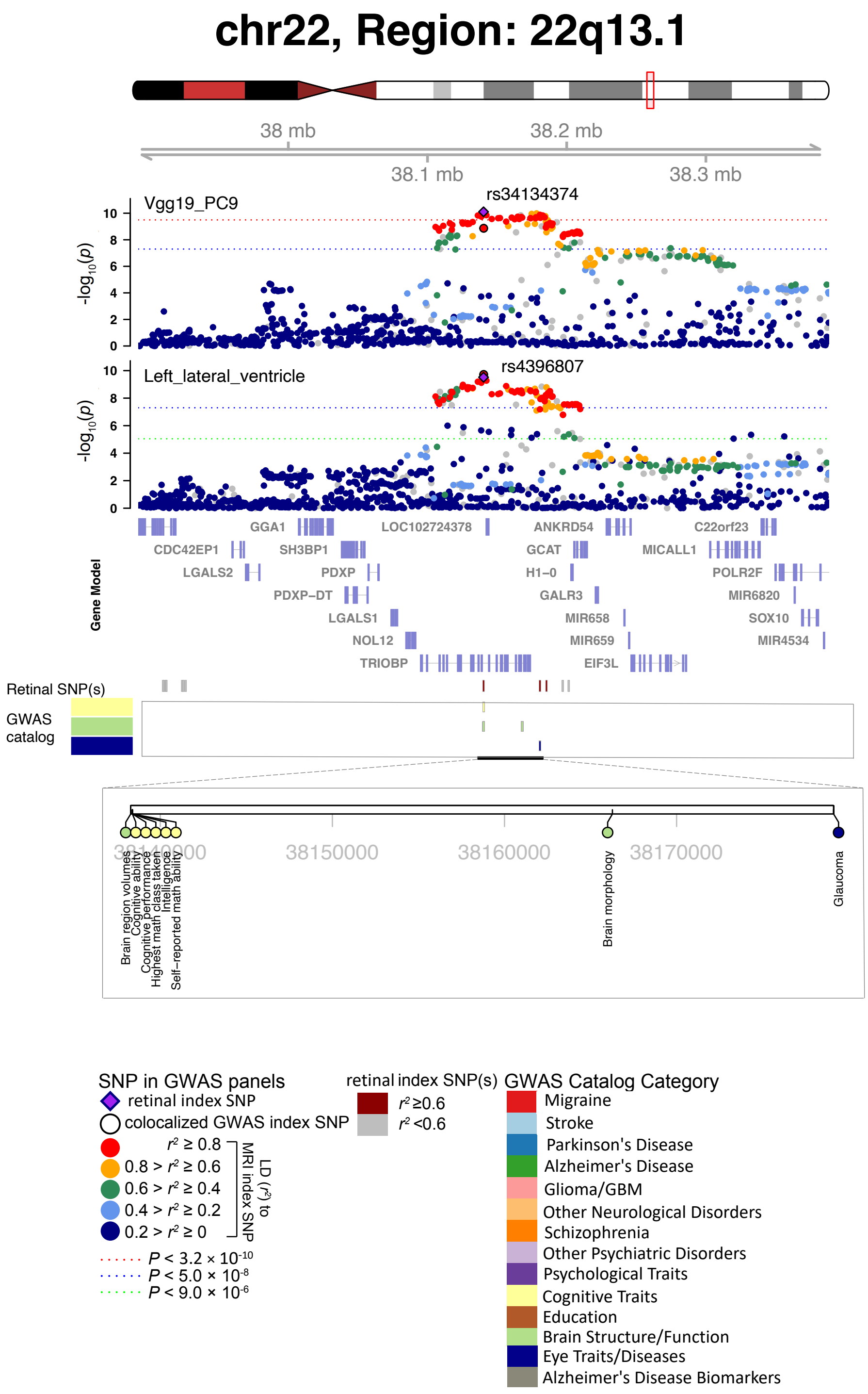


**Fig. S20 Selected genetic loci associated with retinal imaging traits and regional brain volumes.** In 22q13.1, we observed the shared association (LD $r^2 \geq 0.6$) between the retinal imaging trait (Vgg19_PC9, index variant rs34134374) and volume of the left lateral ventricle (index variant rs4396807). We also observed shared associations with cognitive traits.

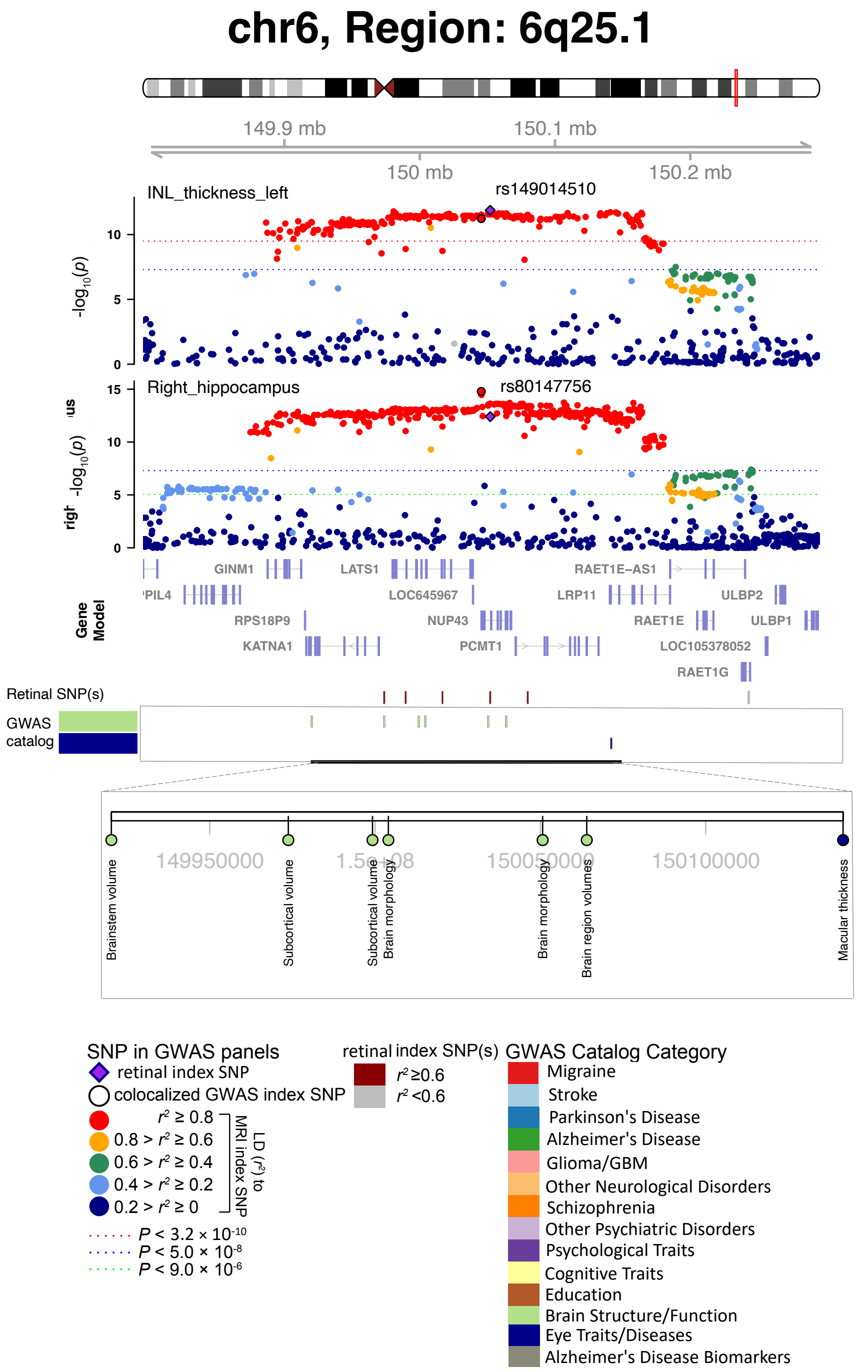


**Fig. S21 Selected genetic loci associated with retinal imaging traits and regional brain volumes.** In 6q25.1, we observed the shared association (LD $r^2 \geq 0.6$) between the retinal imaging trait (INL_thickness_left, index variant rs149014510) and volume of the right hippocampus (index variant rs80147756).

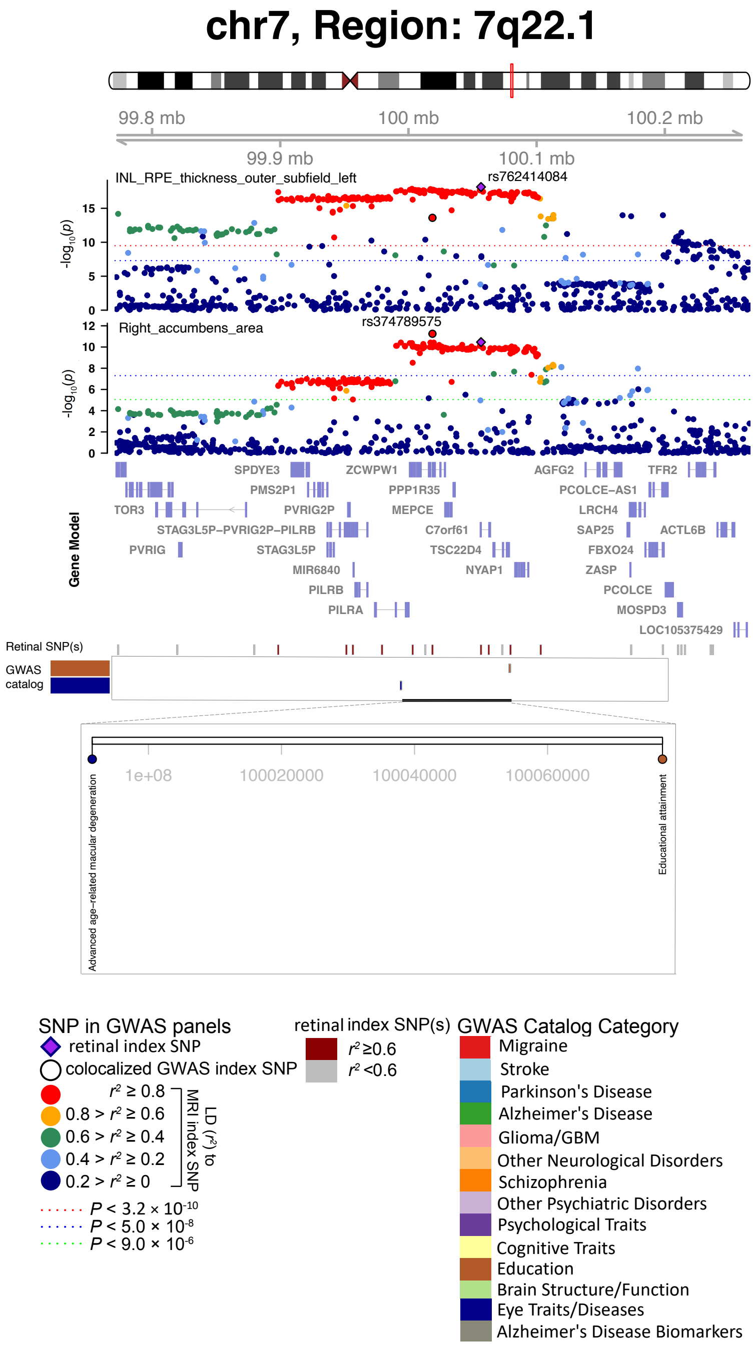


**Fig. S22 Selected genetic loci associated with retinal imaging traits and regional brain volumes.** In 7q22.1, we observed the shared association (LD $r^2 \geq 0.6$) between the retinal imaging trait (INL_RPE_thickness_outer_subfield_left, index variant rs762414084) and volume of the right accumbens area (index variant 374789575). We also observed shared associations with education.

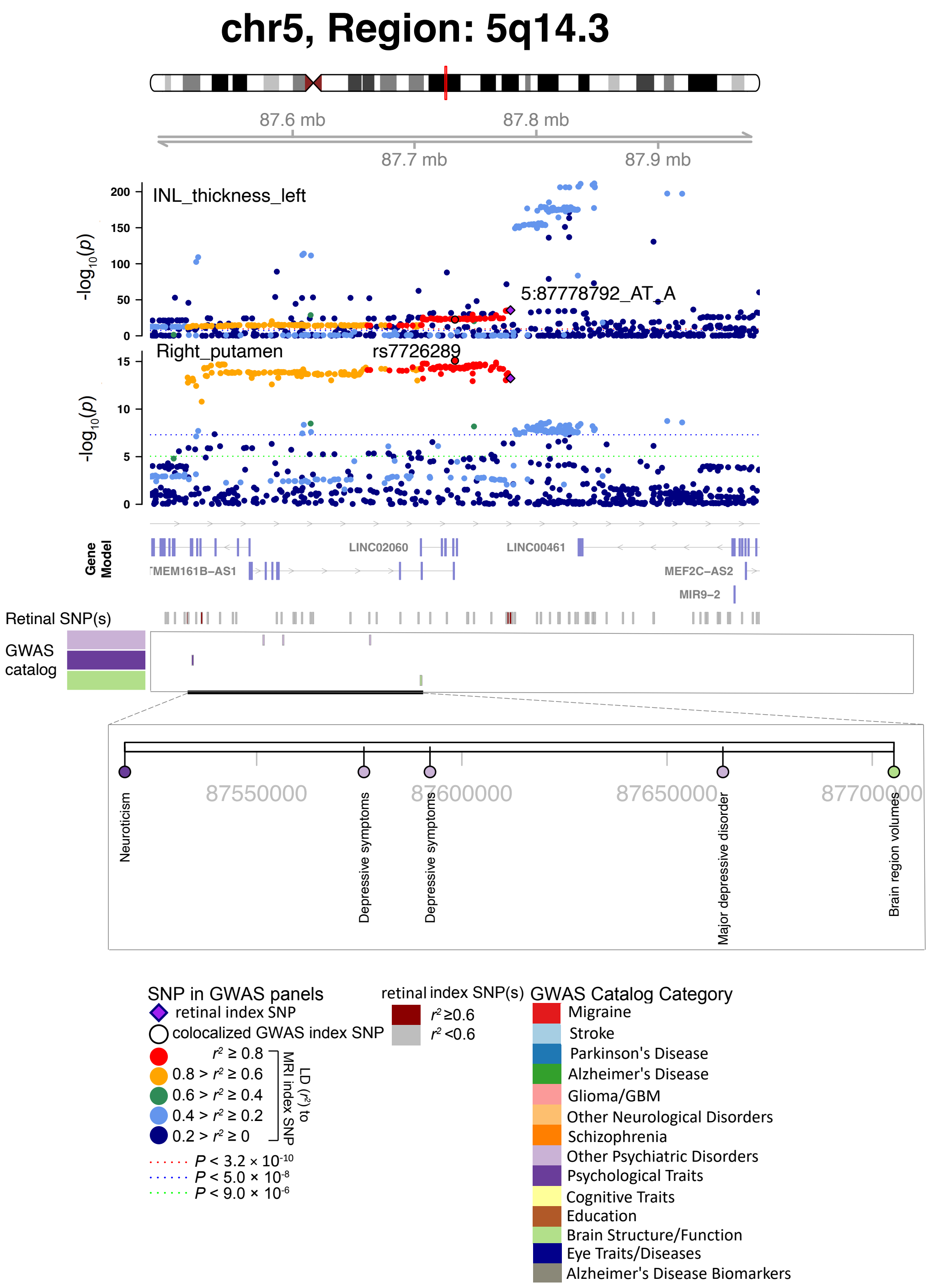


**Fig. S23 Selected genetic loci associated with retinal imaging traits and regional brain volumes.** In 5q14.3, we observed the shared association (LD $r^2 \geq 0.6$) between the retinal imaging trait (INL_thickness_left, index variant 5:87778792_AT_A) and volume of the right putamen (index variant rs7726289). We also observed shared associations with neuroticism and major depressive disorder.

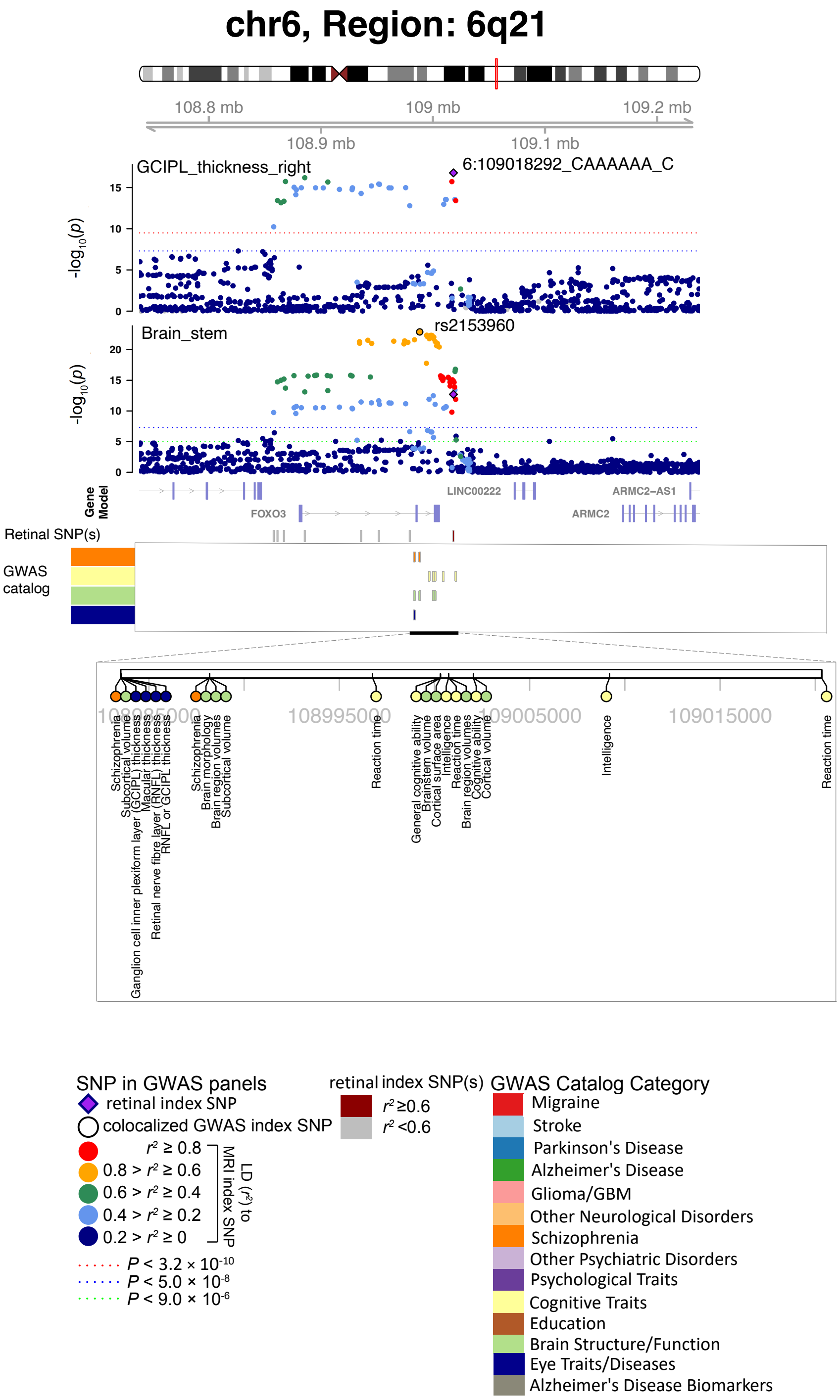


**Fig. S24 Selected genetic loci associated with retinal imaging traits and regional brain volumes.** In 6q21, we observed the shared association (LD $r^2 \geq 0.6$) between the retinal imaging trait (GCIPL_thickness_right, index variant 6:109018292_CAAAAAA_C) and volume of the brain stem (index variant rs2153960). We also observed shared associations with schizophrenia and cognitive traits.

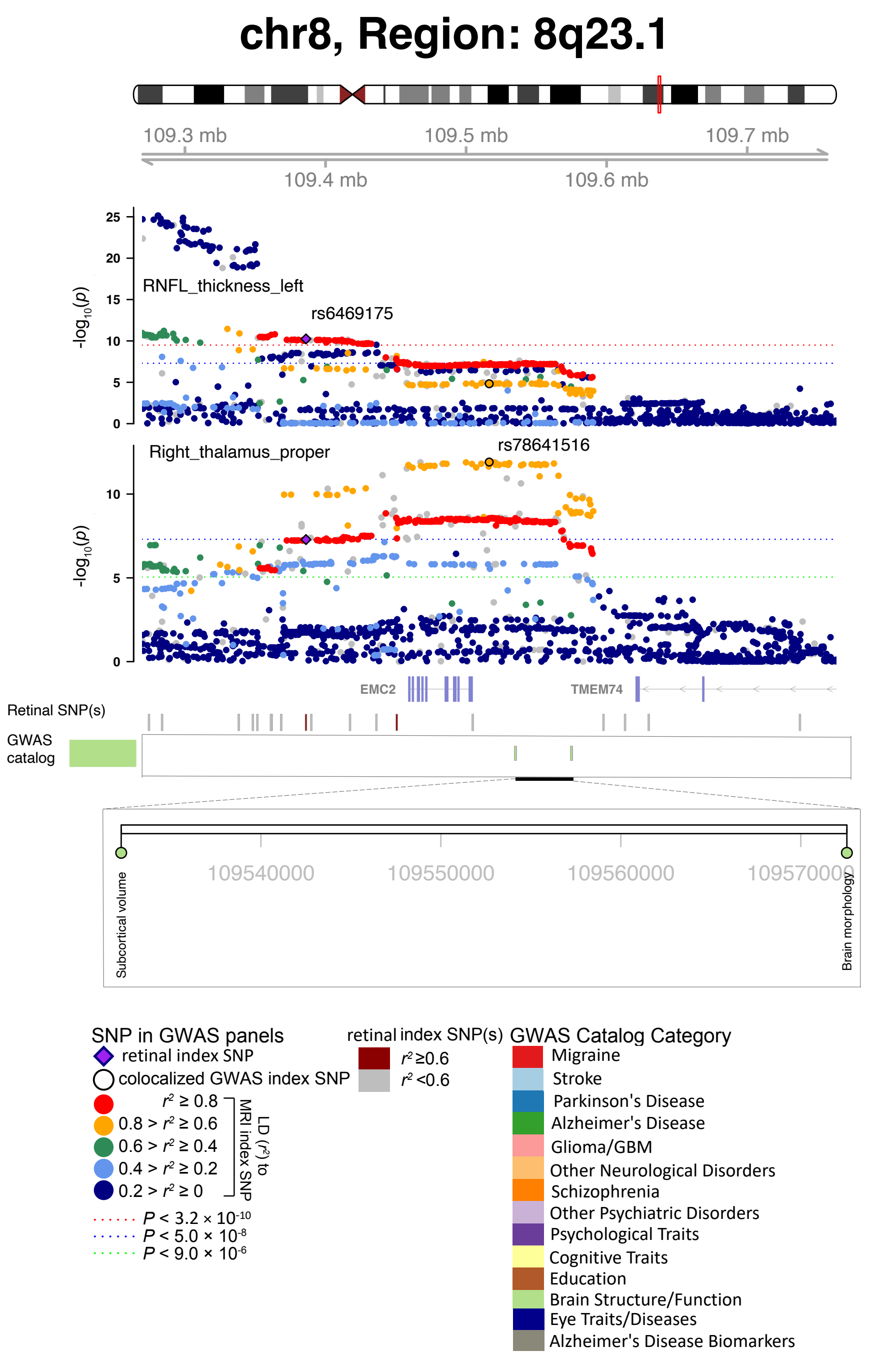


**Fig. S25 Selected genetic loci associated with retinal imaging traits and regional brain volumes.** In 8q23.1, we observed the shared association (LD $r^2 \geq 0.6$) between the retinal imaging trait (RNFL_thickness_left, index variant rs6469175) and volume of the right thalamus proper (index variant rs78641516).

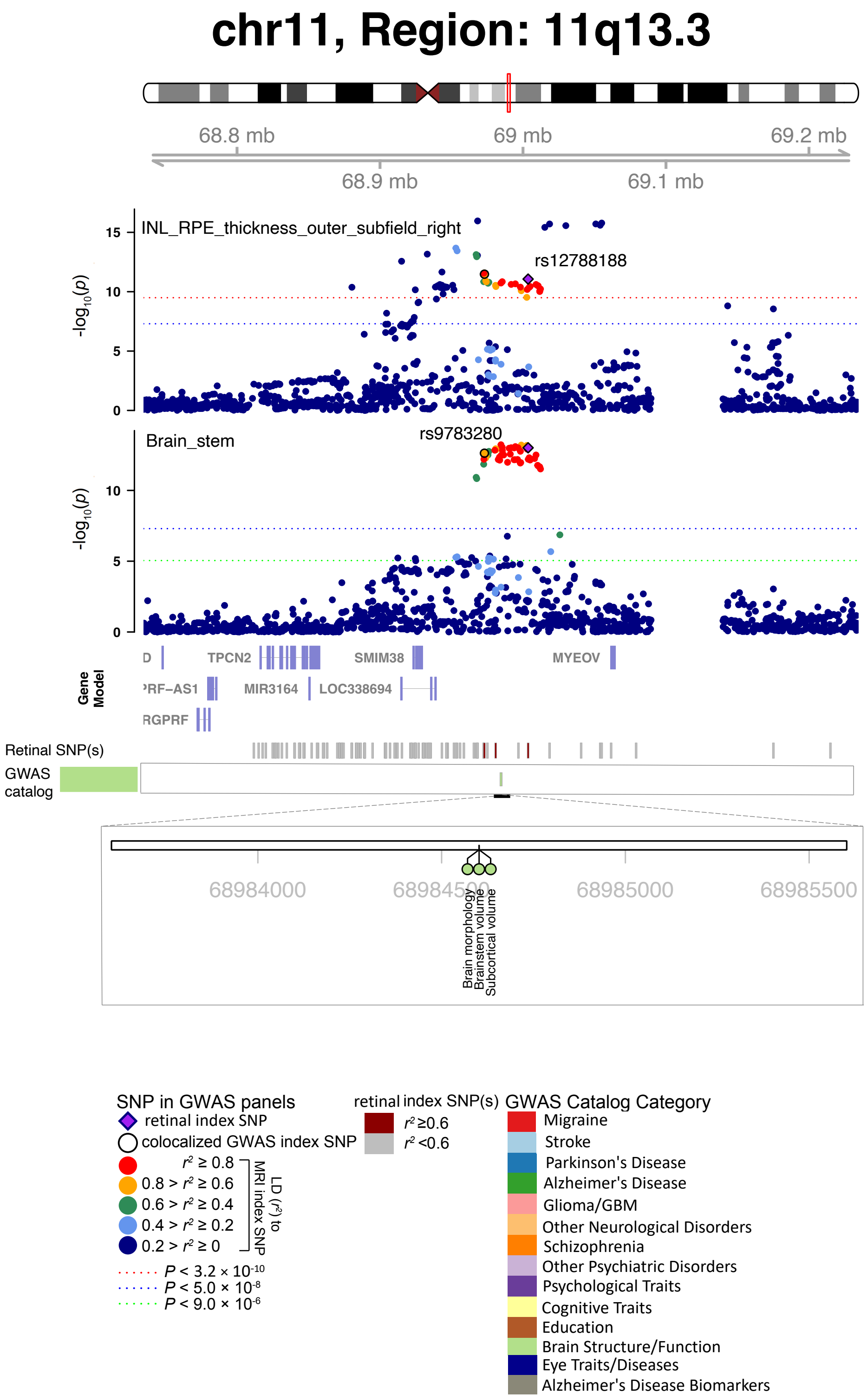


**Fig. S26 Selected genetic loci associated with retinal imaging traits and regional brain volumes.** In 11q13.3, we observed the shared association (LD $r^2 \geq 0.6$) between the retinal imaging trait (INL_RPE_thickness_outer_subfield_right, index variant rs12788188) and volume of the brain stem (index variant rs9783280).

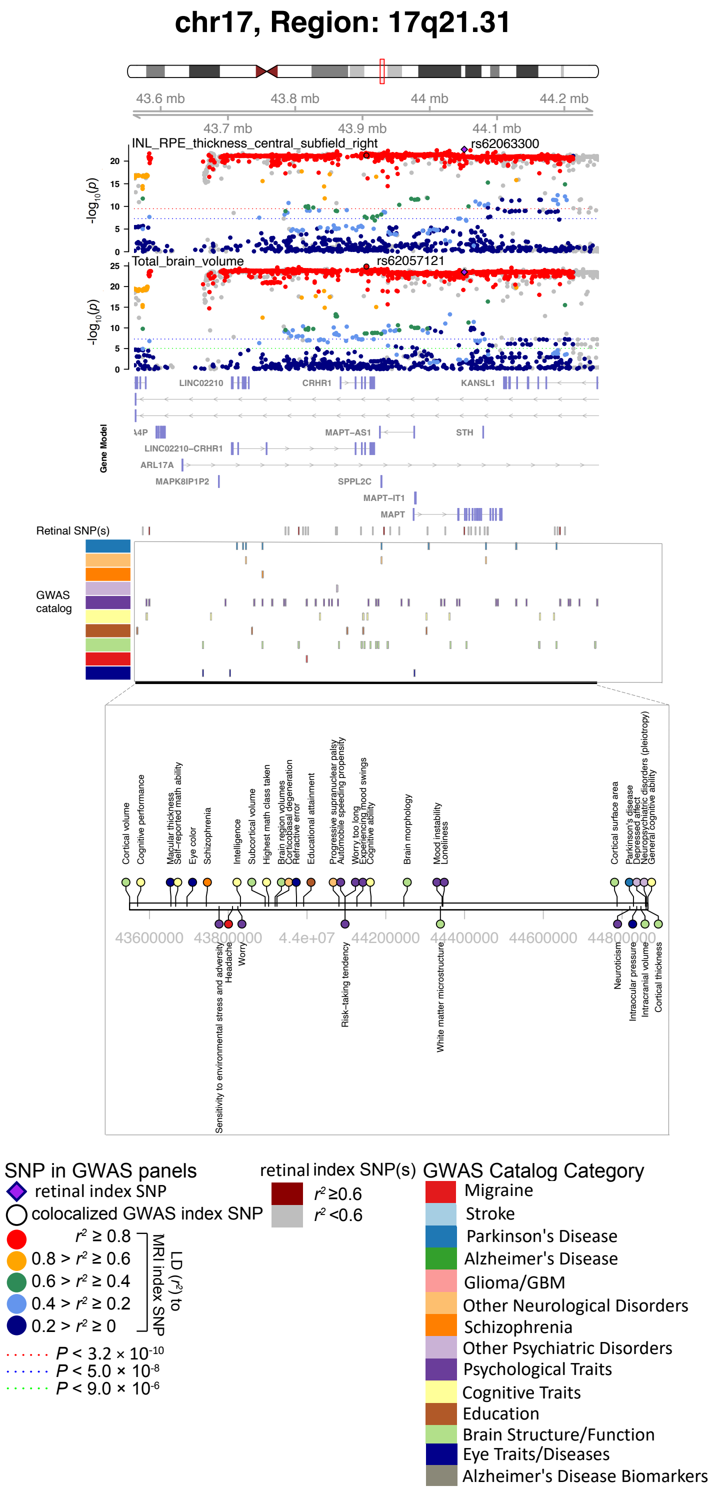


**Fig. S27 Selected genetic loci associated with retinal imaging traits and regional brain volumes.** In 17q21.31, we observed the shared association (LD $r^2 \geq 0.6$) between the retinal imaging trait (INL_RPE_thickness_central_subfield_right, index variant rs62063300) and the total brain volume (index variant rs62057121).

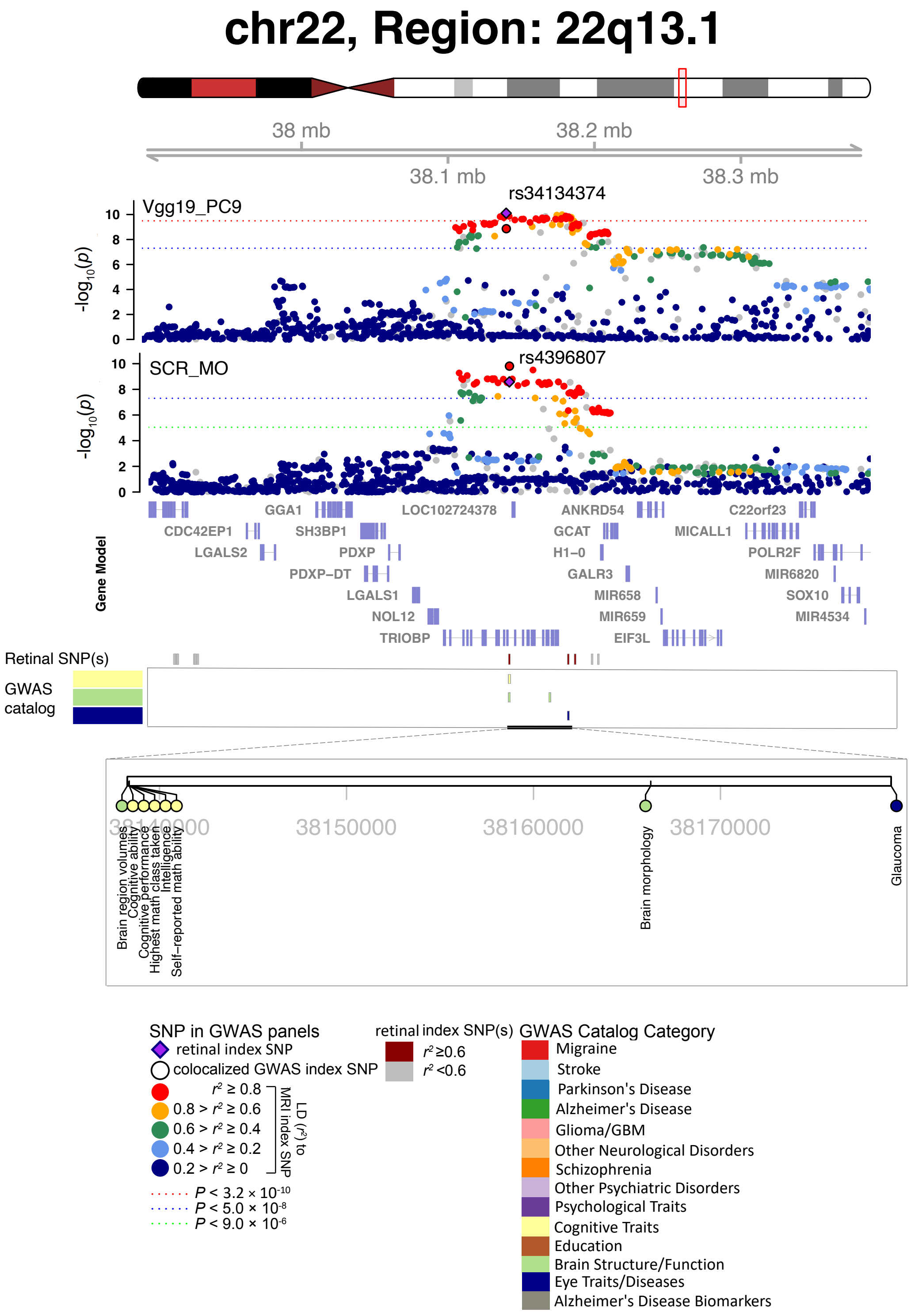


**Fig. S28 Selected genetic loci associated with retinal imaging traits and DTI parameters.** In 22q13.1, we observed the shared association (LD $r^2 \geq 0.6$) between the retinal imaging trait (Vgg19_PC9, index variant rs34134374) and the mean MO of the superior corona radiata (SCR_MO, index variant rs4396807). MO, mode of anisotropy.

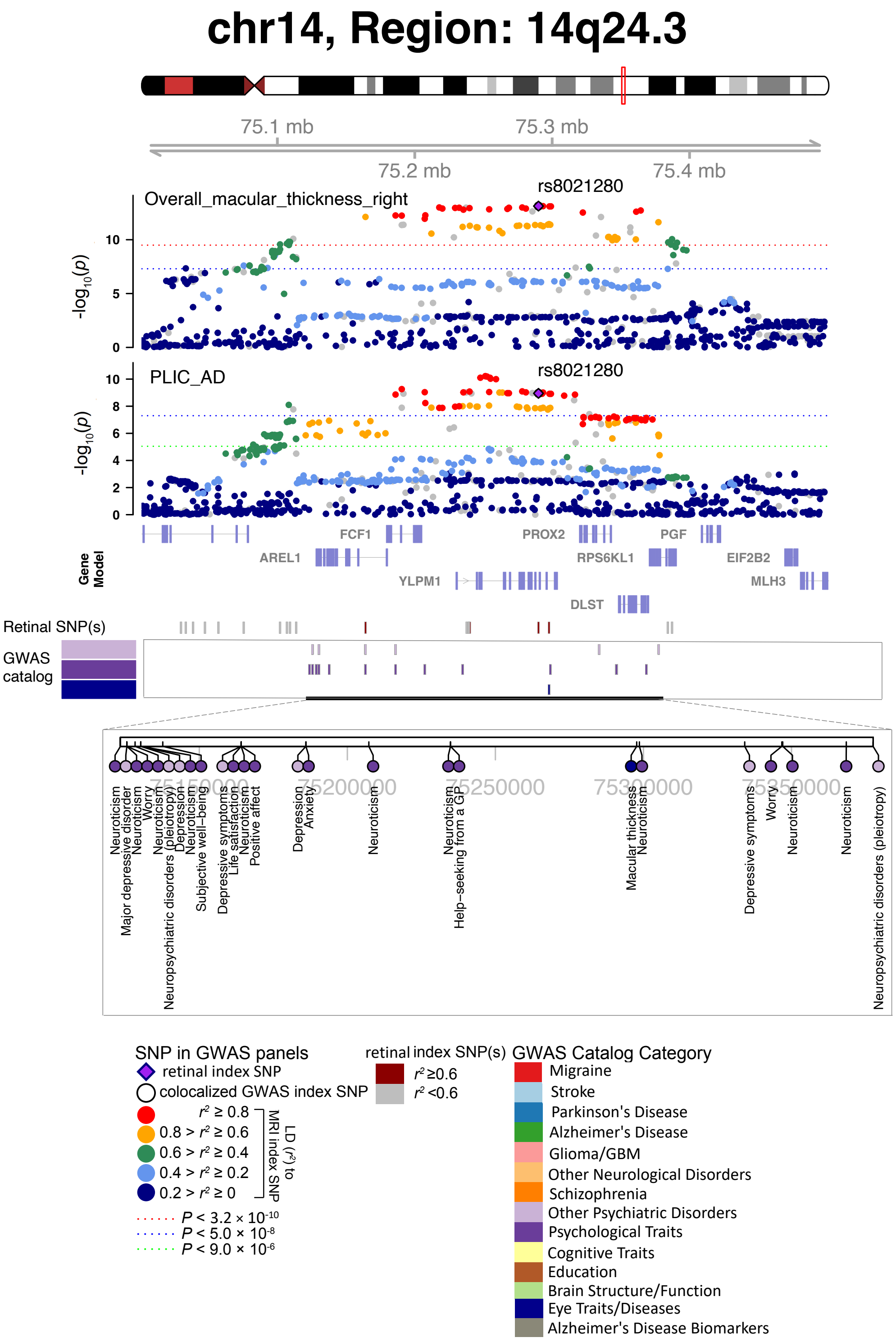


**Fig. S29 Selected genetic loci associated with retinal imaging traits and DTI parameters.** In 14q24.3, we observed the shared association (LD $r^2 \geq 0.6$) between the retinal imaging trait (overall_macular_thickness_right, index variant rs8021280) and the mean AD of the posterior limb of internal capsule (PLIC_AD, index variant rs8021280). AD, axial diffusivity.

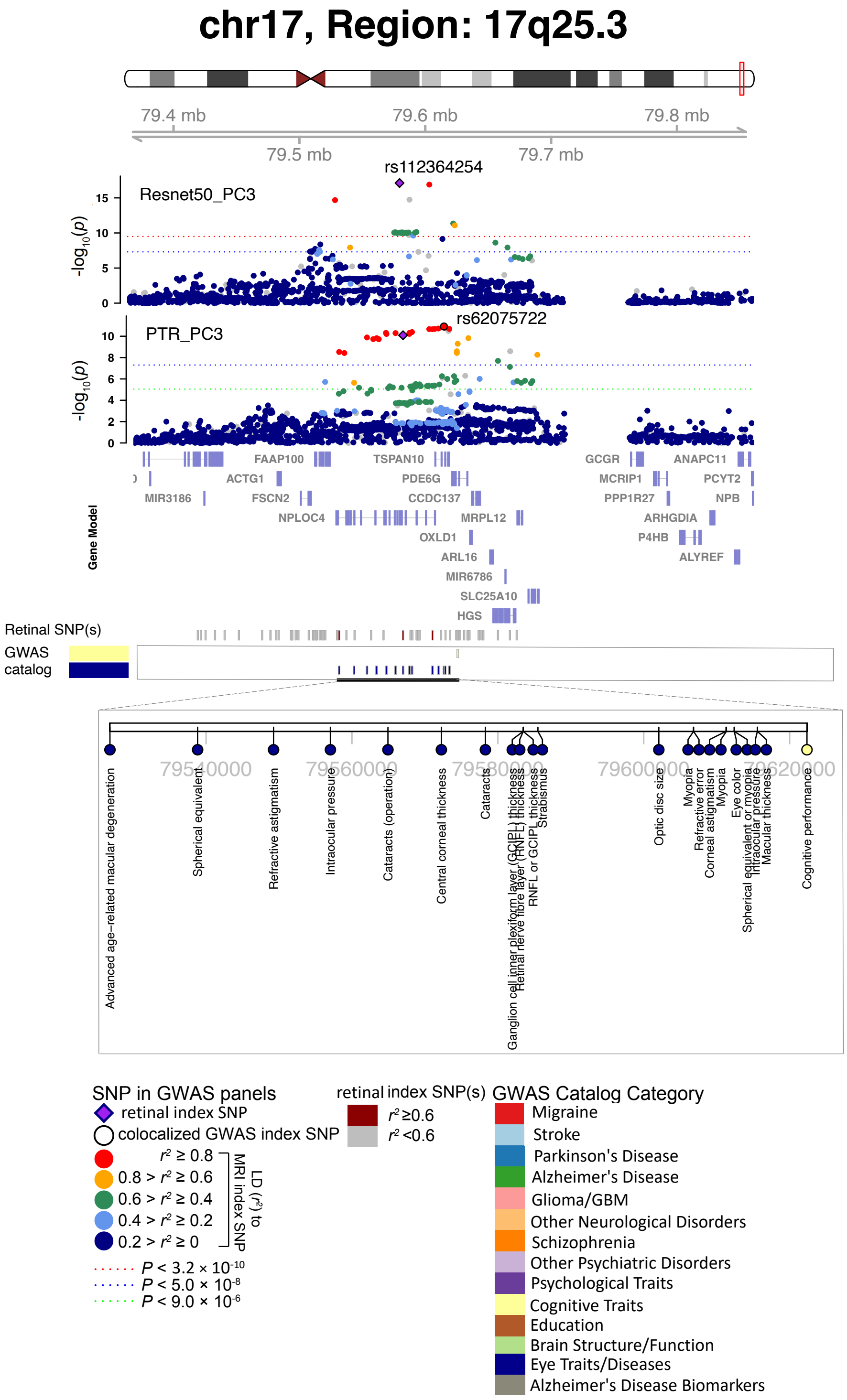


**Fig. S30 Selected genetic loci associated with retinal imaging traits and DTI parameters.** In 17q25.3, we observed the shared association (LD $r^2 \geq 0.6$) between the retinal imaging trait (Resnet50_PC3, index variant rs112364254) and the third PC of the FA of the posterior thalamic radiation (PTR_PC3, index variant rs62075722). FA, fractional anisotropy.

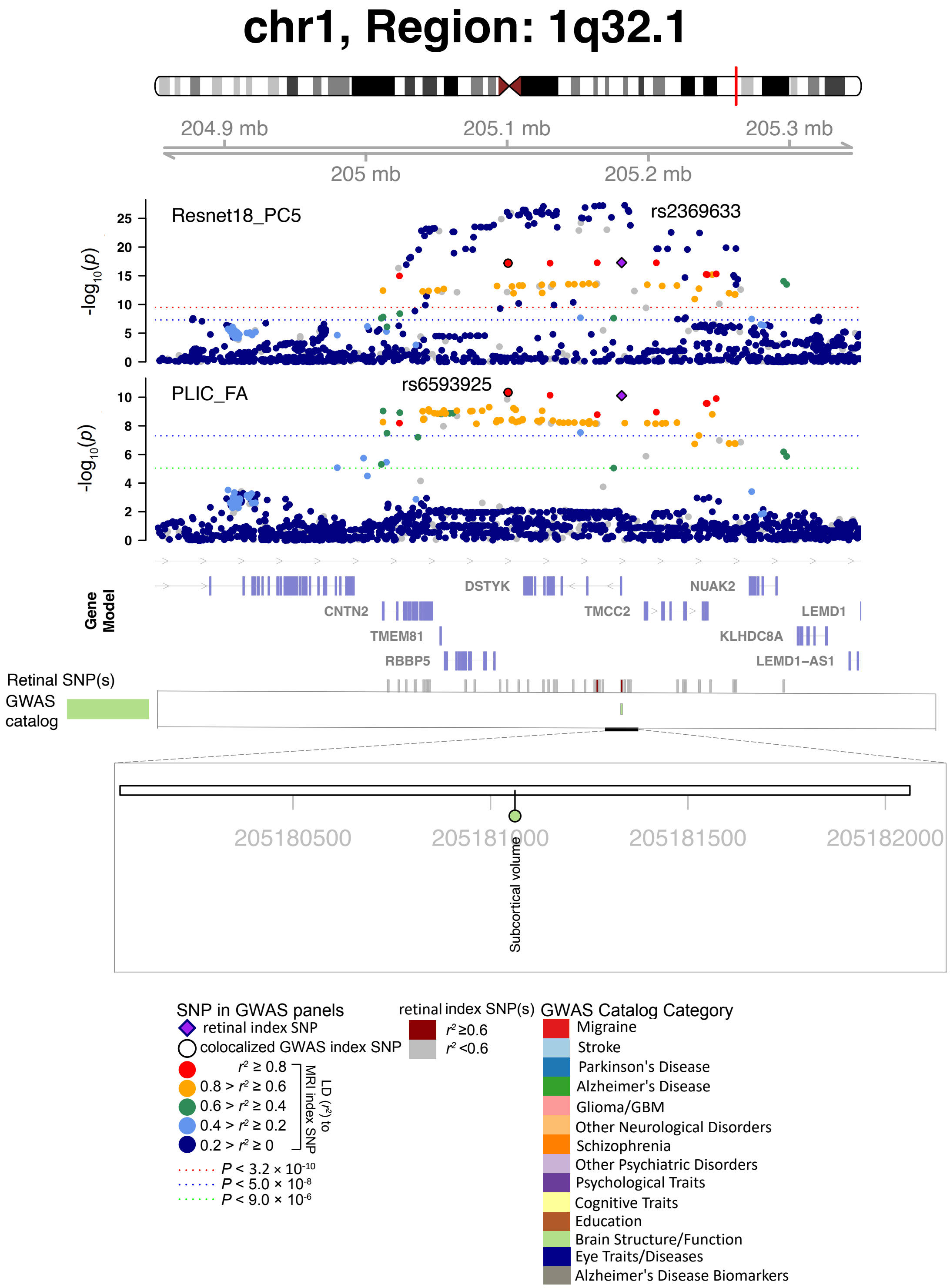


**Fig. S31 Selected genetic loci associated with retinal imaging traits and DTI parameters.** In 1q32.1, we observed the shared association (LD $r^2 \geq 0.6$) between the retinal imaging trait (Resnet18_PC5, index variant rs2369633) and the mean FA of posterior limb of internal capsule (PLIC_FA, index variant rs6593925). FA, fractional anisotropy.

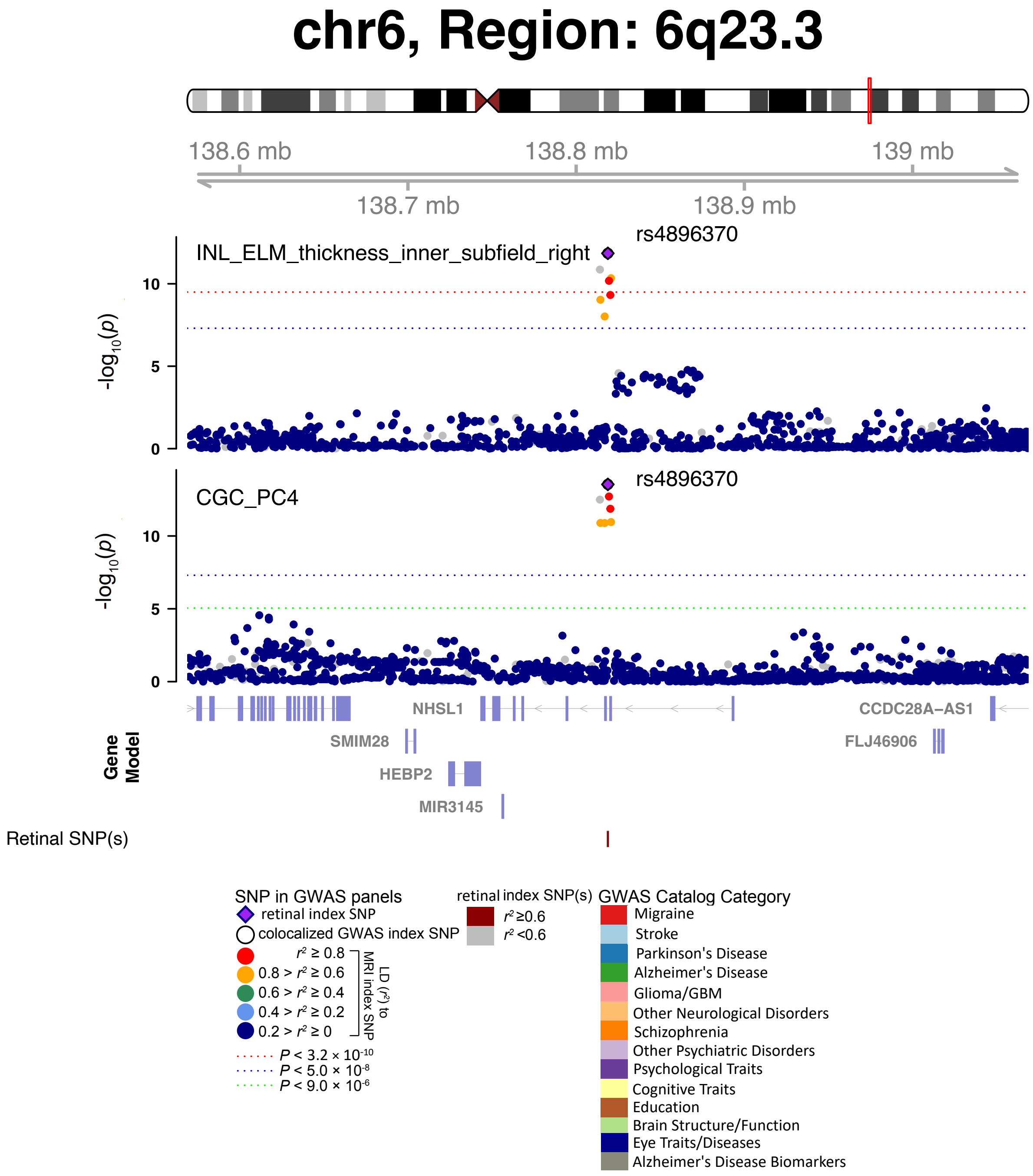


**Fig. S32 Selected genetic loci associated with retinal imaging traits and DTI parameters.** In 6q23.3, we observed the shared association (LD $r^2 \geq 0.6$) between the retinal imaging trait (INL_ELM_thickness_inner_subfield_right, index variant rs4896370) and the fourth PC of the FA of the cingulum (CGC_PC4, index variant rs4896370). FA, fractional anisotropy.

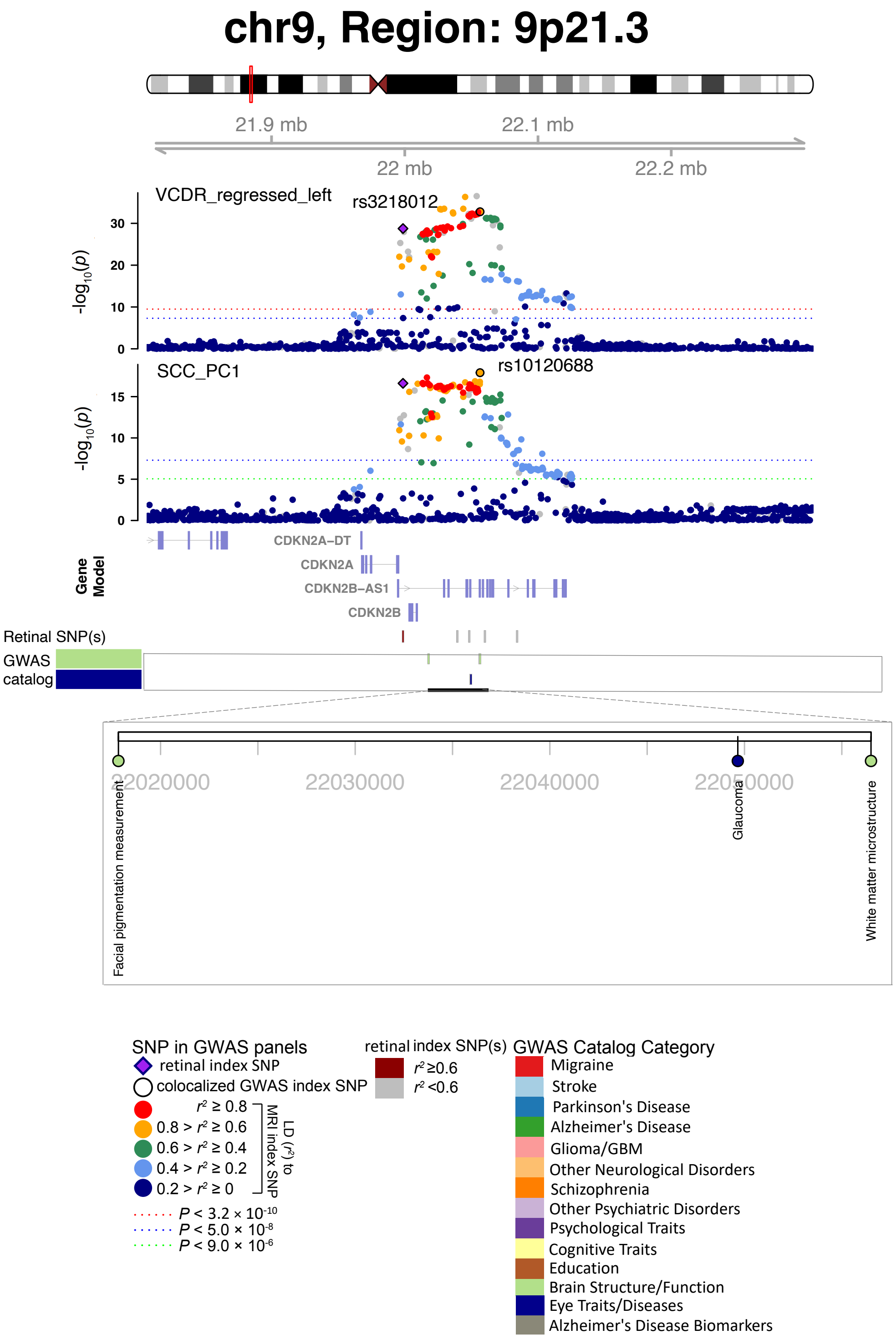


**Fig. S33 Selected genetic loci associated with retinal imaging traits and DTI parameters.** In 9p21.3, we observed the shared association (LD $r^2 \geq 0.6$) between the retinal imaging trait (VCDR_regressed_left, index variant rs3218012) and the first PC of the FA of the splenium of corpus callosum (SCC_PC1, index variant rs10120688). FA, fractional anisotropy.

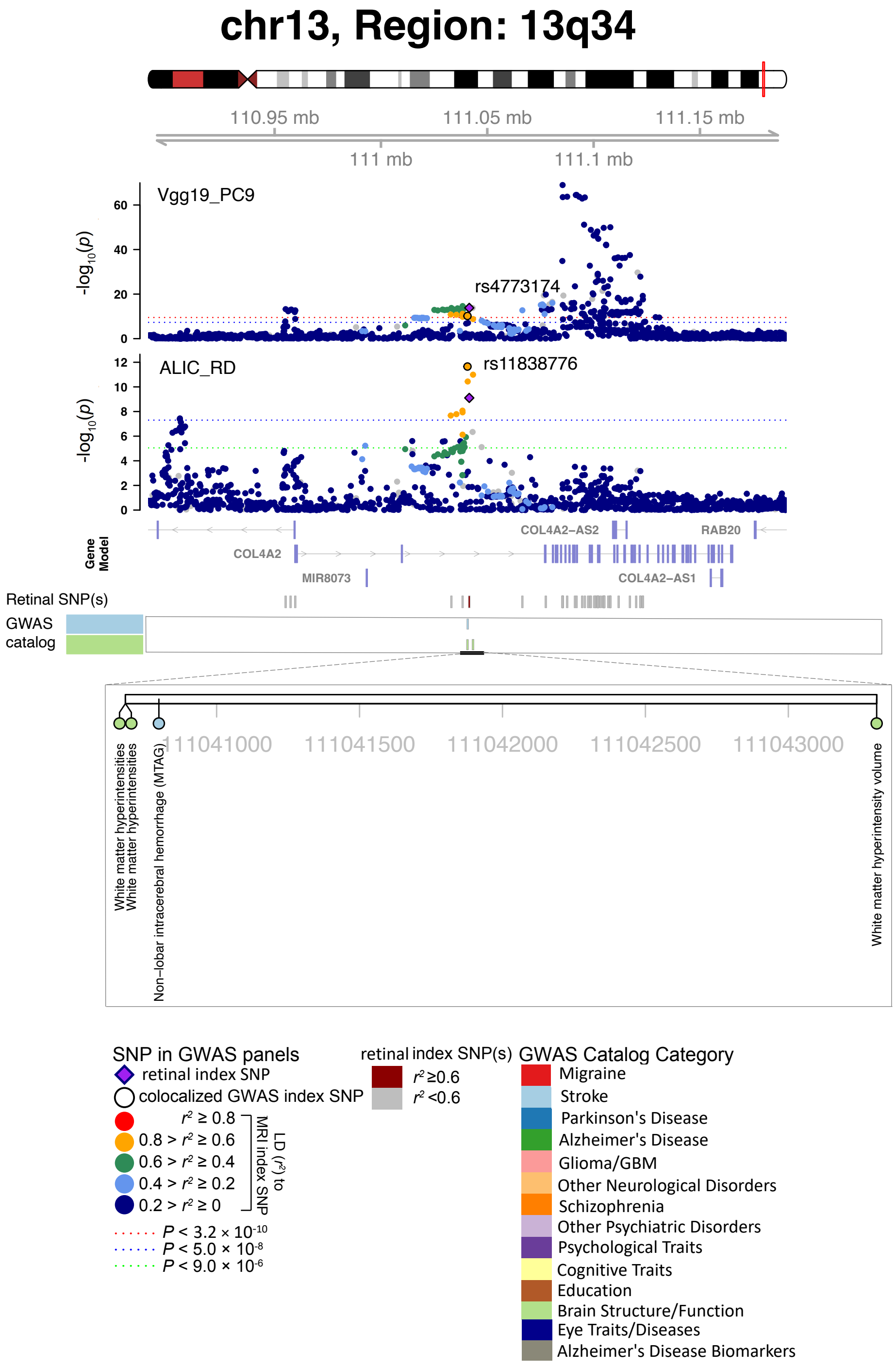


**Fig. S34 Selected genetic loci associated with retinal imaging traits and DTI parameters.** In 13q34, we observed the shared association (LD $r^2 \geq 0.6$) between the retinal imaging trait (Vgg19_PC9, index variant rs4773174) and the mean RD of the anterior limb of internal capsule (ALIC_RD, index variant rs11838776). RD, radial diffusivity.

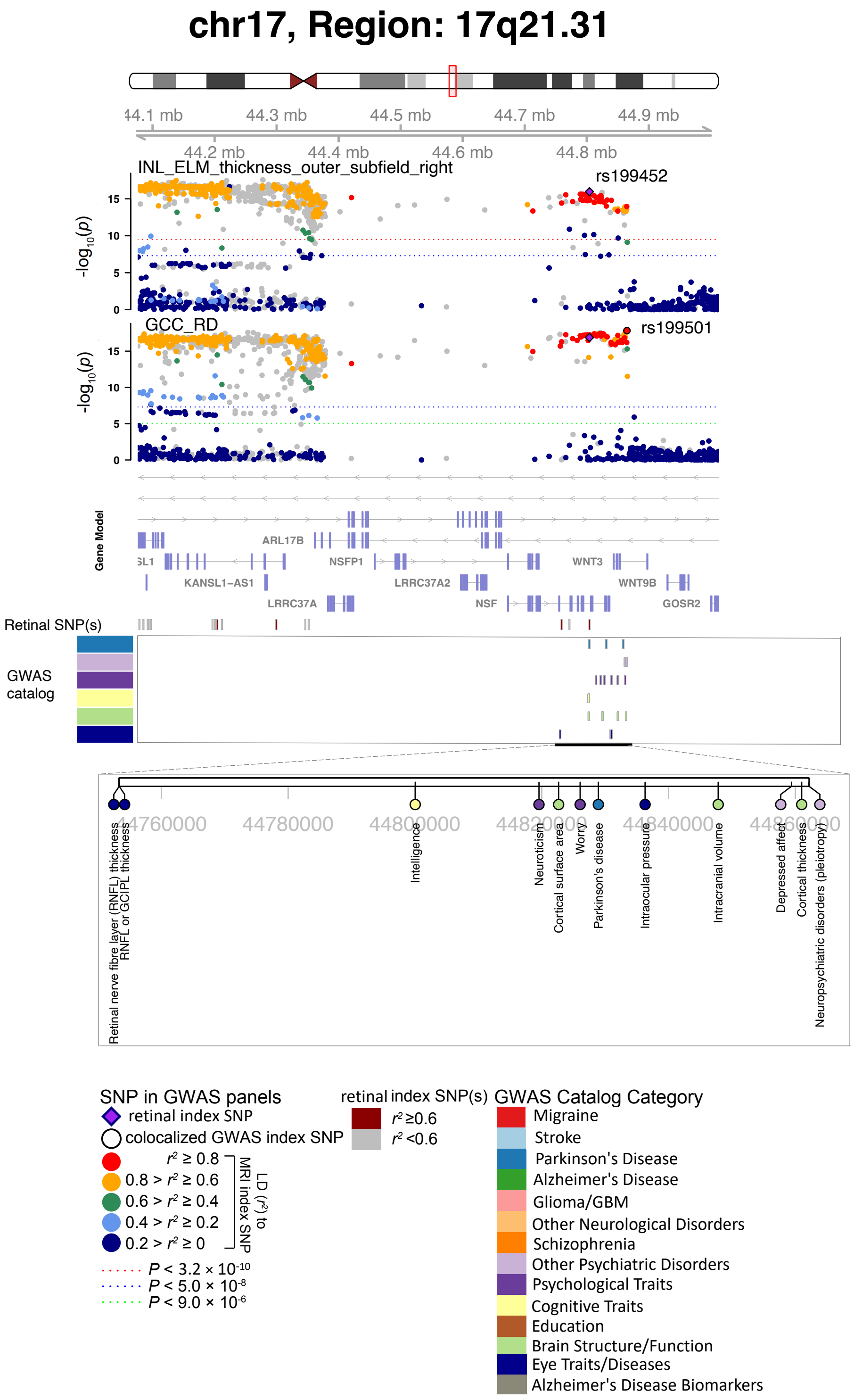


**Fig. S35 Selected genetic loci associated with retinal imaging traits and DTI parameters.** In 17q21.31, we observed the shared association (LD $r^2 \geq 0.6$) between the retinal imaging trait (INL_ELM_thickness_outer_subfield_right, index variant rs199452) and the mean RD of the genu of corpus callosum (GCC_RD, index variant rs199501). RD, radial diffusivity.

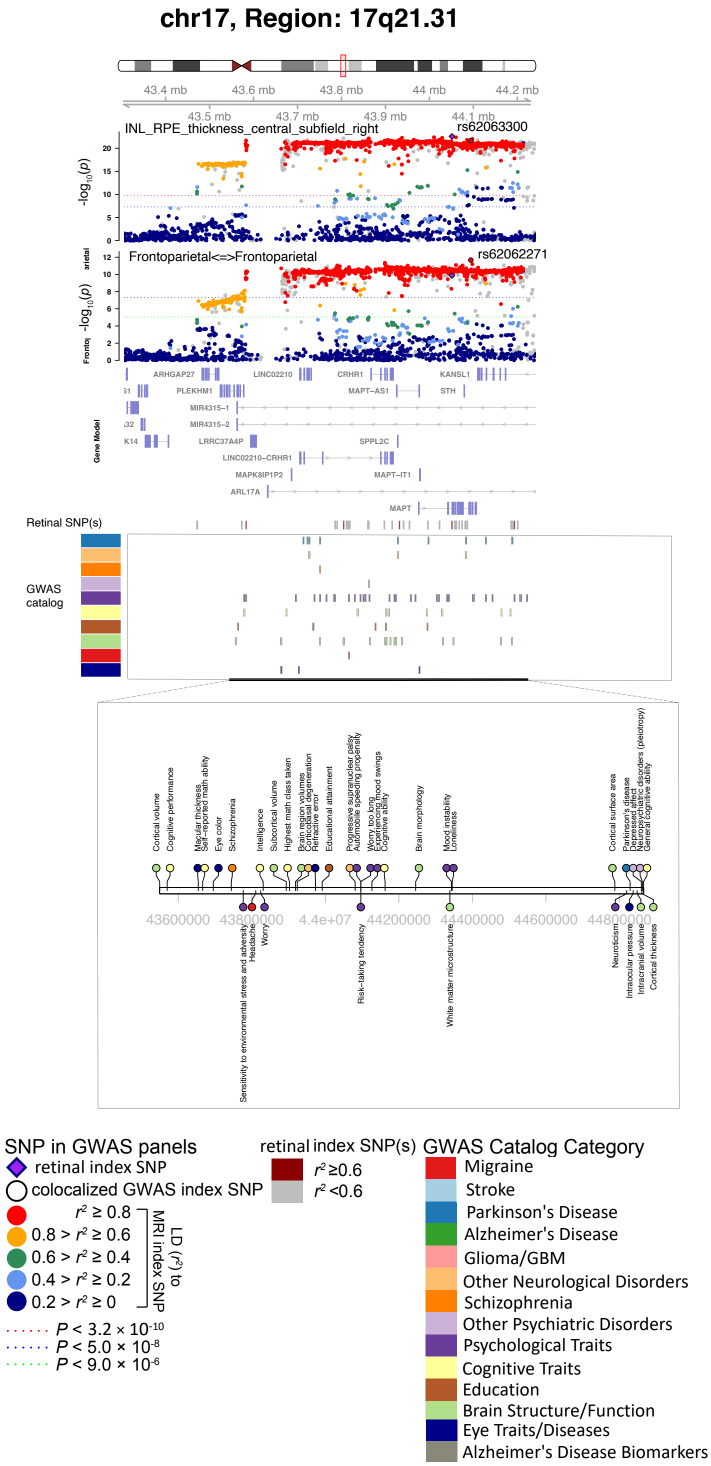


**Fig. S36 Selected genetic loci associated with retinal imaging traits and fMRI traits.** In 17q21.31, we observed the shared association (LD $r^2 \geq 0.6$) between the retinal imaging trait (INL_RPE_thickness_central_subfield_right, index variant rs62063300) and the functional connectivity of the frontoparietal network (Frontoparietal<=>Frontoparietal, index variant rs62062271).

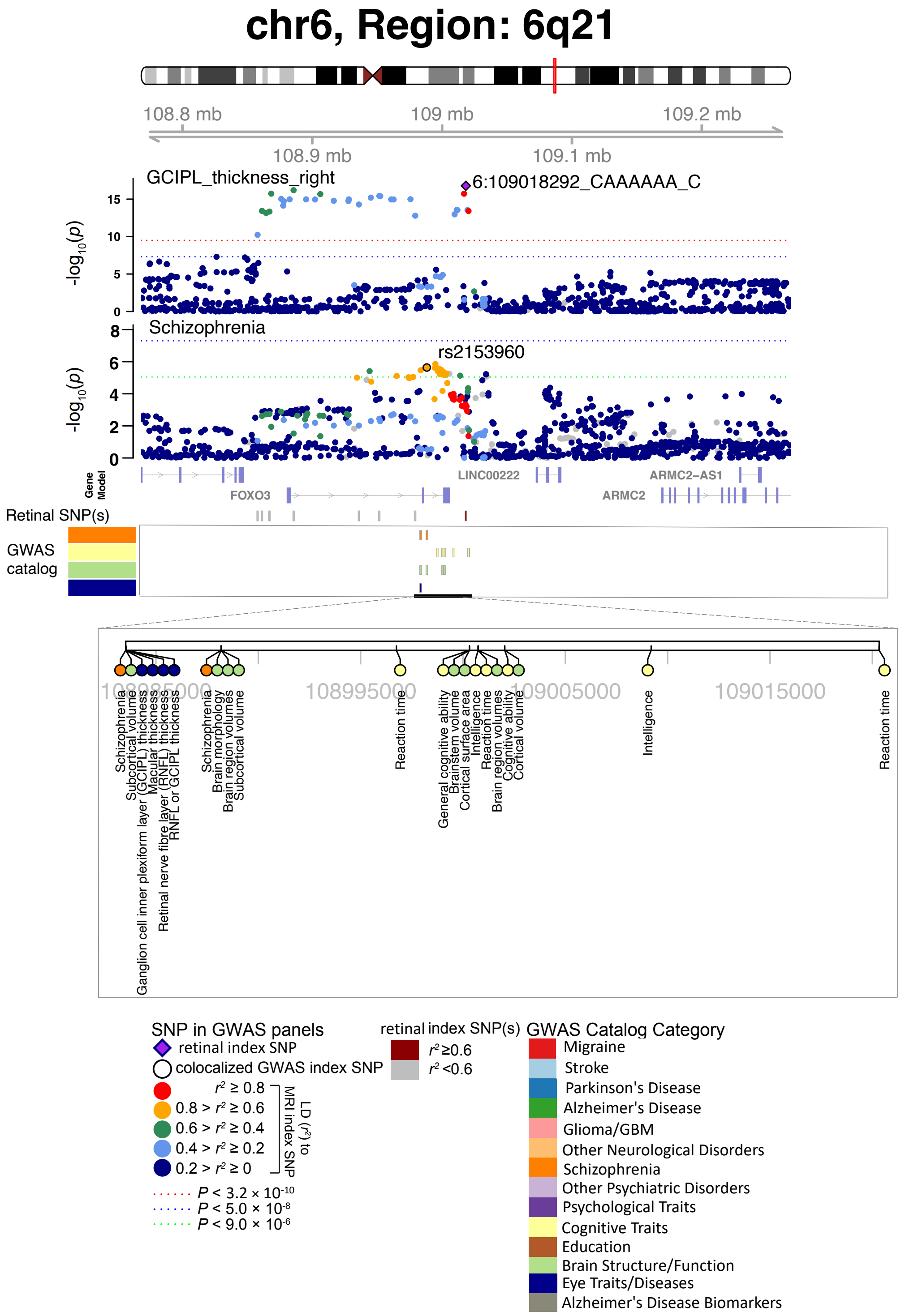


**Fig. S37 Selected genetic loci associated with retinal imaging traits and brain-related complex traits and diseases.** In 6q21, we observed the shared association (LD $r^2 \geq 0.6$) between the retinal imaging trait (GCIPL_thickness_right, index variant 6:109018292_CAAAAAA_C) and schizophrenia (index variant rs2153960). We also observed shared associations with intelligence, general cognitive ability, and reaction time.

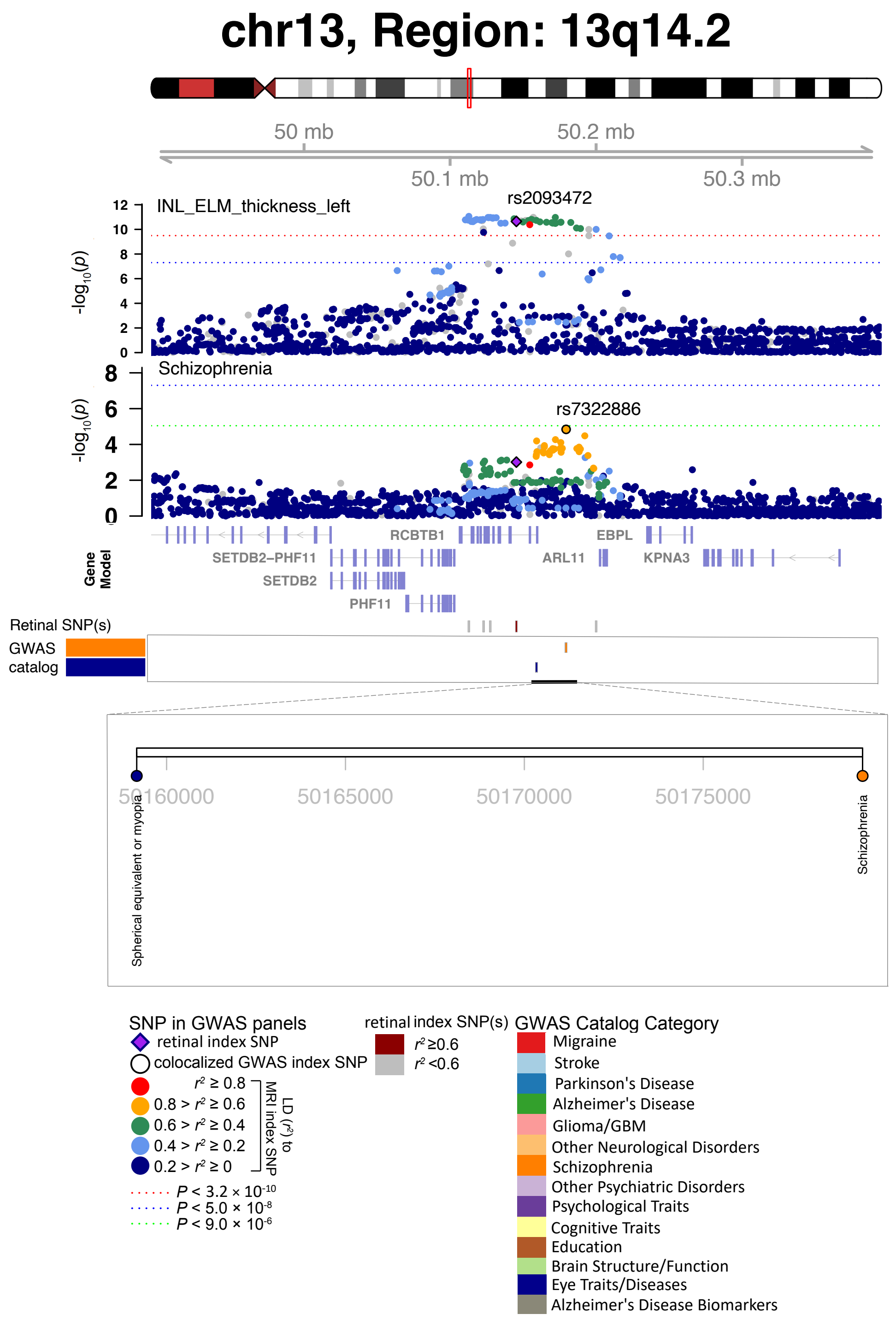


**Fig. S38 Selected genetic loci associated with retinal imaging traits and brain-related complex traits and diseases.** In 13q14.2, we observed the shared association (LD $r^2 \geq 0.6$) between the retinal imaging trait (INL_ELM_thickness_left, index variant rs2093472) and schizophrenia (index variant rs7322886).

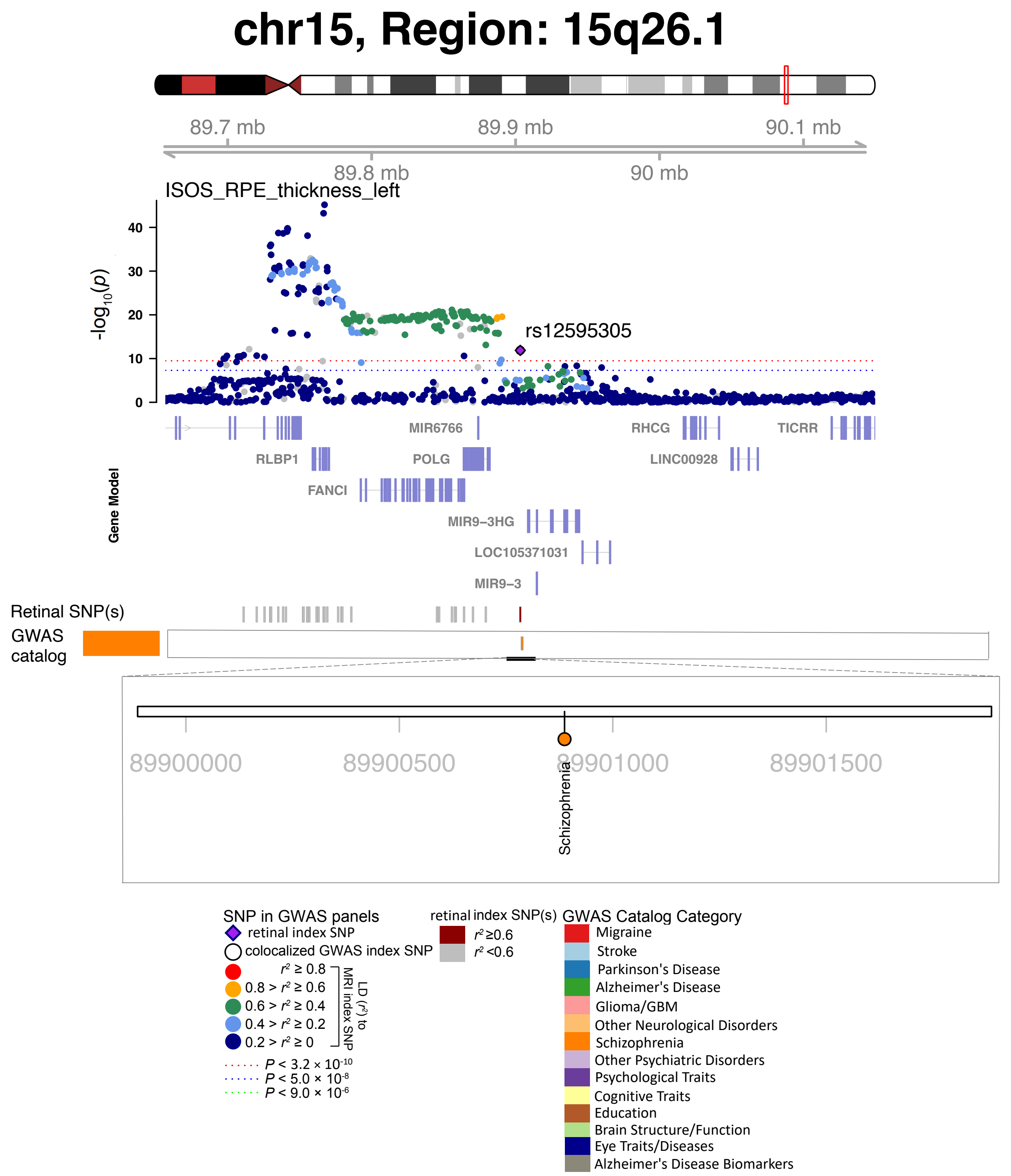


**Fig. S39 Selected genetic loci associated with retinal imaging traits and brain-related complex traits and diseases.** In 15q26.1, we observed the shared association (LD $r^2 \geq 0.6$) between the retinal imaging trait (ISOS_RPE_thickness_left) and schizophrenia (shared index variant rs12595305).

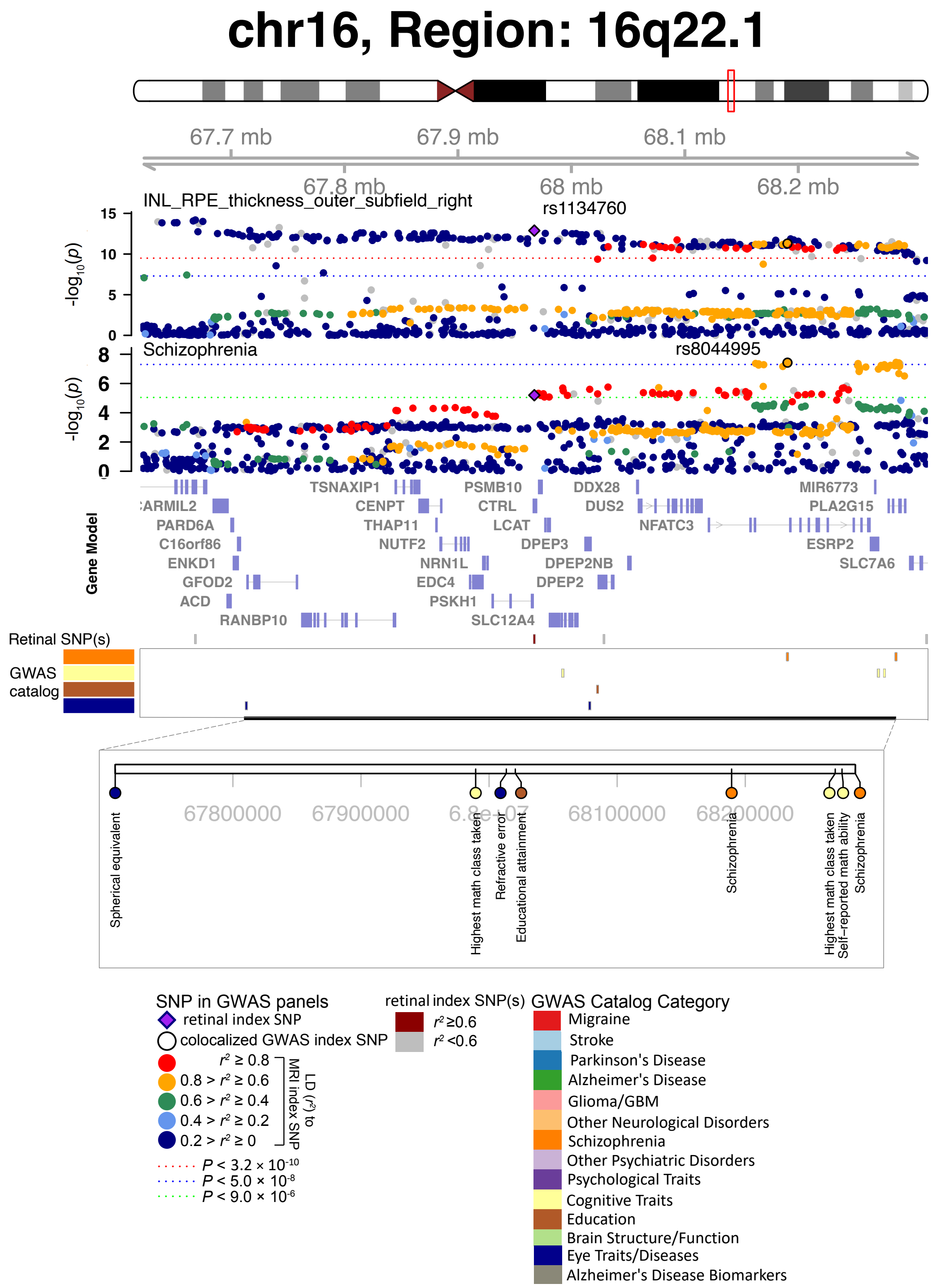


**Fig. S40 Selected genetic loci associated with retinal imaging traits and brain-related complex traits and diseases.** In 16q22.1, we observed the shared association (LD $r^2 \geq 0.6$) between the retinal imaging trait (INL_RPE_thickness_outer_subfield_right, index variant rs1134760) and schizophrenia (index variant rs8044995). We also observed shared associations with the highest math class taken and educational attainment.

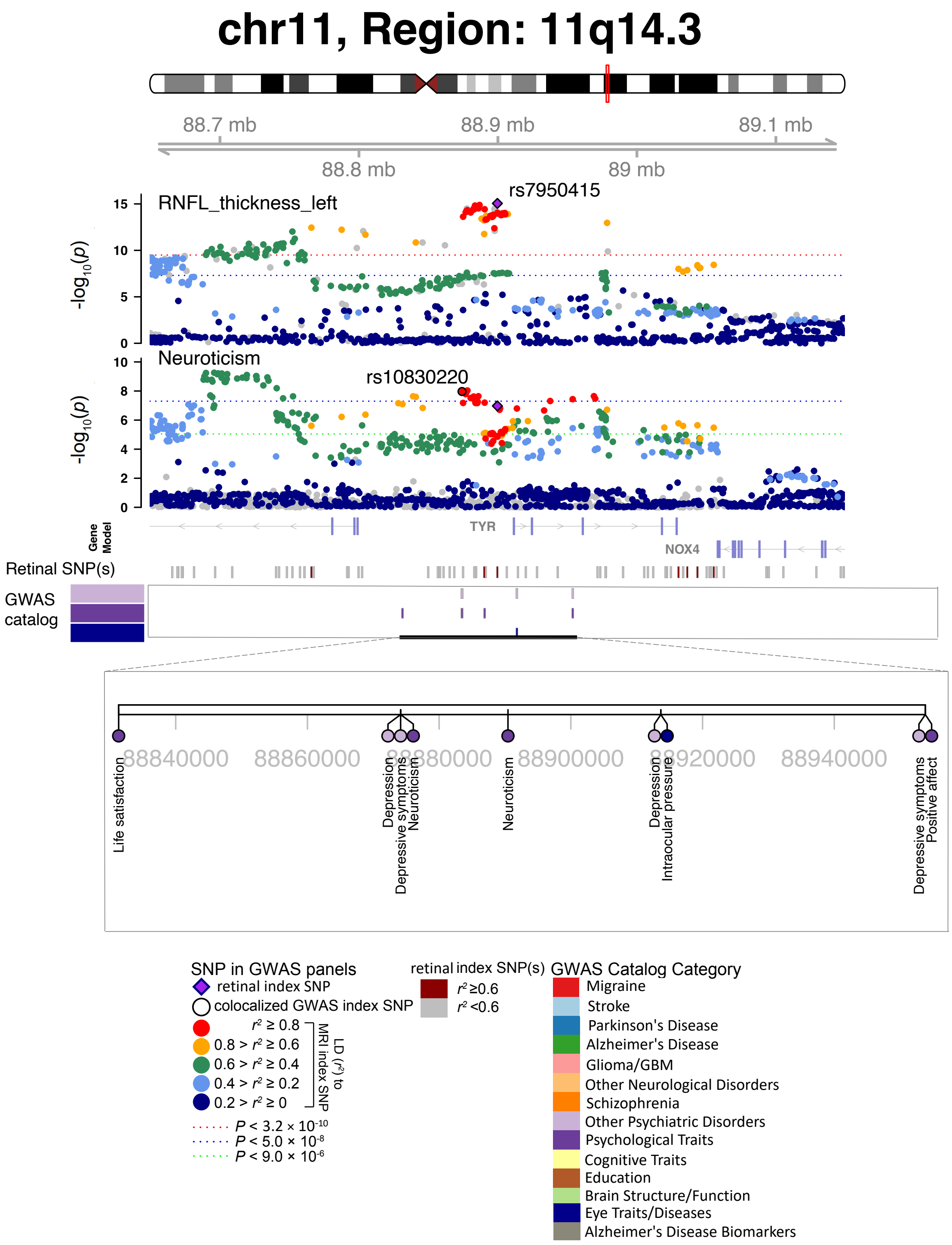


**Fig. S41 Selected genetic loci associated with retinal imaging traits and brain-related complex traits and diseases.** In 11q14.3, we observed the shared association (LD $r^2 \geq 0.6$) between the retinal imaging trait (RNFL_thickness_left, index variant rs7950415) and neuroticism (index variant rs10830220). We also observed shared associations with depression, depressive symptoms, life satisfaction, and positive affect.

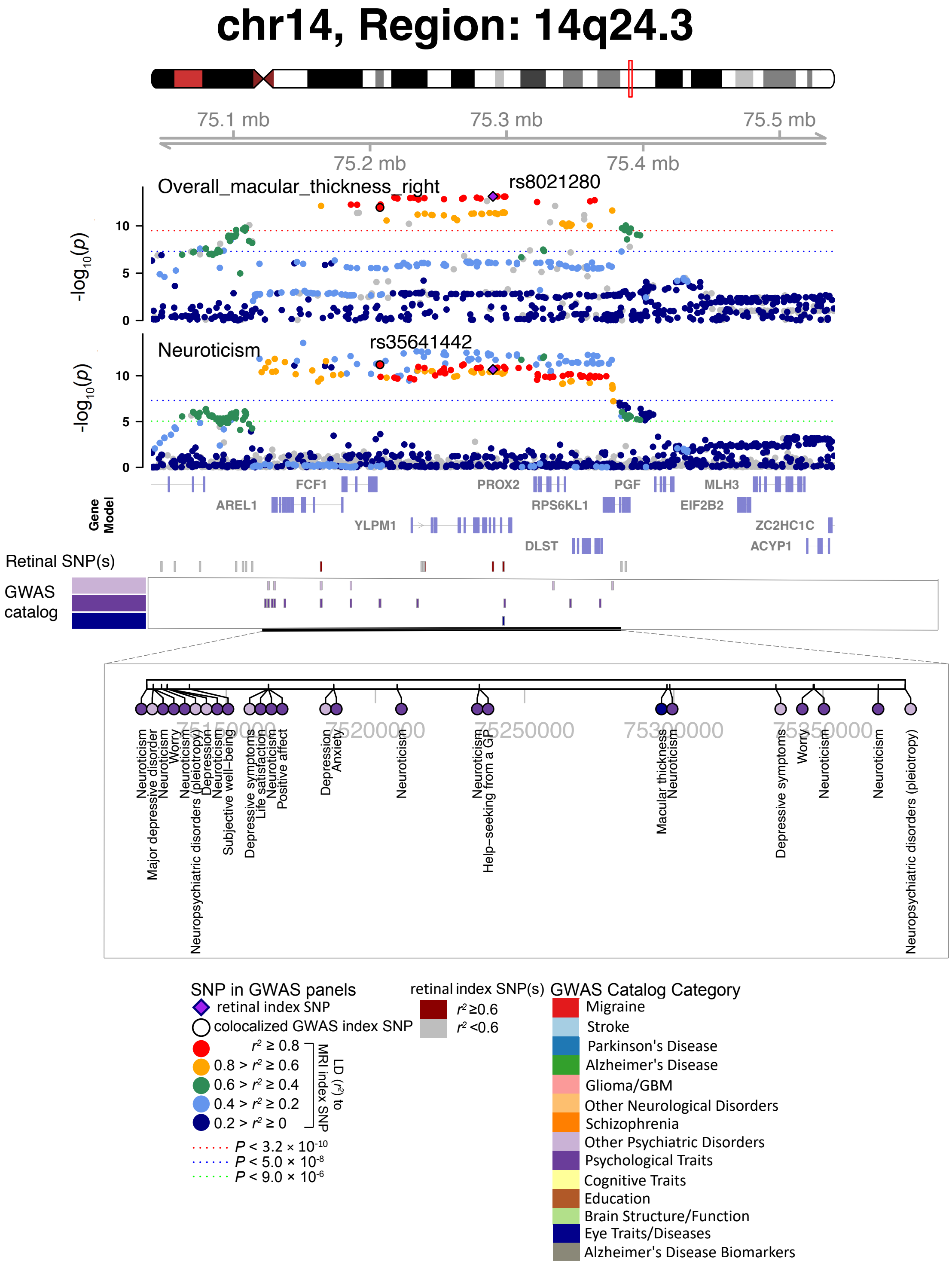


**Fig. S42 Selected genetic loci associated with retinal imaging traits and brain-related complex traits and diseases.** In 11q14.3, we observed the shared association (LD $r^2 \geq 0.6$) between the retinal imaging trait (overall_macular_thickness_right, index variant rs8021280) and neuroticism (index variant rs35641442). We also observed shared associations with major depressive disorder, depression, anxiety, life satisfaction, and psychiatric disorders (pleiotropy).

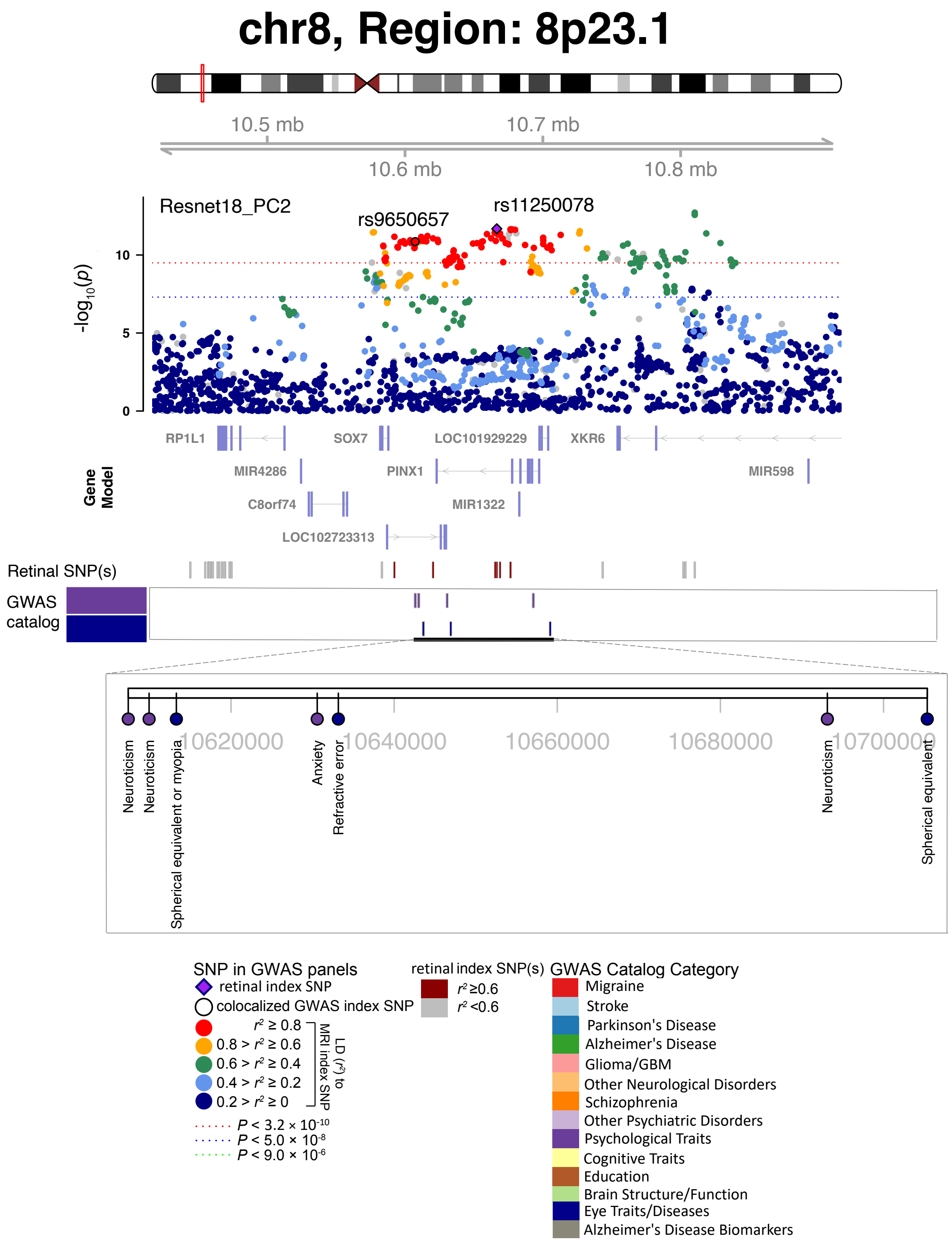


**Fig. S43 Selected genetic loci associated with retinal imaging traits and brain-related complex traits and diseases.** In 8p23.1, we observed the shared association (LD $r^2 \geq 0.6$) between the retinal imaging trait (Resnet18_PC2, index variant rs11250078) and neuroticism (index variant rs9650657). We also observed shared associations with anxiety.

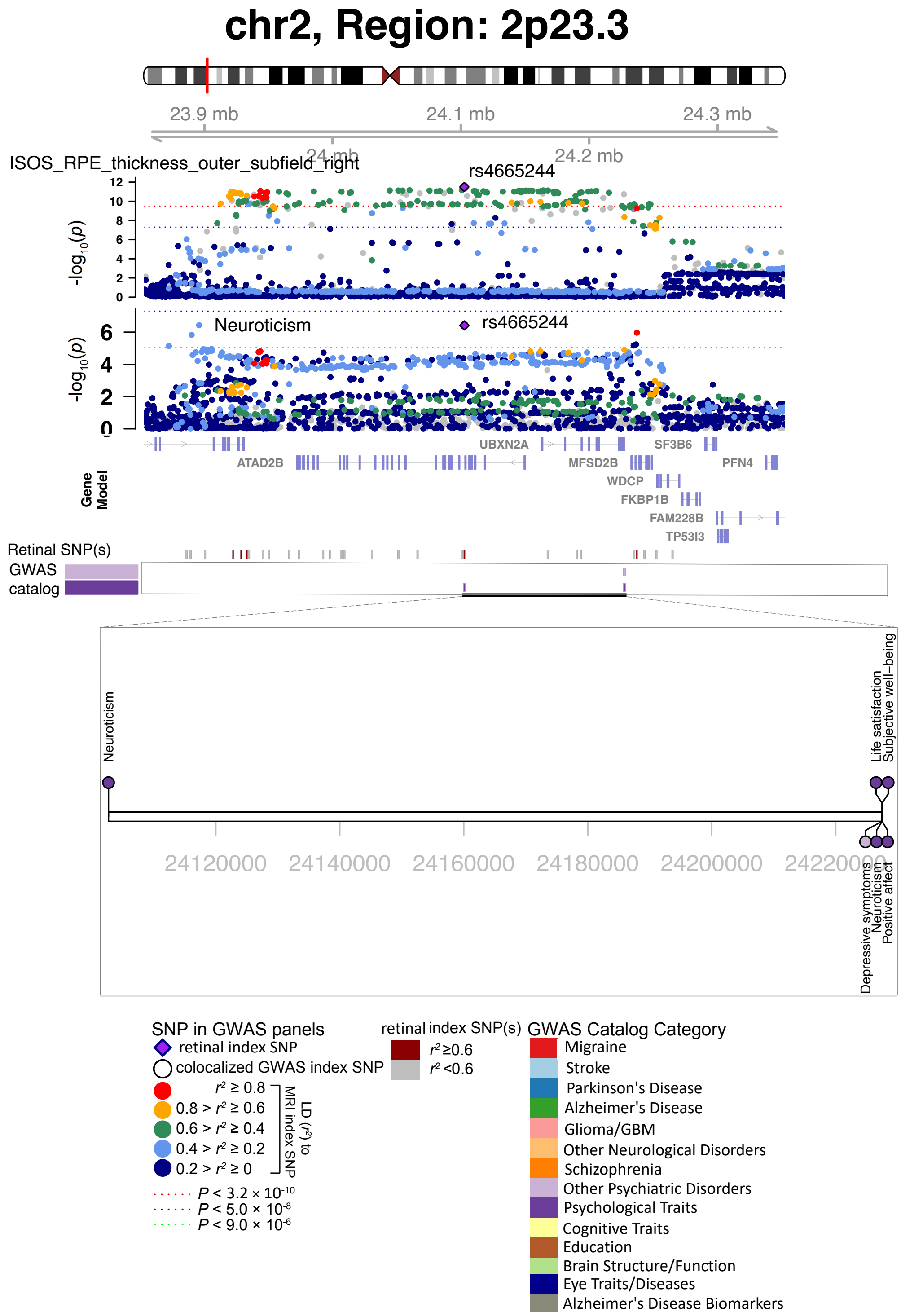


**Fig. S44 Selected genetic loci associated with retinal imaging traits and brain-related complex traits and diseases.** In 2p23.3, we observed the shared association (LD $r^2 \geq 0.6$) between the retinal imaging trait (ISOS_RPE_thickness_outer_subfield_right) and neuroticism (shared index variant rs4665244). We also observed shared associations with depressive symptoms, life satisfaction, positive affect, and well-being spectrum.

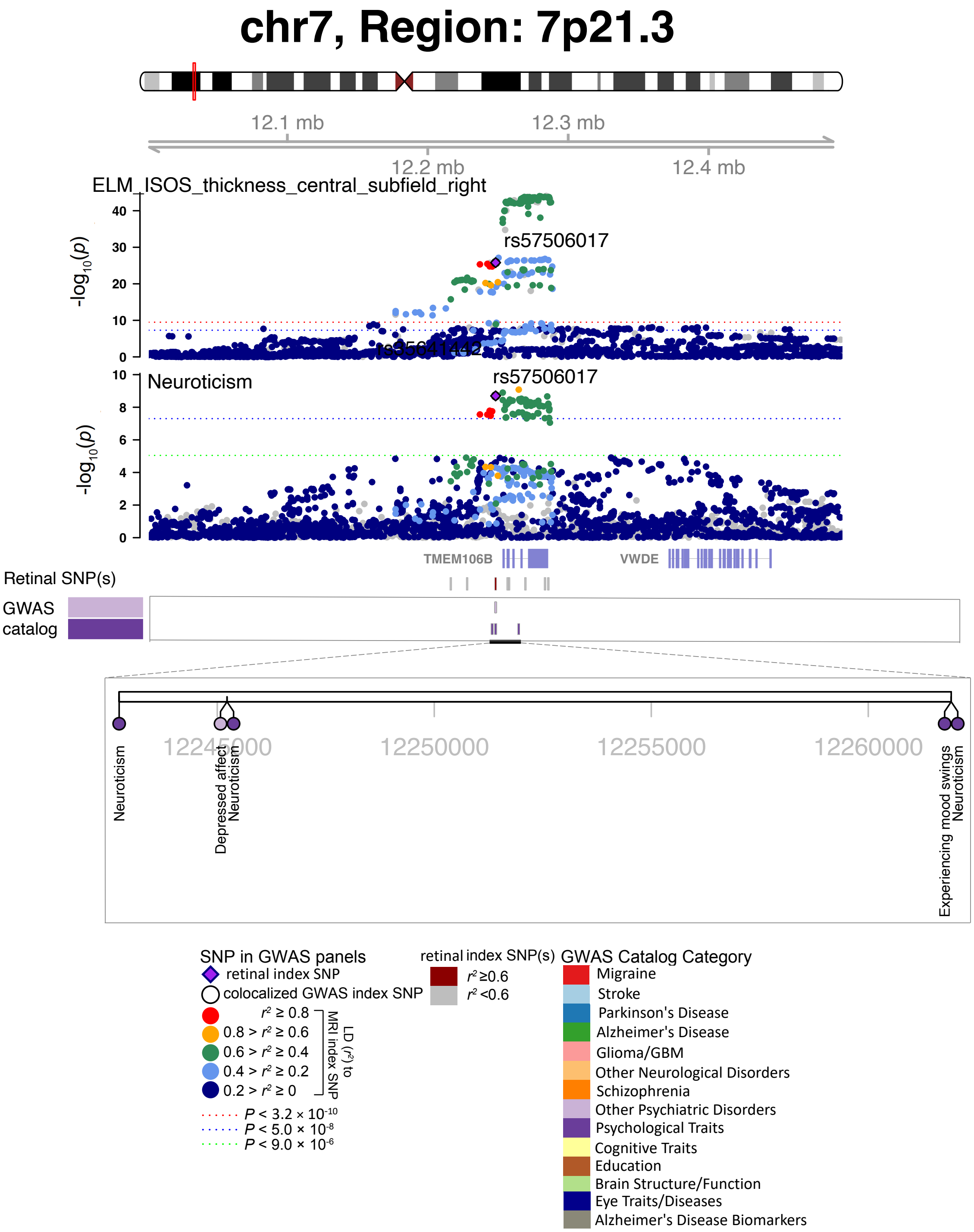


**Fig. S45 Selected genetic loci associated with retinal imaging traits and brain-related complex traits and diseases.** In 7p21.3, we observed the shared association (LD $r^2 \geq 0.6$) between the retinal imaging trait (ELM_ISOS_thickness_central_subfield_right) and neuroticism (shared index variant rs57506017). We also observed shared associations with depressed affect and experiencing mood swings.

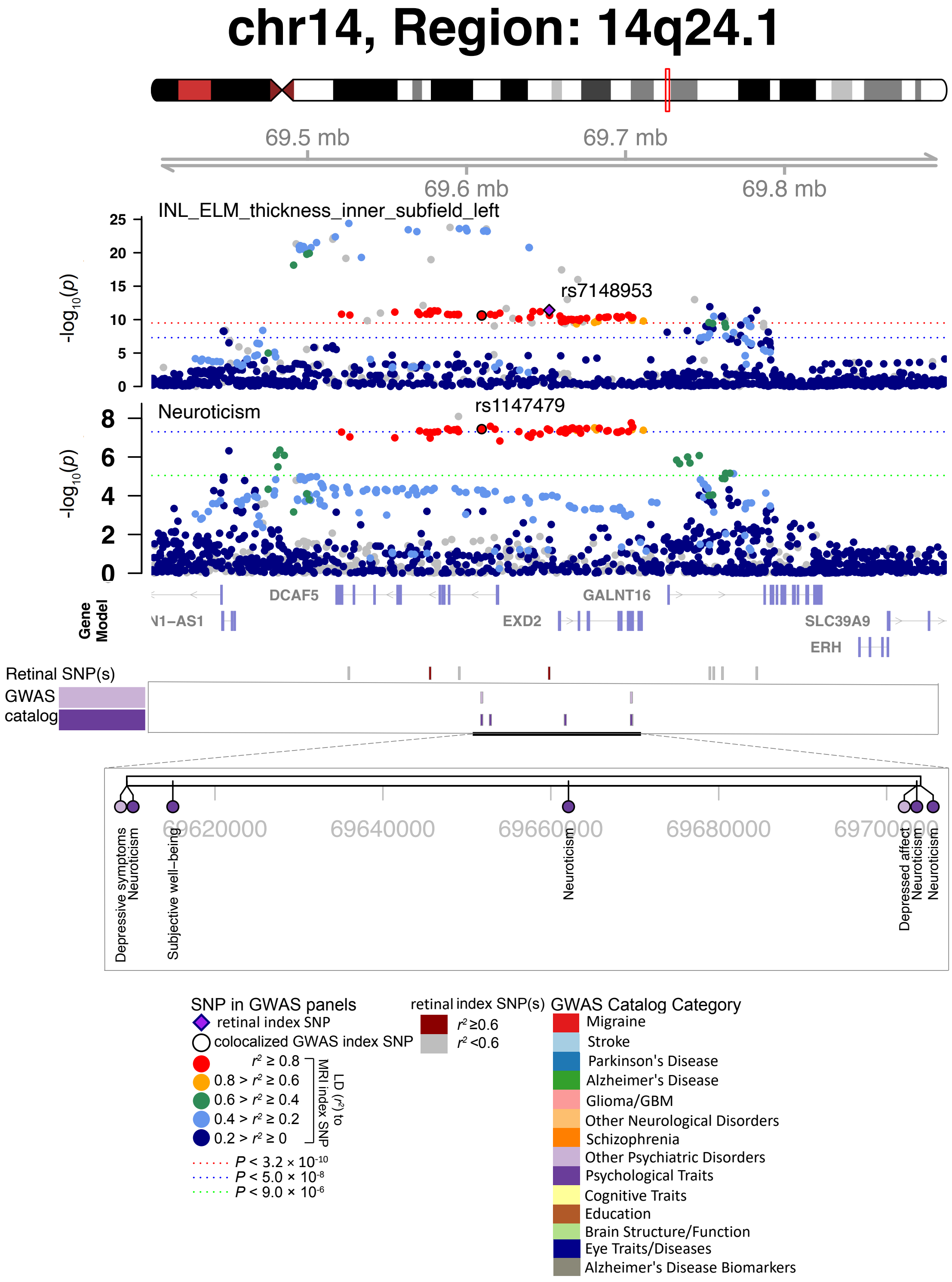


**Fig. S46 Selected genetic loci associated with retinal imaging traits and brain-related complex traits and diseases.** In 14q24.1, we observed the shared association (LD $r^2 \geq 0.6$) between the retinal imaging trait (INL_ELM_thickness_inner_subfield_left, index variant rs7148953) and neuroticism (index variant rs1147479). We also observed shared associations with depressed affect, depressive symptoms, and well-being spectrum.

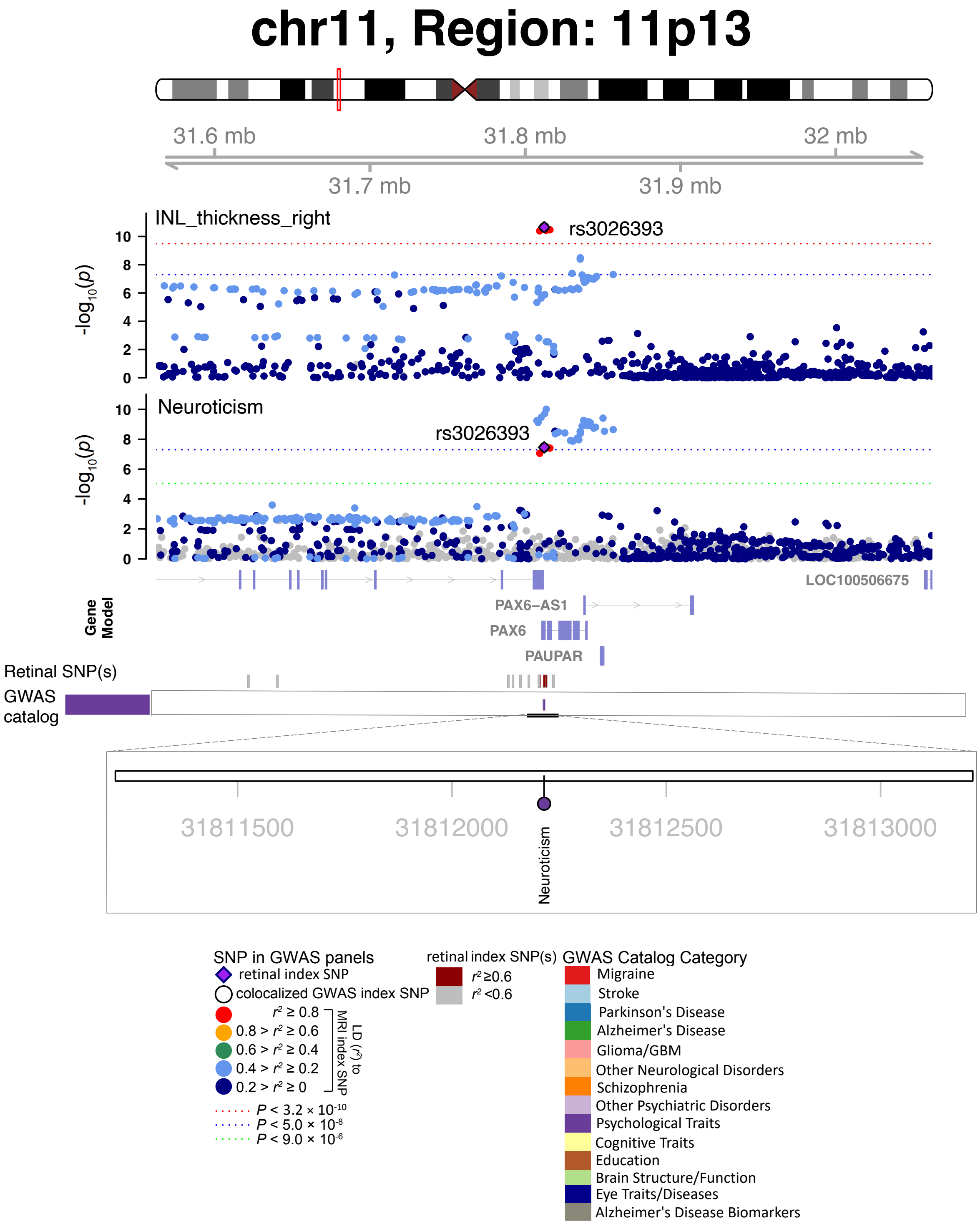


**Fig. S47 Selected genetic loci associated with retinal imaging traits and brain-related complex traits and diseases.** In 11p13, we observed the shared association (LD $r^2 \geq 0.6$) between the retinal imaging trait (INL_thickness_right) and neuroticism (shared index variant rs3026393).

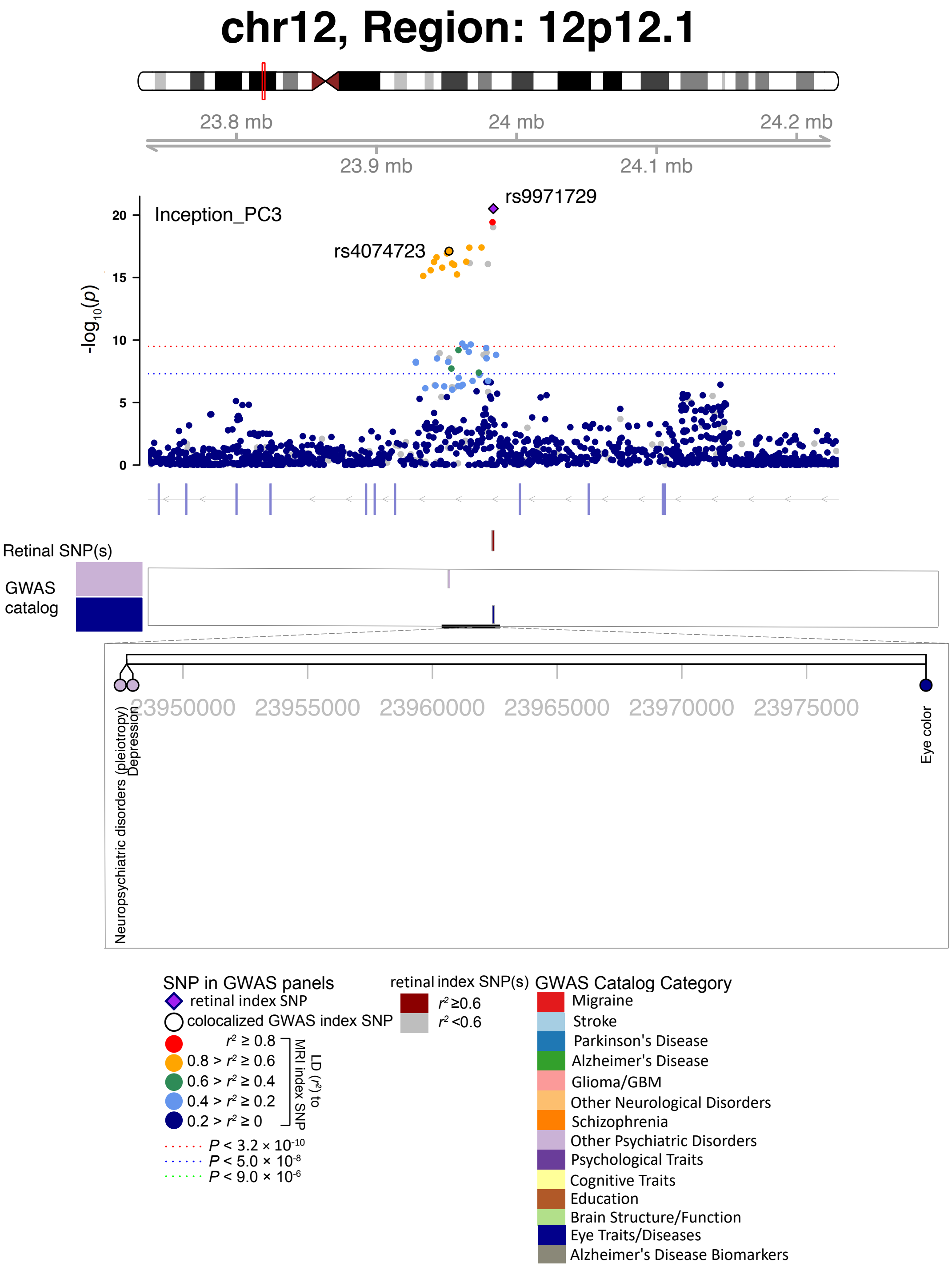


**Fig. S48 Selected genetic loci associated with retinal imaging traits and brain-related complex traits and diseases.** In 12p12.1, we observed the shared association (LD $r^2 \geq 0.6$) between the retinal imaging trait (Inception_PC3, index variant rs9971729) and depression (index variant rs4074723). We also observed shared associations with bipolar disorder or major depressive disorder.

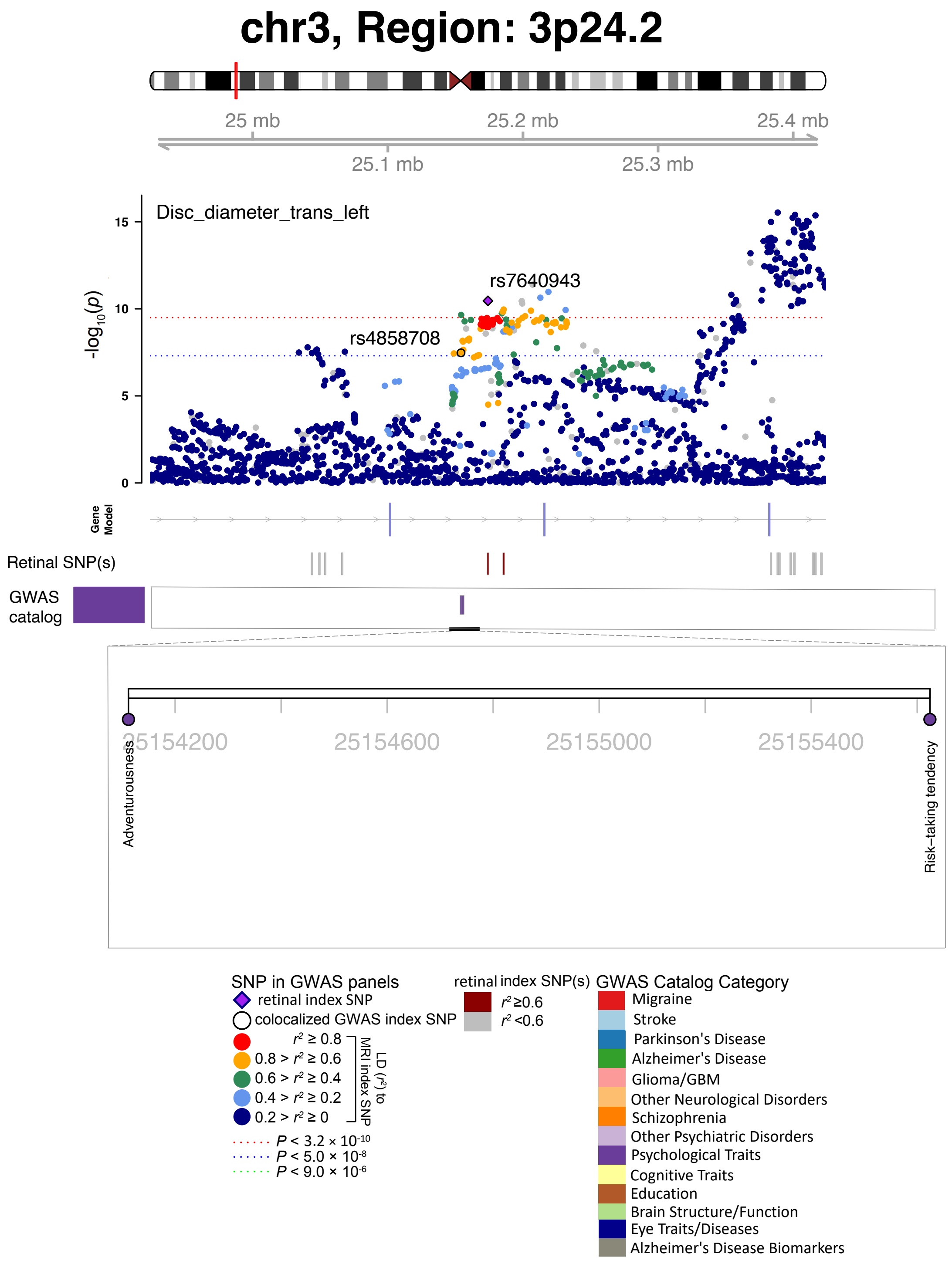


**Fig. S49 Selected genetic loci associated with retinal imaging traits and brain-related complex traits and diseases.** In 3p24.2, we observed the shared association (LD $r^2 \geq 0.6$) between the retinal imaging trait (Disc_diameter_trans_left, index variant rs7640943) and adventurousness (index variant rs4858708). We also observed shared associations with risk tolerance.

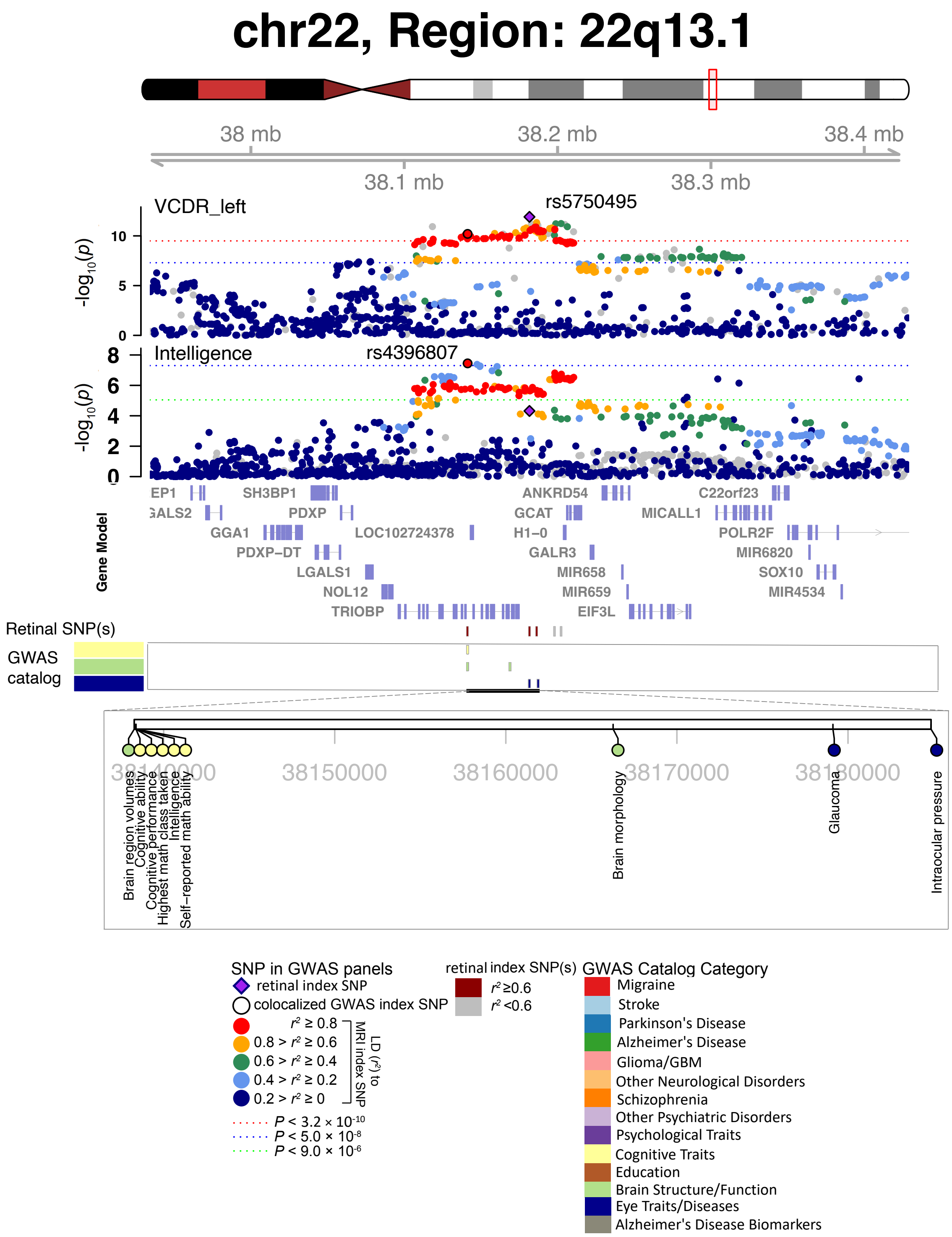


**Fig. S50 Selected genetic loci associated with retinal imaging traits and brain-related complex traits and diseases.** In 22q13.1, we observed the shared association (LD $r^2 \geq 0.6$) between the retinal imaging trait (VCDR_left, index variant rs5750495) and intelligence (index variant rs4396807).

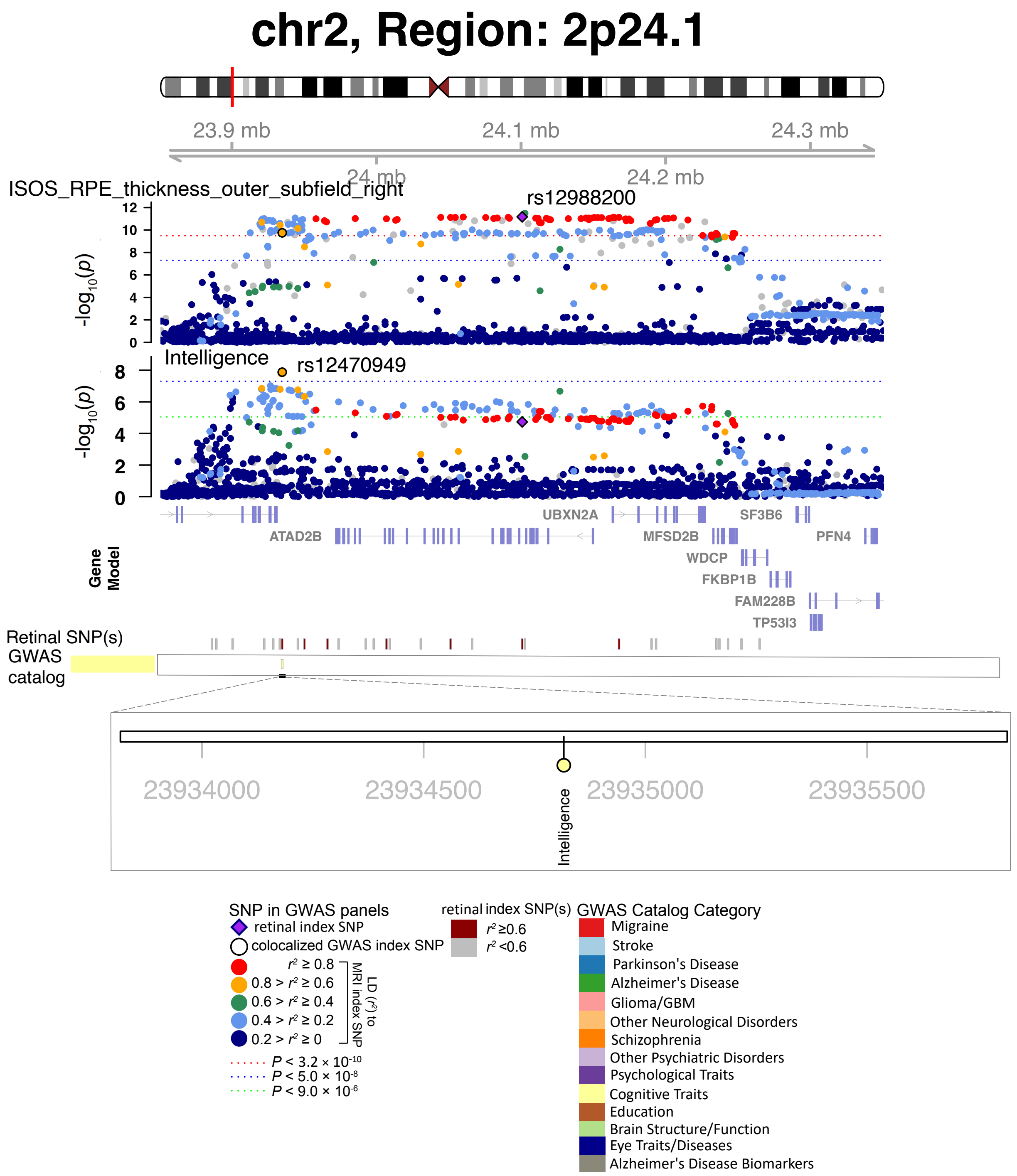


**Fig. S51 Selected genetic loci associated with retinal imaging traits and brain-related complex traits and diseases.** In 2p24.1, we observed the shared association (LD $r^2 \geq 0.6$) between the retinal imaging trait (ISOS_RPE_thickness_outer_subfield_right, index variant rs12988200) and intelligence (index variant rs12470949).

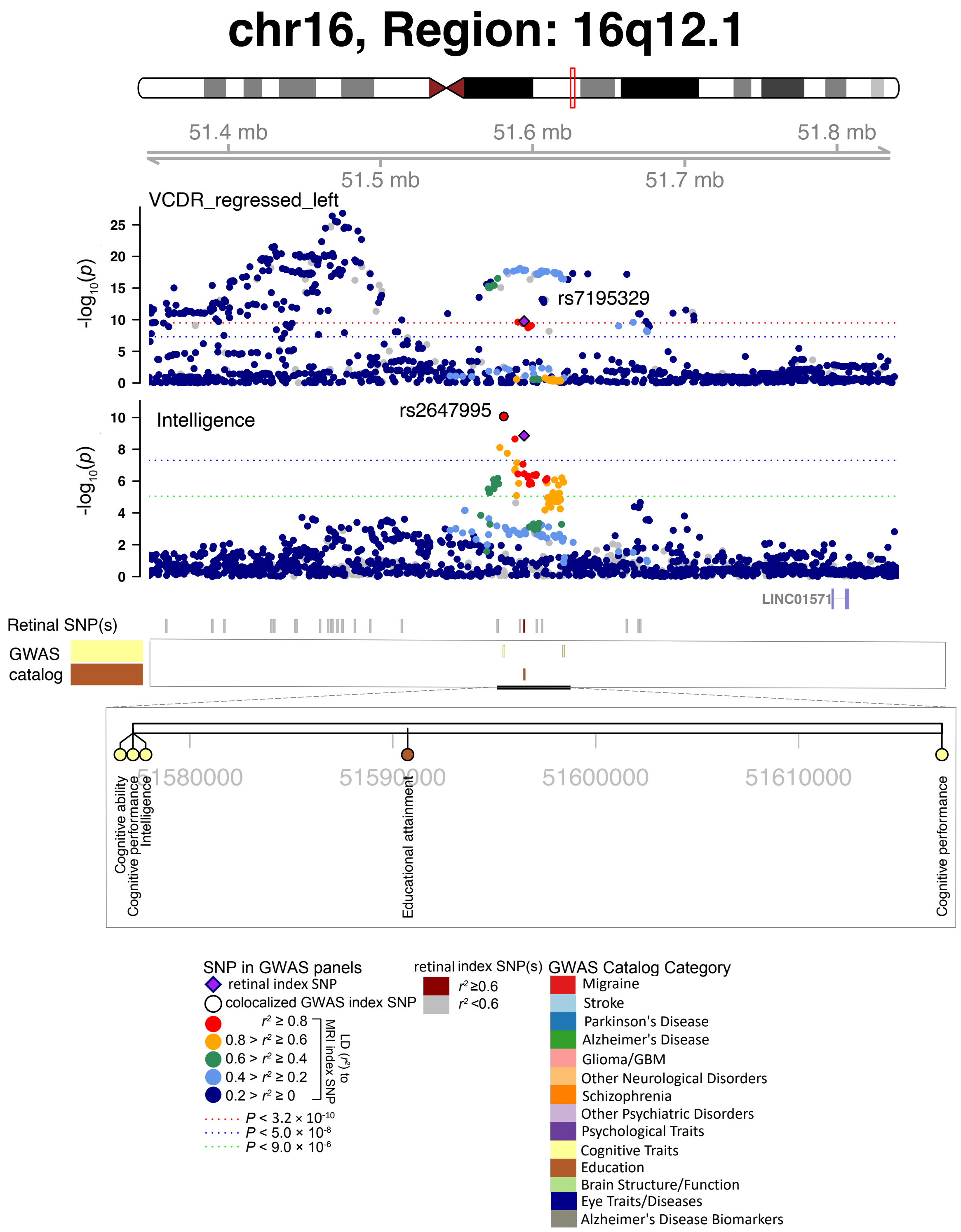


**Fig. S52 Selected genetic loci associated with retinal imaging traits and brain-related complex traits and diseases.** In 16q12.1, we observed the shared association (LD $r^2 \geq 0.6$) between the retinal imaging trait (VCDR_regressed_left, index variant rs7195329) and intelligence (index variant rs2647995). We also observed shared associations with educational attainment.

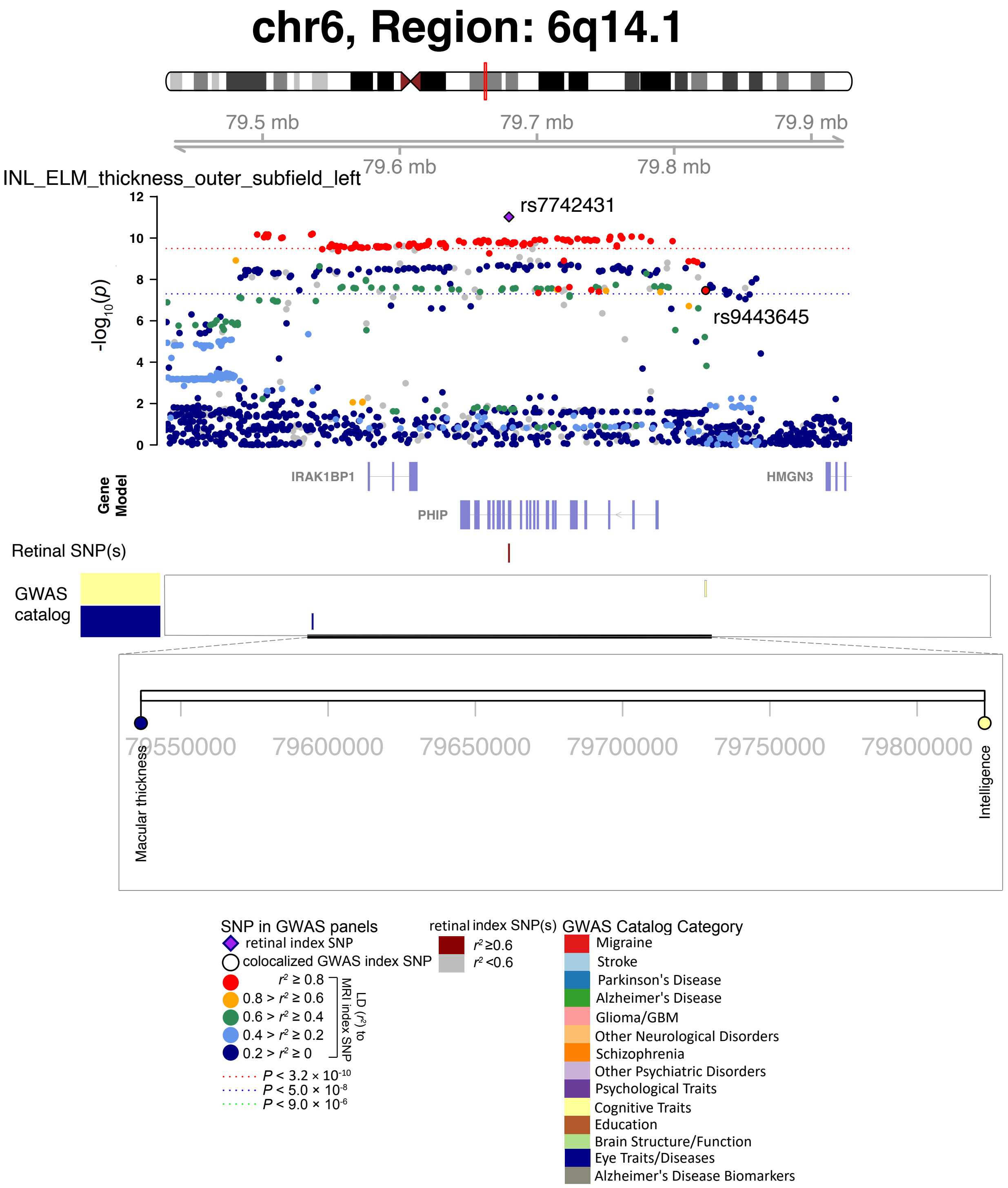


**Fig. S53 Selected genetic loci associated with retinal imaging traits and brain-related complex traits and diseases.** In 6q14.1, we observed the shared association (LD $r^2 \geq 0.6$) between the retinal imaging trait (INL_ELM_thickness_outer_subfield_left, index variant rs7742431) and intelligence (index variant rs9443645).

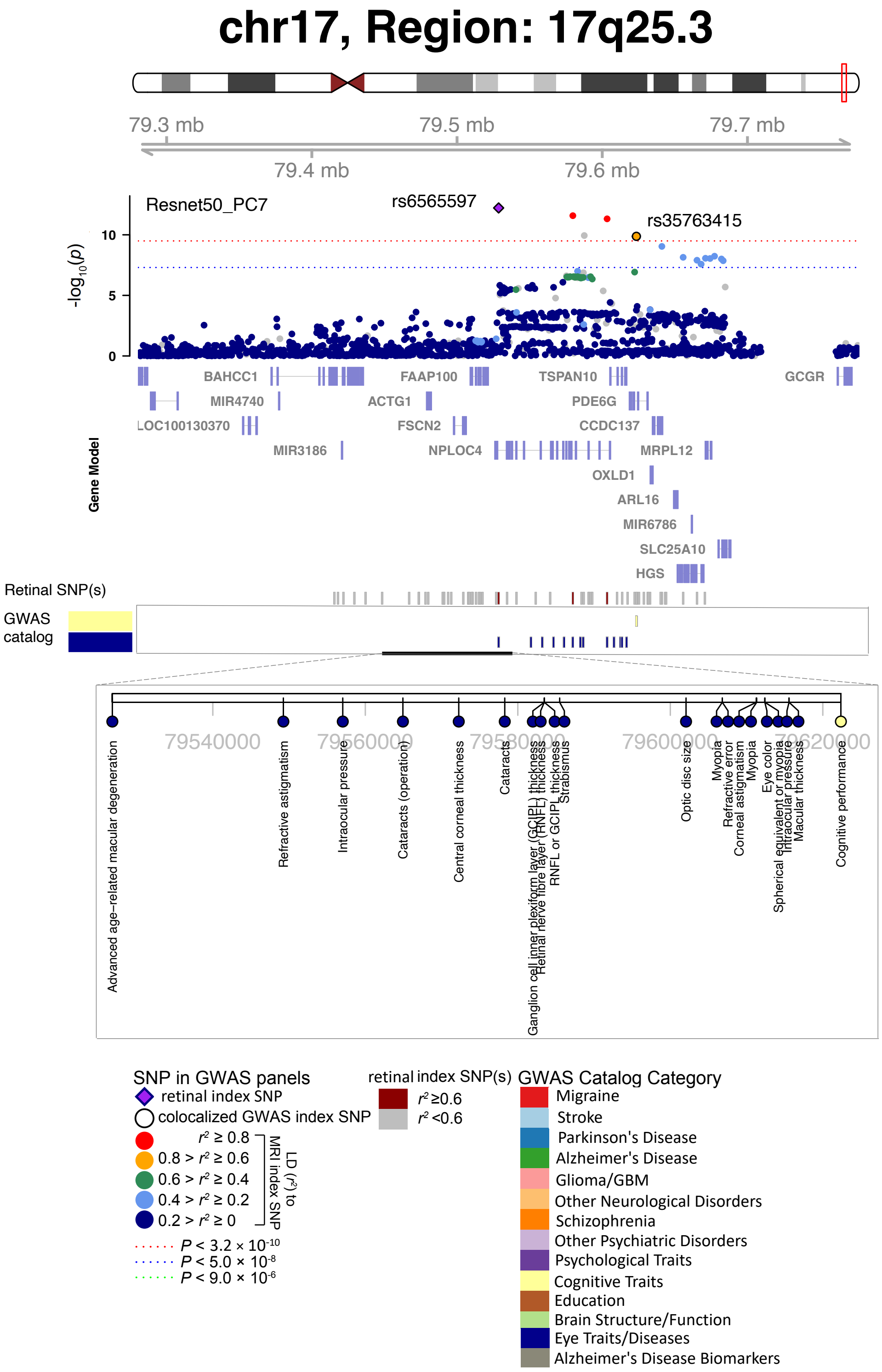


**Fig. S54 Selected genetic loci associated with retinal imaging traits and brain-related complex traits and diseases.** In 17q25.3, we observed the shared association (LD $r^2 \geq 0.6$) between the retinal imaging trait (Resnet50_PC7, index variant rs6565597) and cognitive performance (index variant rs35763415).

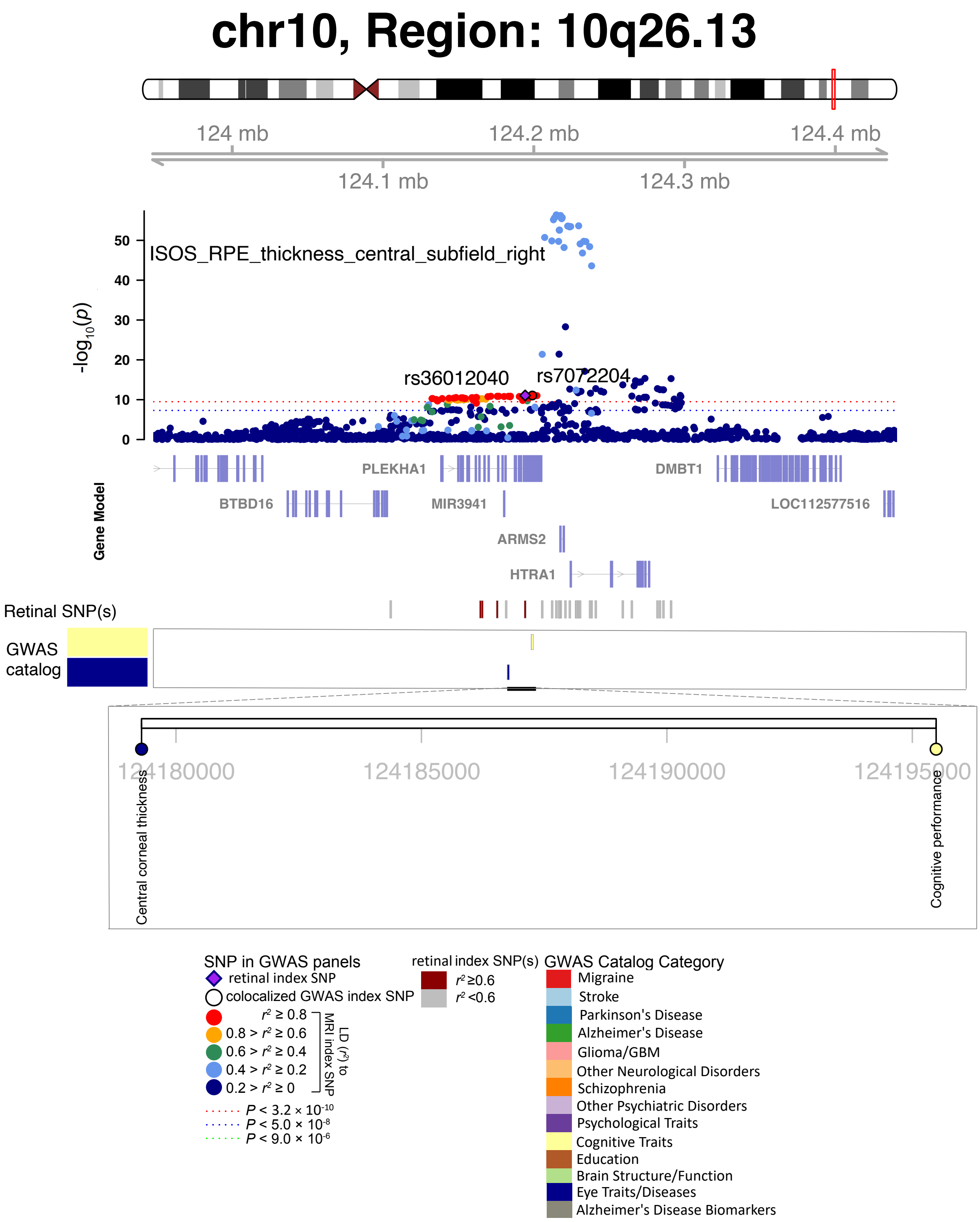


**Fig. S55 Selected genetic loci associated with retinal imaging traits and brain-related complex traits and diseases.** In 10q26.13, we observed the shared association (LD $r^2 \geq 0.6$) between the retinal imaging trait (ISOS_RPE_thickness_central_subfield_right, index variant rs36012040) and cognitive performance (index variant rs7072204).

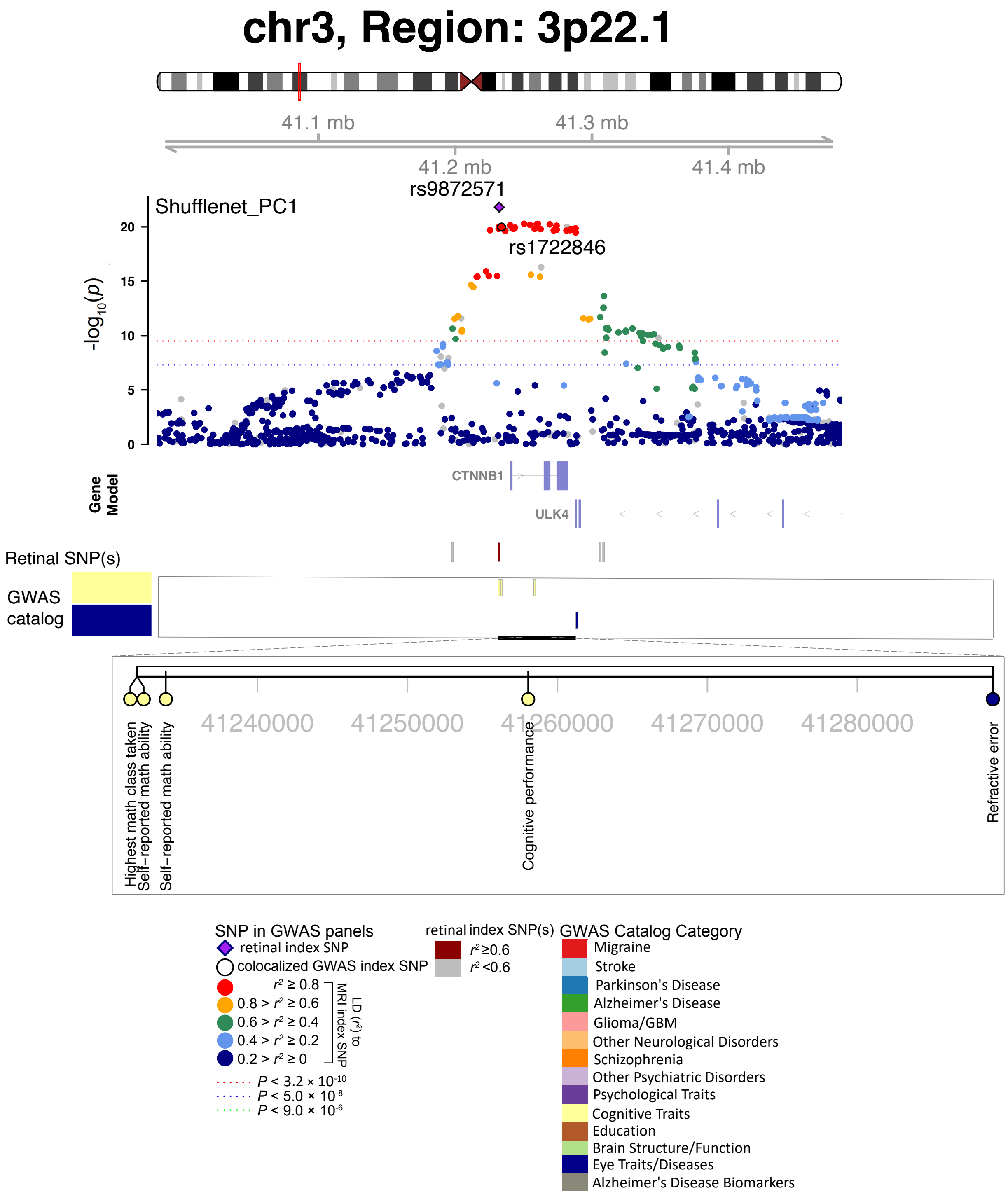


**Fig. S56 Selected genetic loci associated with retinal imaging traits and brain-related complex traits and diseases.** In 3p22.1, we observed the shared association (LD $r^2 \geq 0.6$) between the retinal imaging trait (Shufflenet_PC1, index variant rs9872571) and self-reported math ability (index variant rs1722846).

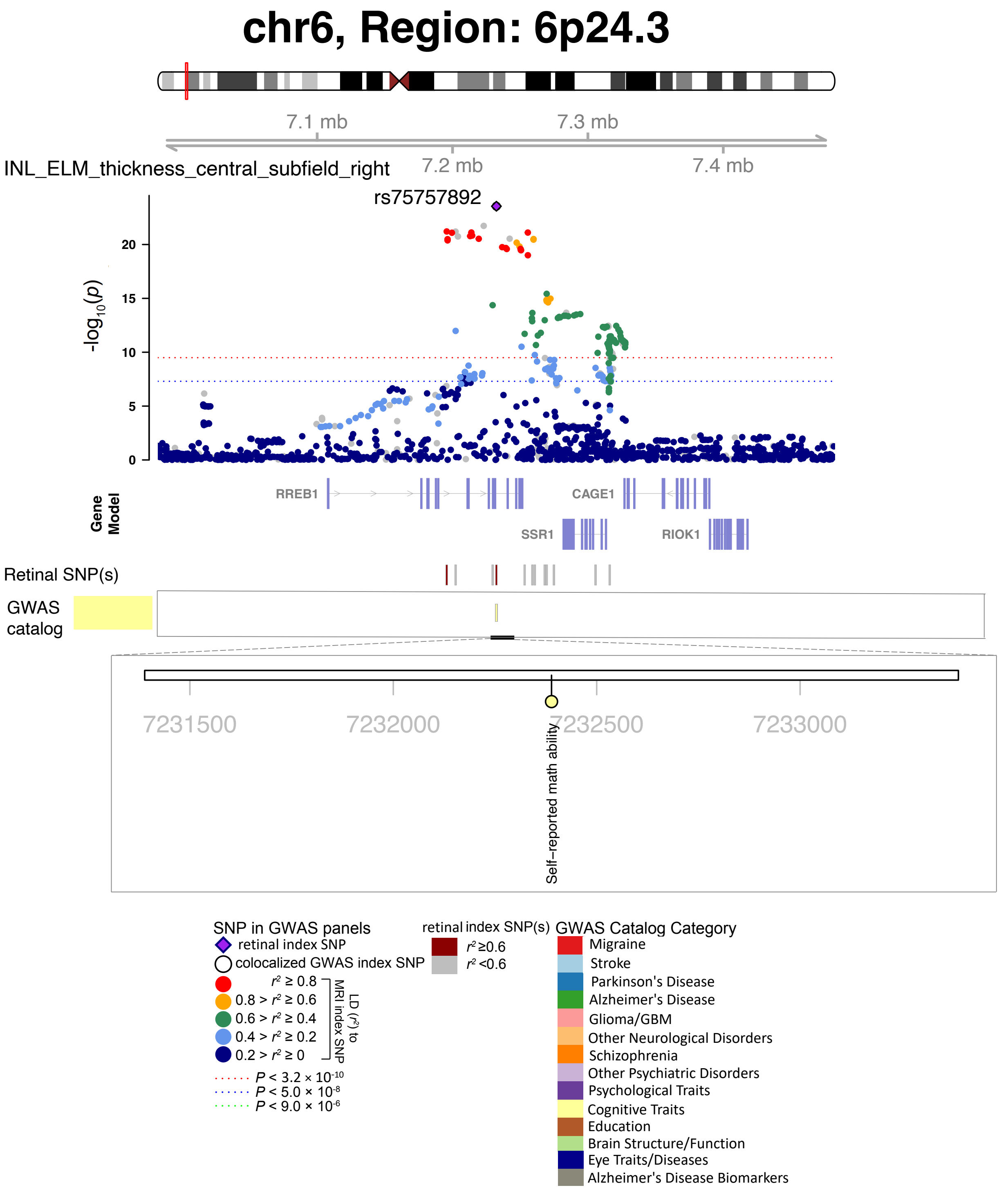


**Fig. S57 Selected genetic loci associated with retinal imaging traits and brain-related complex traits and diseases.** In 6p24.3, we observed the shared association (LD $r^2 \geq 0.6$) between the retinal imaging trait (INL_ELM_thickness_central_subfield_right) and self-reported math ability (shared index variant rs75757892).

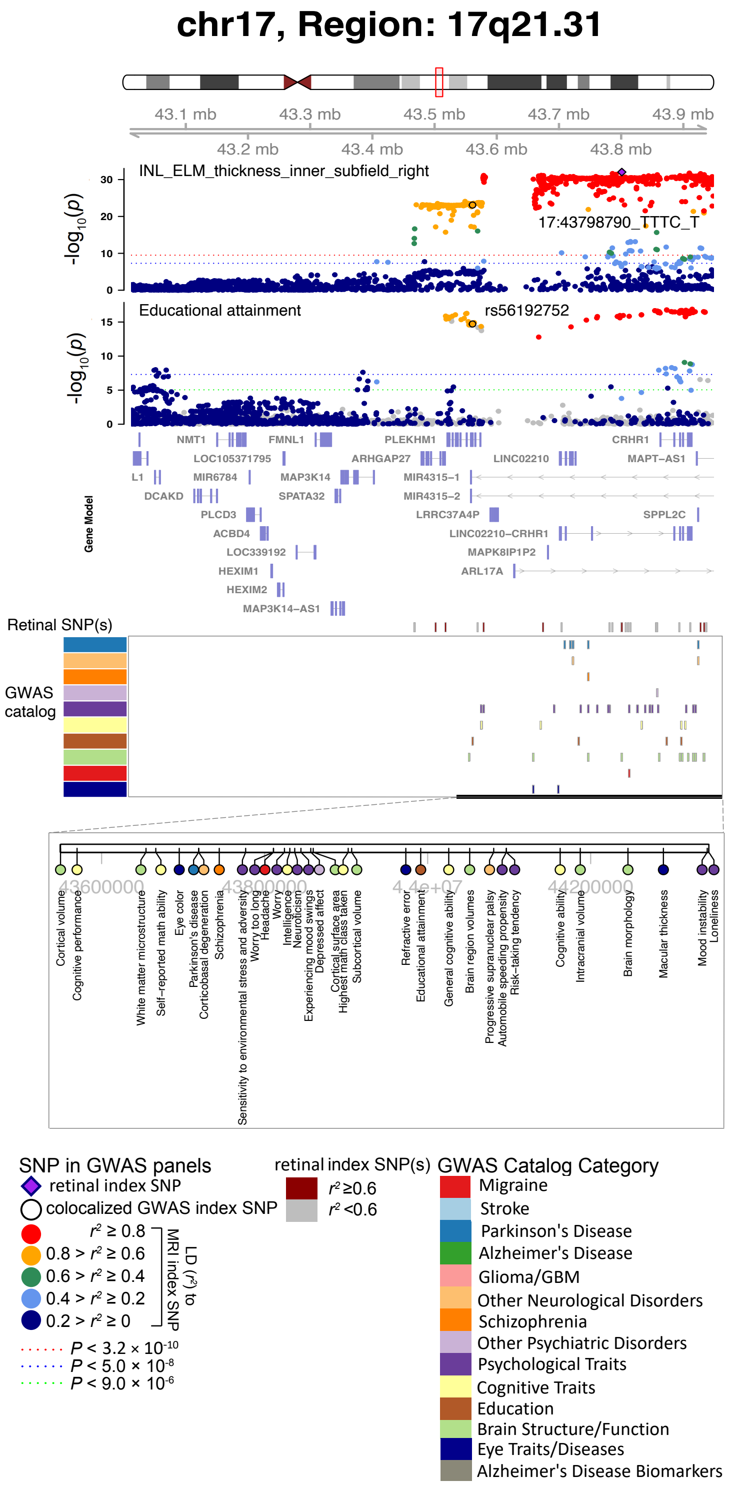


**Fig. S58 Selected genetic loci associated with retinal imaging traits and brain-related complex traits and diseases.** In 17q21.31, we observed the shared association (LD $r^2 \geq 0.6$) between the retinal imaging trait (INL_ELM_thickness_inner_subfield_right, index variant 17:43798790_TTTC_T) and educational attainment (index variant rs56192752).

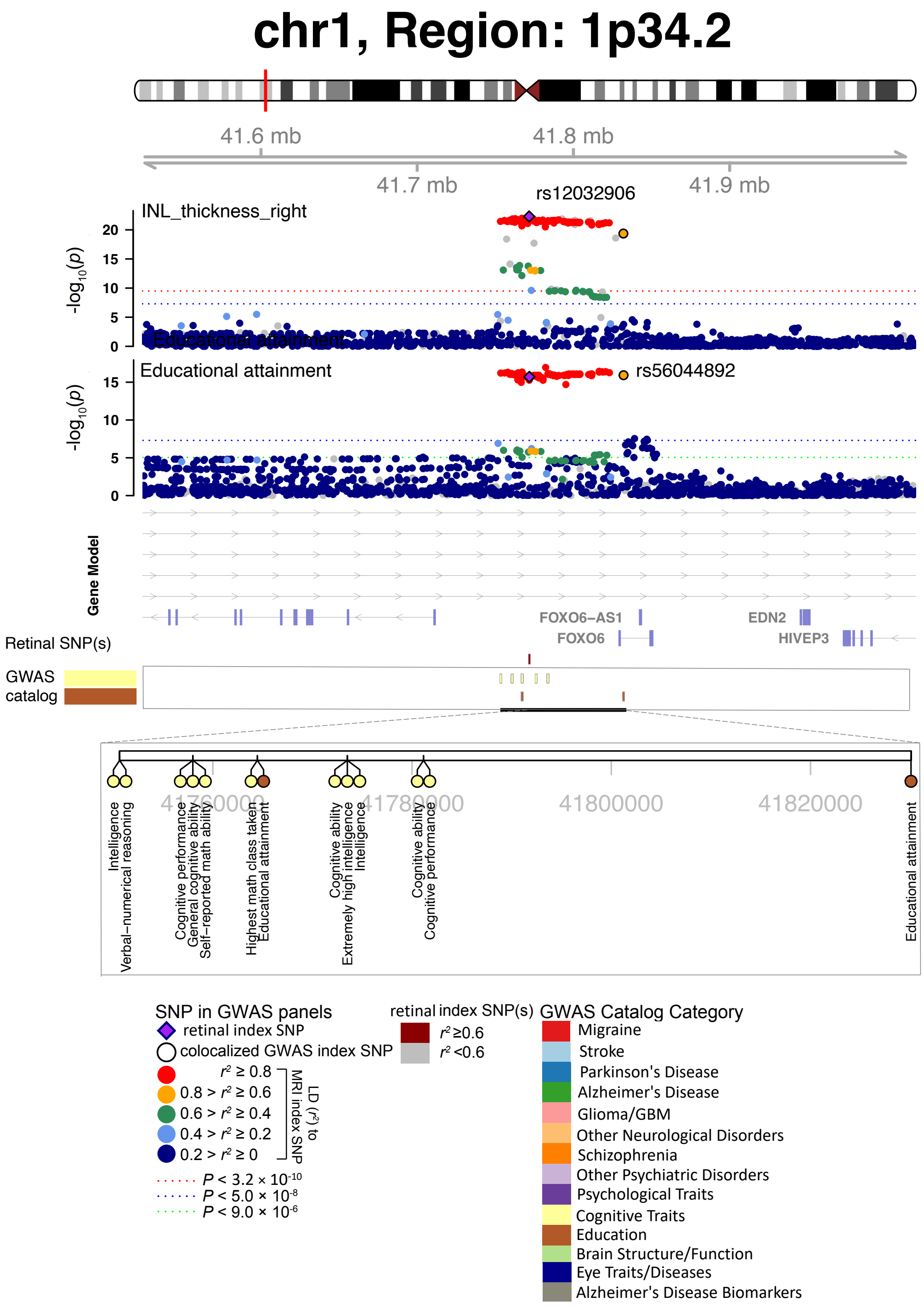


**Fig. S59 Selected genetic loci associated with retinal imaging traits and brain-related complex traits and diseases.** In 1p34.2, we observed the shared association (LD $r^2 \geq 0.6$) between the retinal imaging trait (INL_thickness_right, index variant rs12032906) and educational attainment (index variant rs56044892). We also observed shared associations with cognitive ability, cognitive performance, extremely high intelligence, general cognitive ability, highest math class taken, intelligence, and verbal-numerical reasoning.

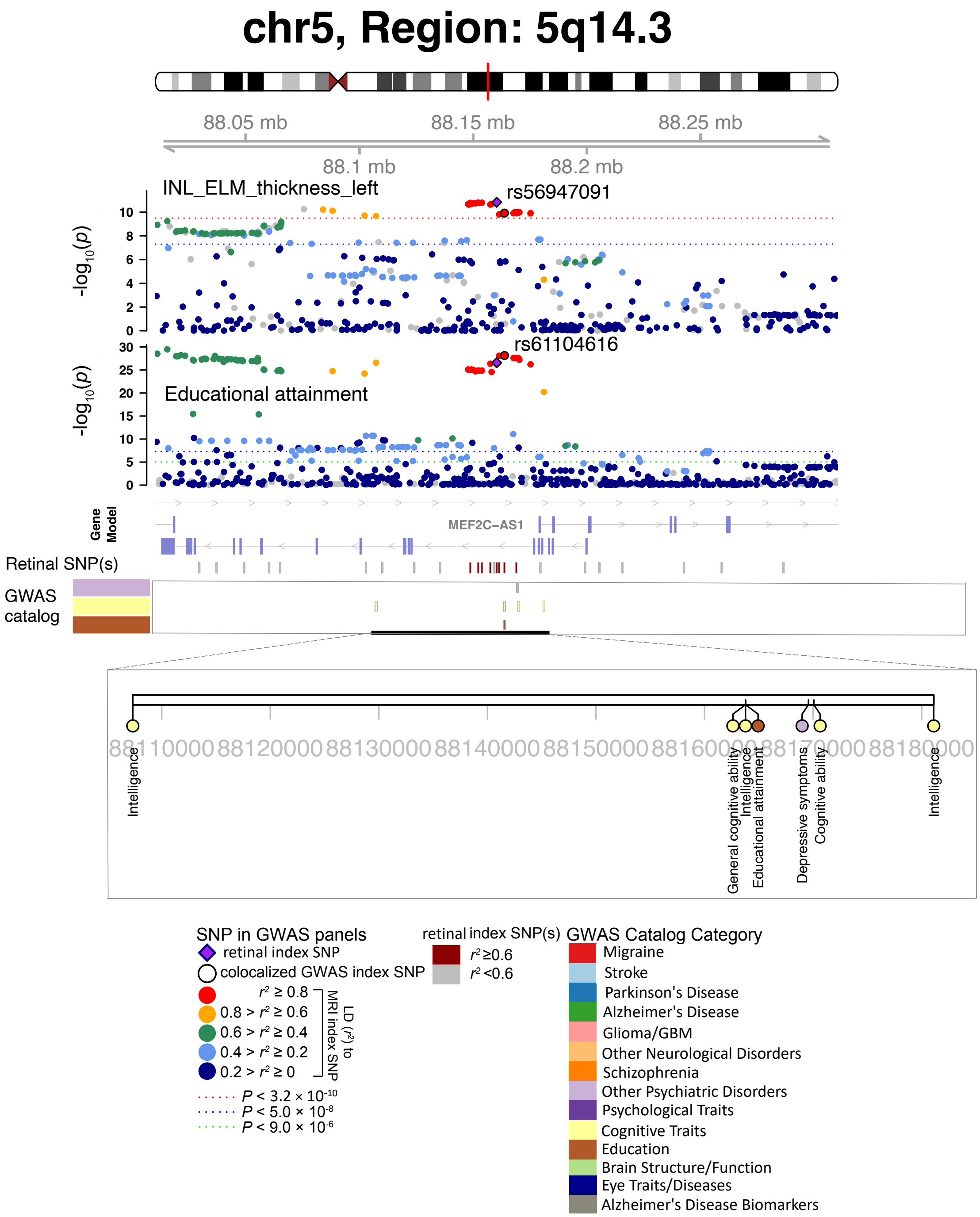


**Fig. S60 Selected genetic loci associated with retinal imaging traits and brain-related complex traits and diseases.** In 5q14.3, we observed the shared association (LD $r^2 \geq 0.6$) between the retinal imaging trait (INL_ELM_thickness_left, index variant rs56947091) and educational attainment (index variant rs61104616). We also observed shared associations with general cognitive ability, intelligence, and depressive symptoms.

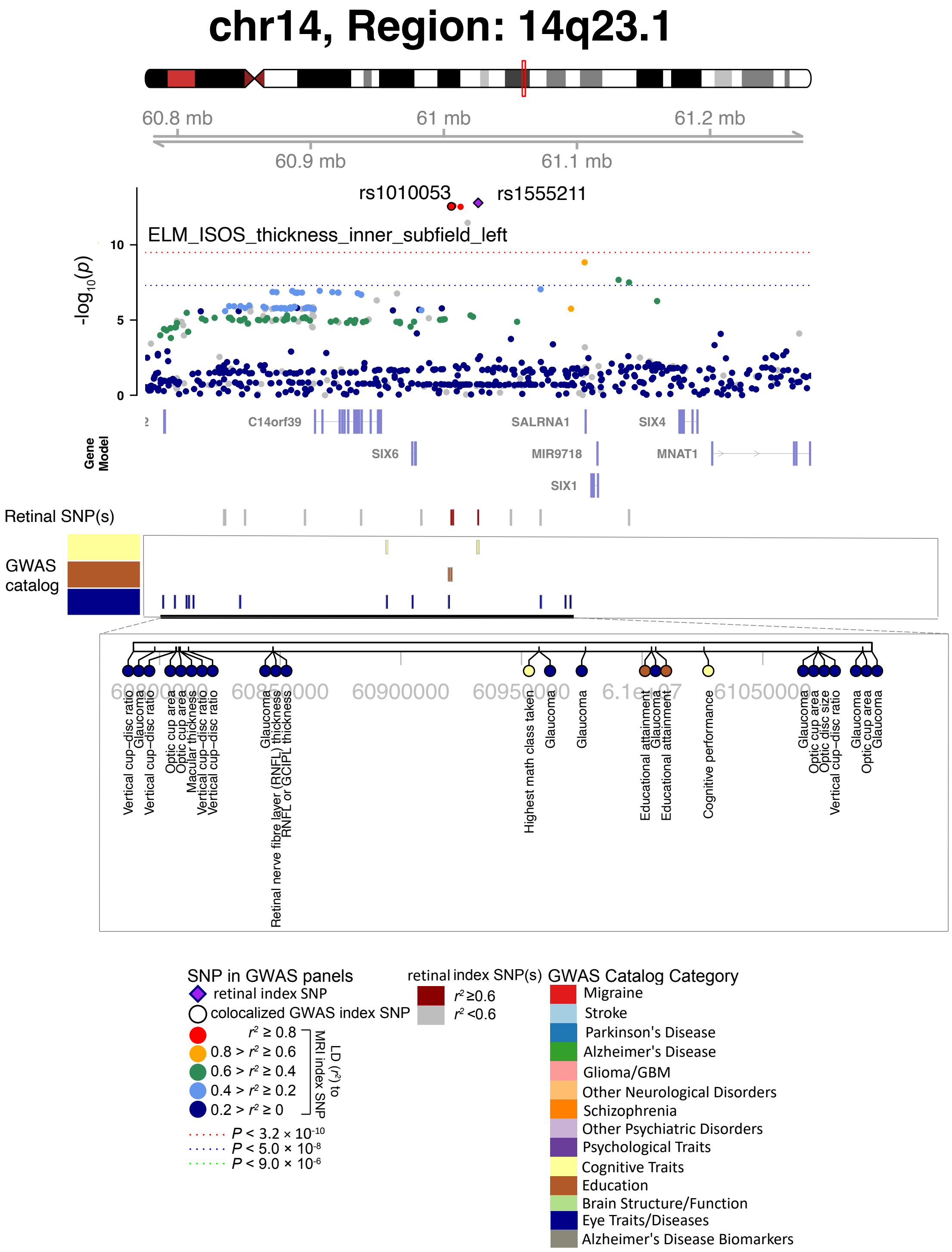


**Fig. S61 Selected genetic loci associated with retinal imaging traits and brain-related complex traits and diseases.** In 14q23.1, we observed the shared association (LD $r^2 \geq 0.6$) between the retinal imaging trait (ELM_ISOS_thickness_inner_subfield_left, index variant rs1555211) and educational attainment (index variant rs1010053). We also observed shared associations with cognitive performance and the highest math class taken.

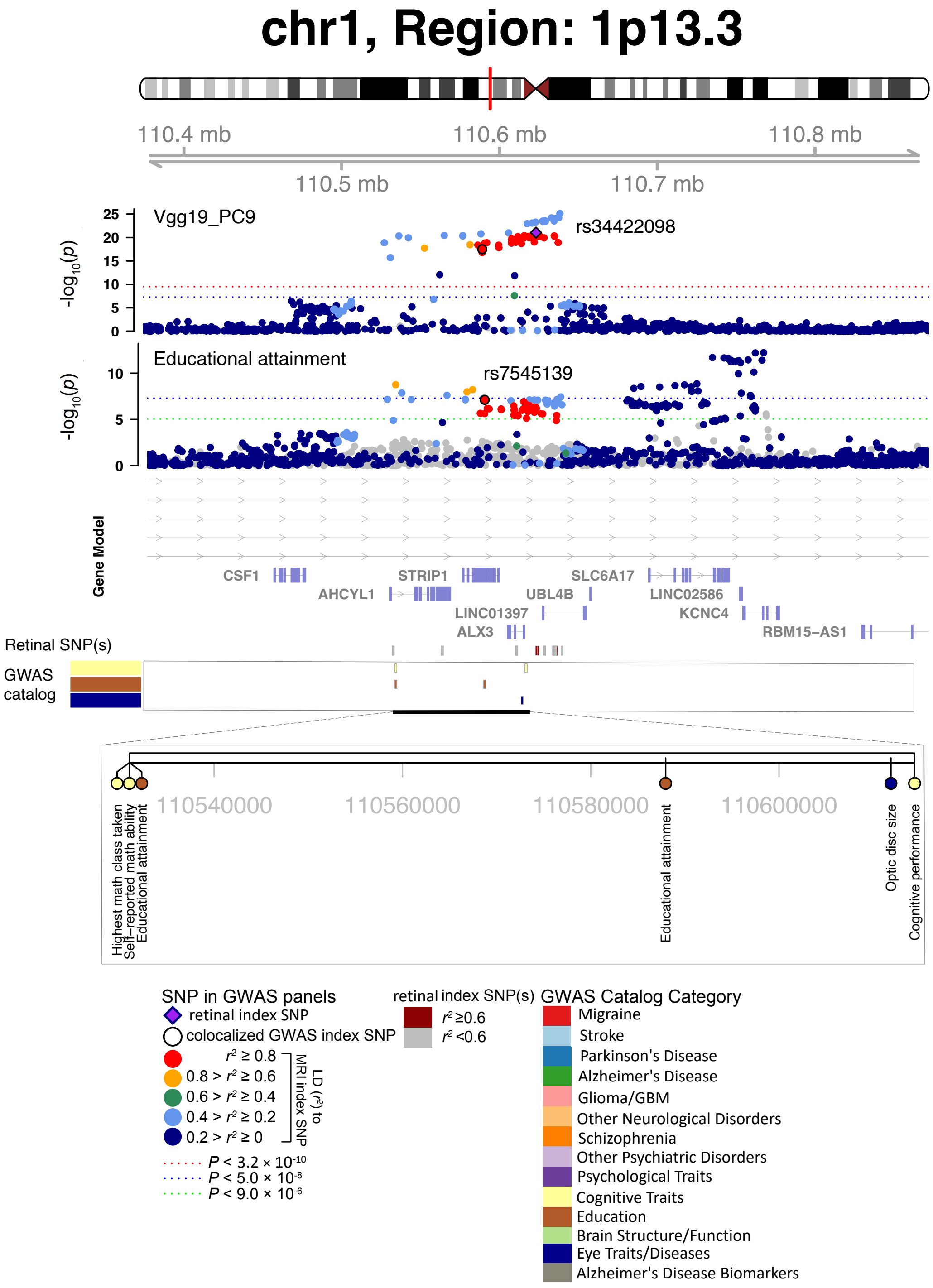


**Fig. S62 Selected genetic loci associated with retinal imaging traits and brain-related complex traits and diseases.** In 1p13.3, we observed the shared association (LD $r^2 \geq 0.6$) between the retinal imaging trait (Vgg19_PC9, index variant rs34422098) and educational attainment (index variant rs7545139). We also observed shared associations with self-reported math ability and the highest math class taken.

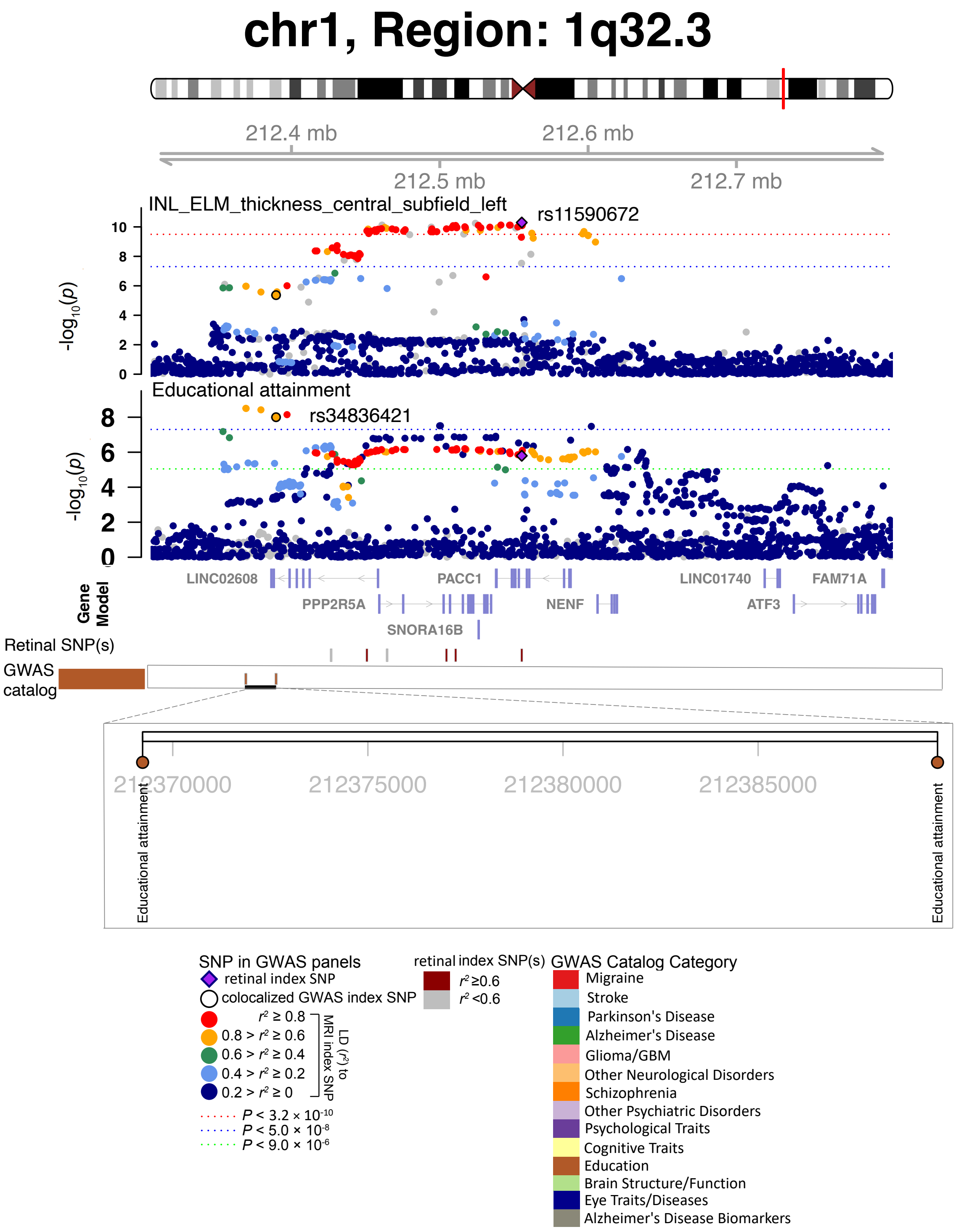


**Fig. S63 Selected genetic loci associated with retinal imaging traits and brain-related complex traits and diseases.** In 1p13.3, we observed the shared association (LD $r^2 \geq 0.6$) between the retinal imaging trait (INL_ELM_thickness_central_subfield_left, index variant rs11590672) and educational attainment (index variant rs34836421).

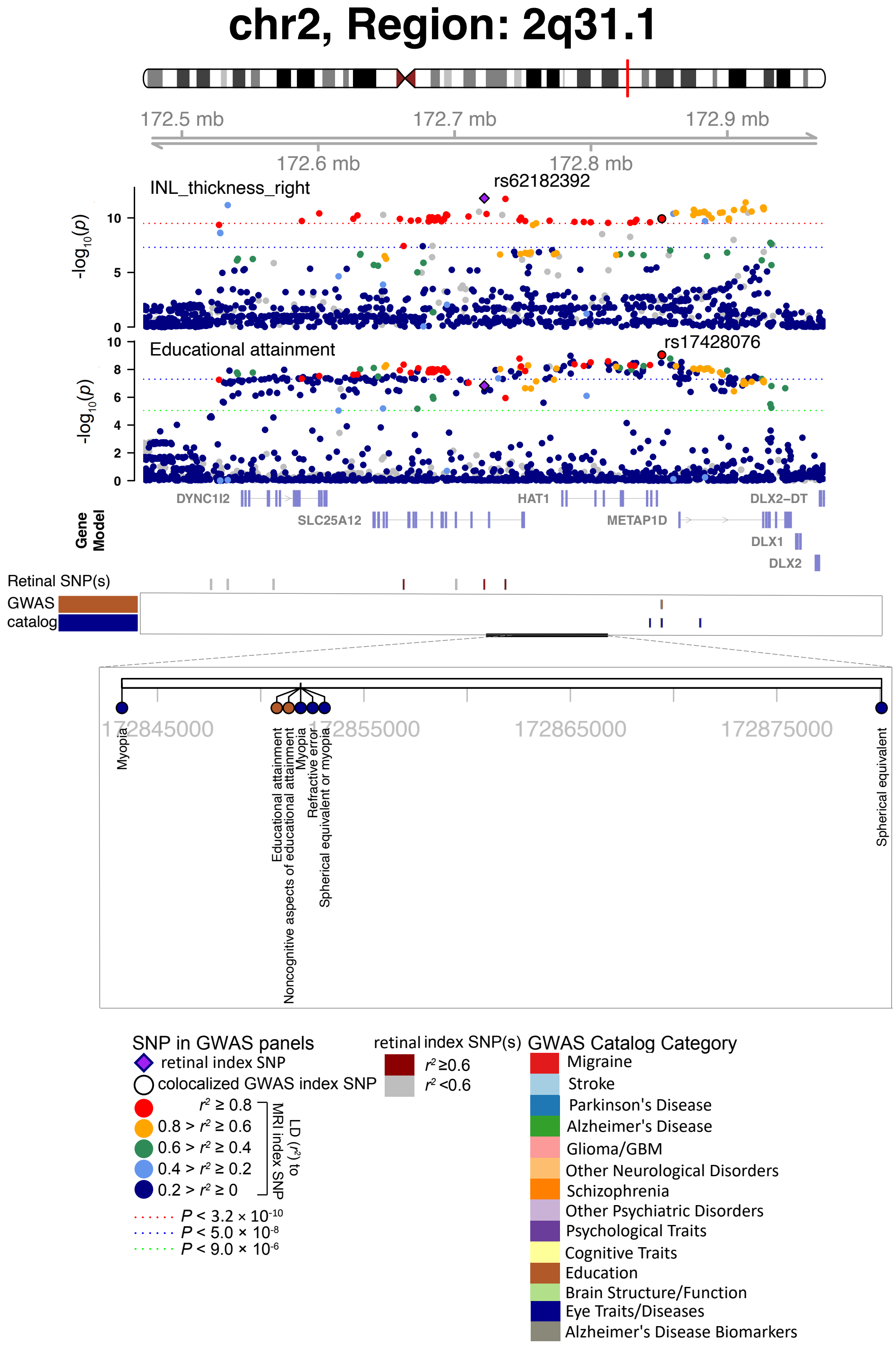


**Fig. S64 Selected genetic loci associated with retinal imaging traits and brain-related complex traits and diseases.** In 2q31.1, we observed the shared association (LD $r^2 \geq 0.6$) between the retinal imaging trait (INL_thickness_right, index variant rs62182392) and educational attainment (index variant rs17428076).

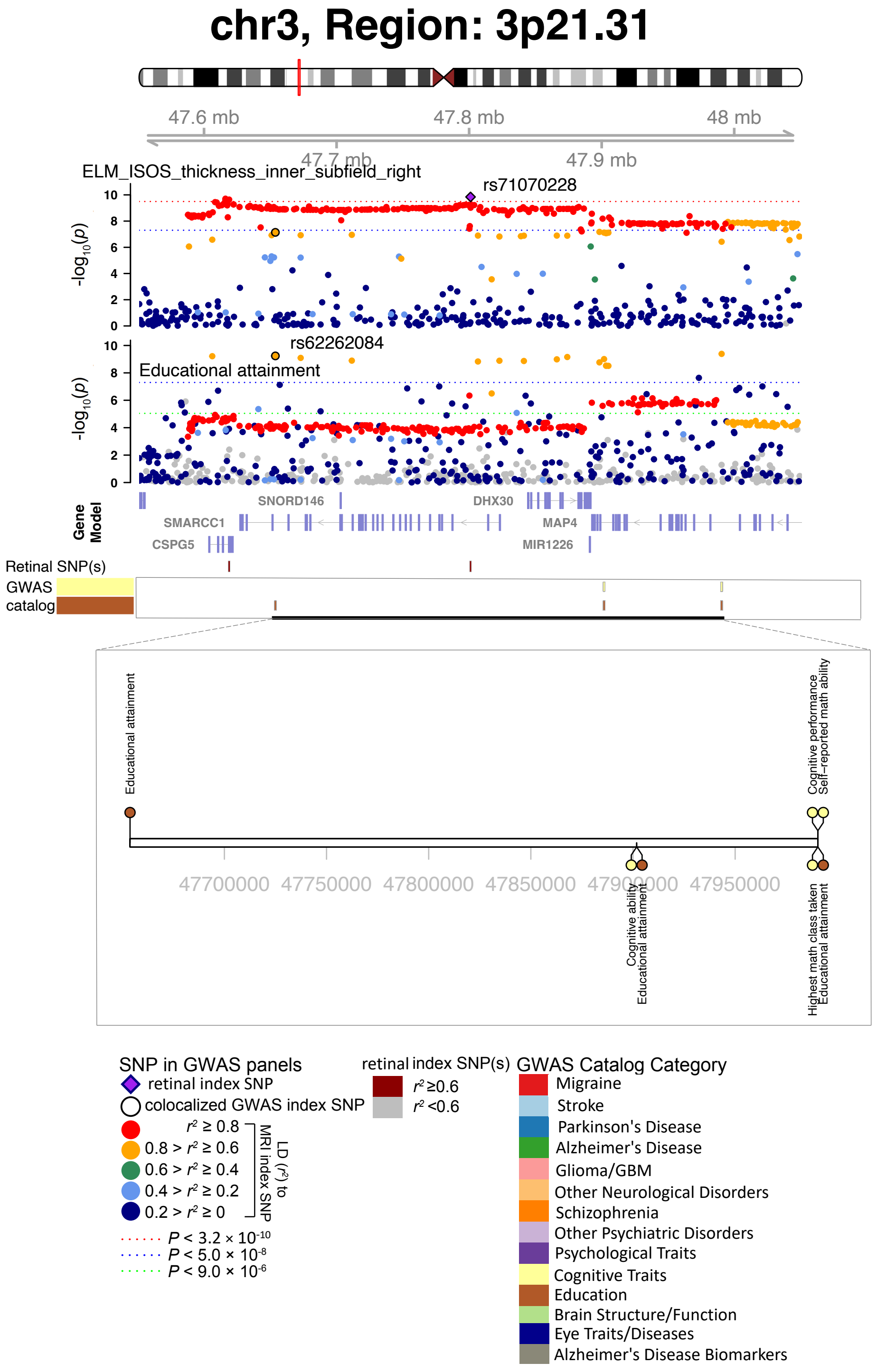


**Fig. S65 Selected genetic loci associated with retinal imaging traits and brain-related complex traits and diseases.** In 3p21.31, we observed the shared association (LD $r^2 \geq 0.6$) between the retinal imaging trait (ELM_ISOS_thickness_inner_subfield_right, index variant rs71070228) and educational attainment (index variant rs62262084).

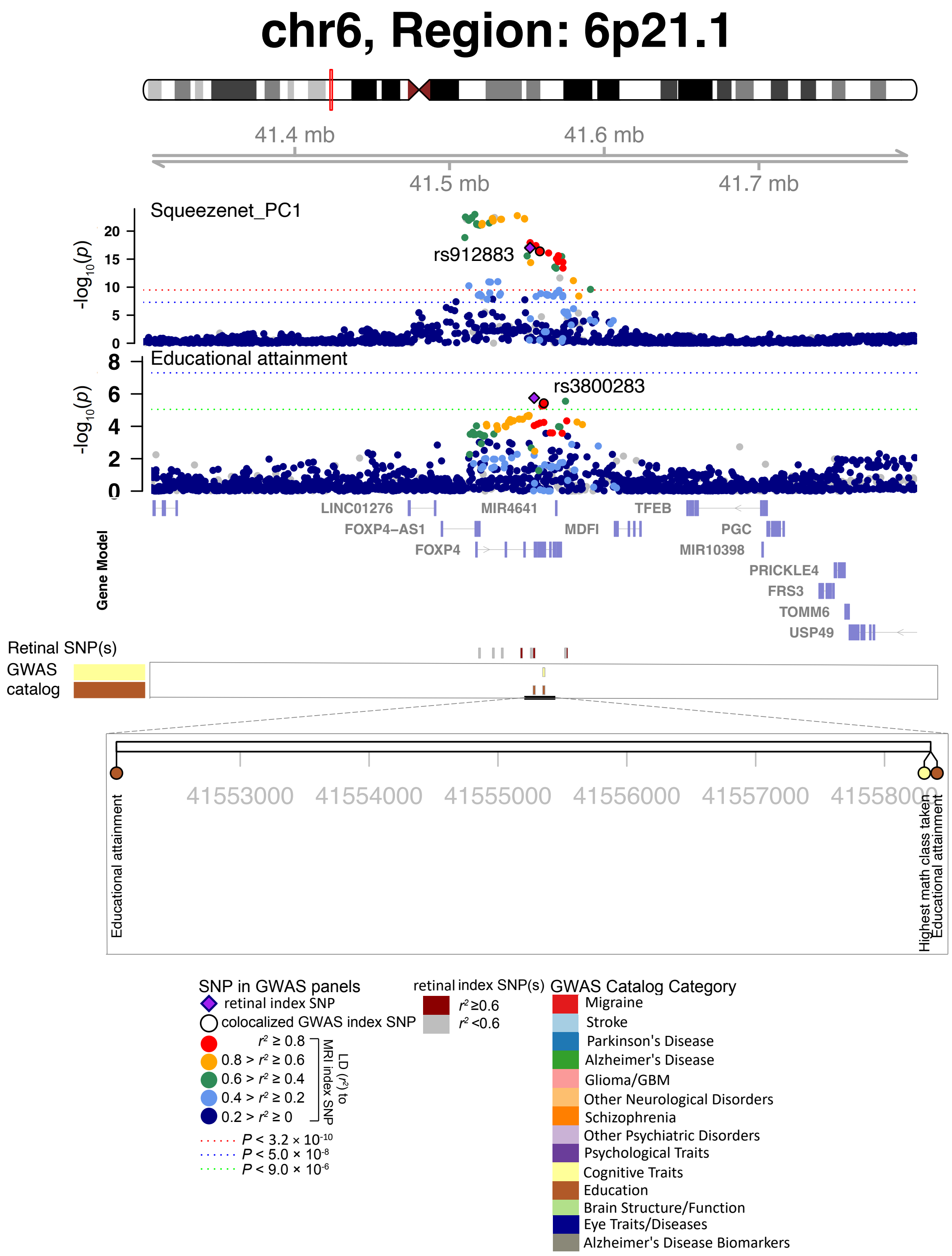


**Fig. S66 Selected genetic loci associated with retinal imaging traits and brain-related complex traits and diseases.** In 6p21.1, we observed the shared association (LD $r^2 \geq 0.6$) between the retinal imaging trait (Squeezenet_PC1, index variant rs912883) and educational attainment (index variant rs3800283).

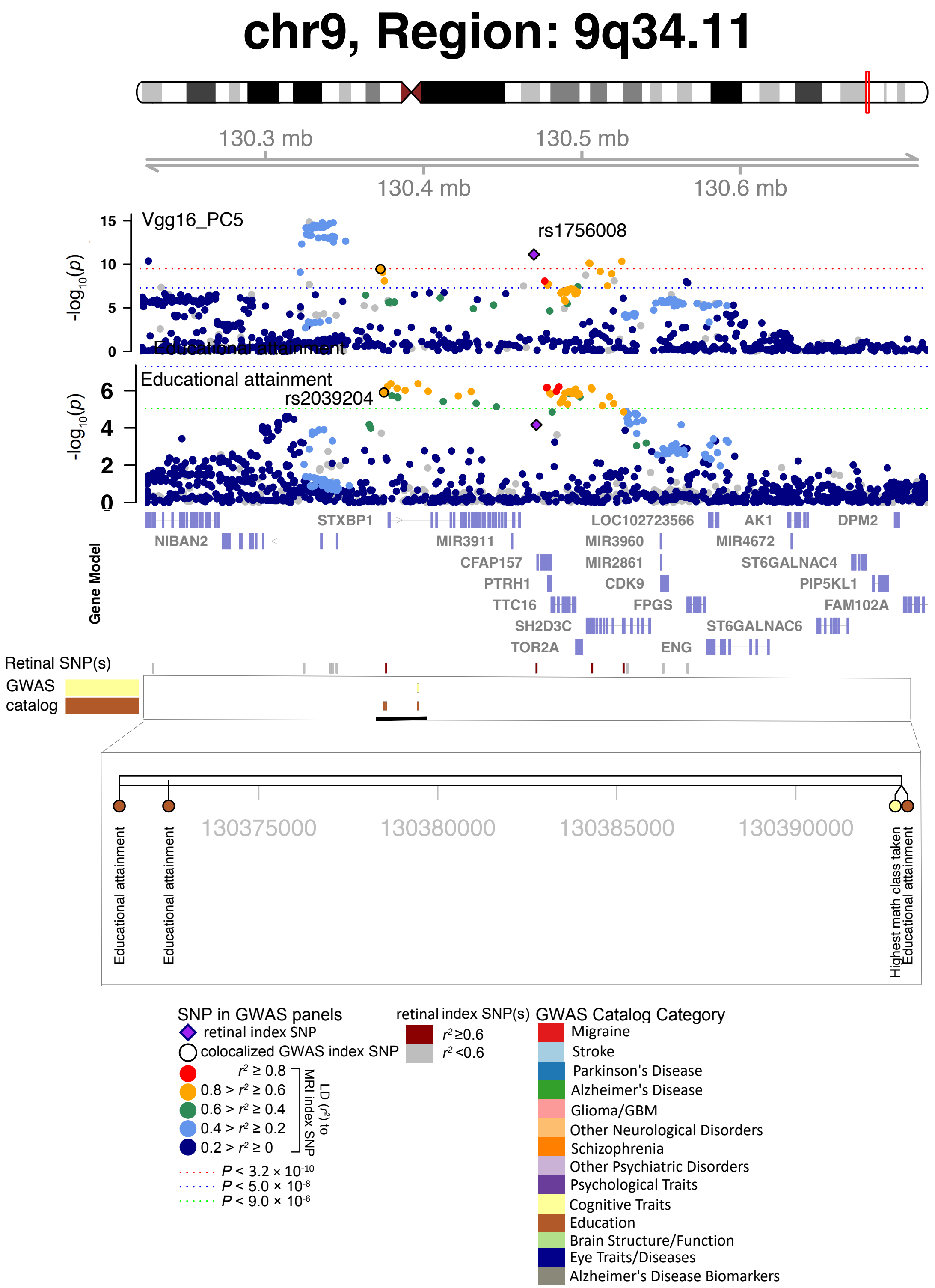


**Fig. S67 Selected genetic loci associated with retinal imaging traits and brain-related complex traits and diseases.** In 9q34.11, we observed the shared association (LD $r^2 \geq 0.6$) between the retinal imaging trait (Vgg16_PC5, index variant rs1756008) and educational attainment (index variant rs2039204).

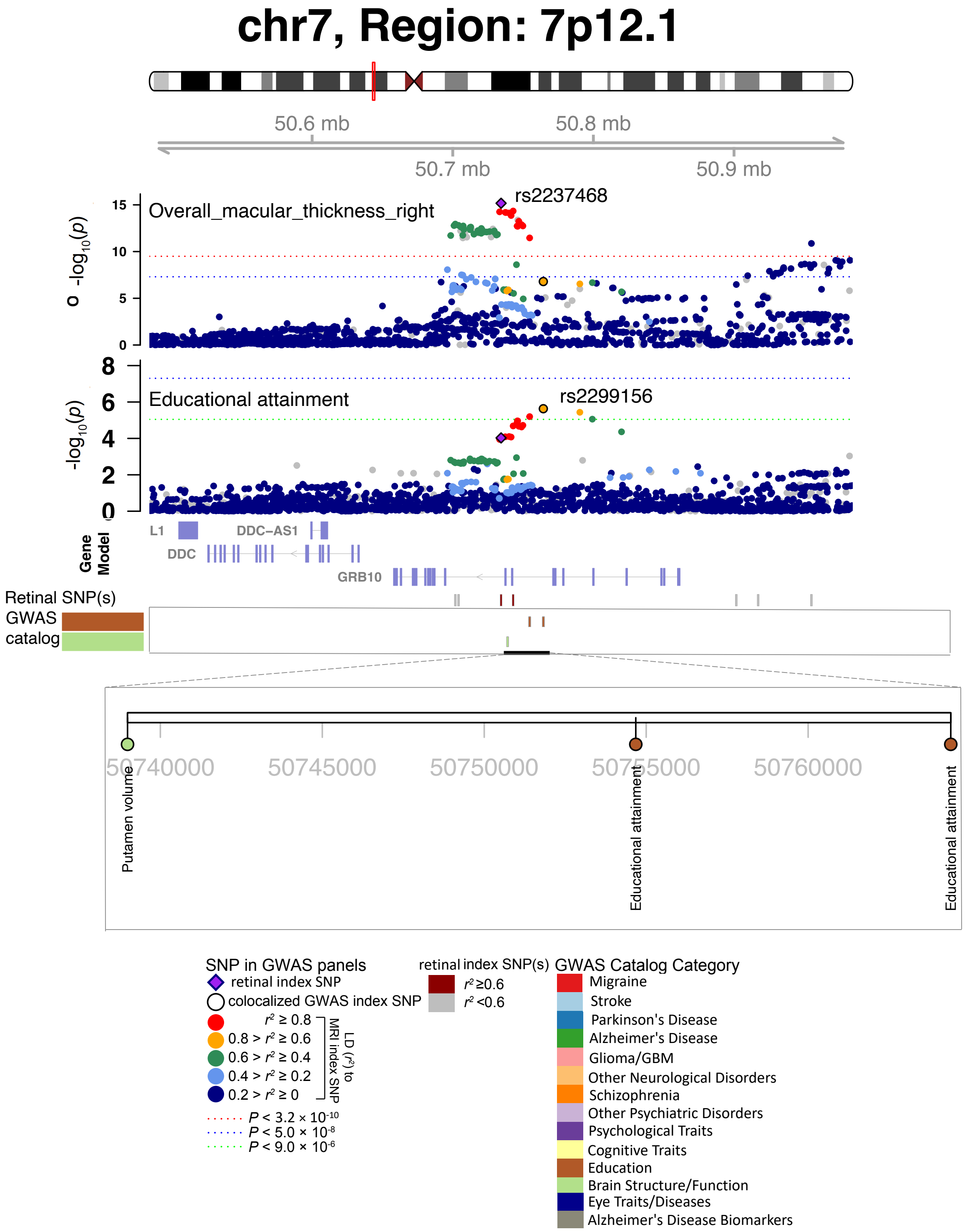


**Fig. S68 Selected genetic loci associated with retinal imaging traits and brain-related complex traits and diseases.** In 7p12.1, we observed the shared association (LD $r^2 \geq 0.6$) between the retinal imaging trait (overall_macular_thickness_right, index variant rs2237468) and educational attainment (index variant rs2299156).

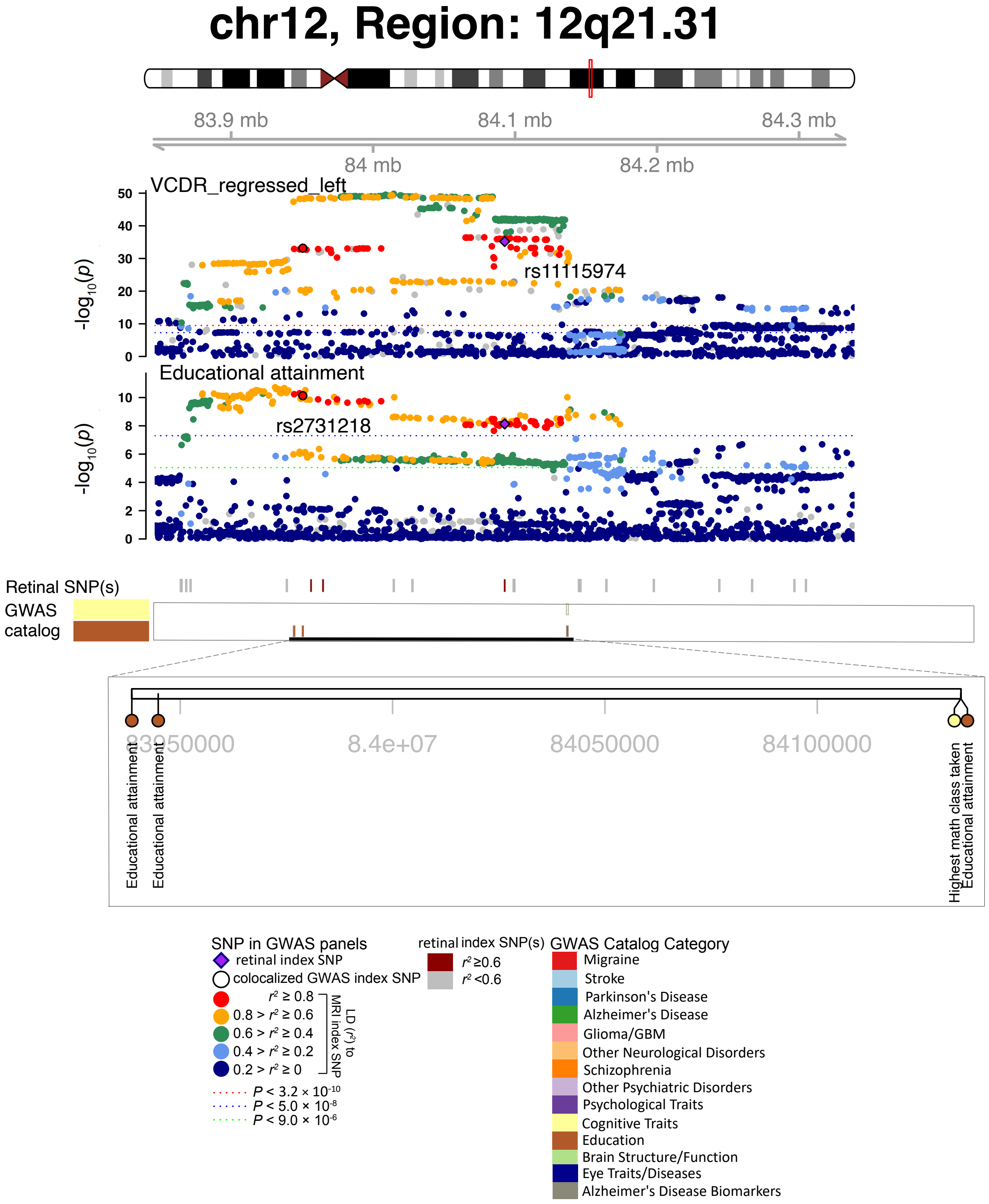


**Fig. S69 Selected genetic loci associated with retinal imaging traits and brain-related complex traits and diseases.** In 12q21.31, we observed the shared association (LD $r^2 \geq 0.6$) between the retinal imaging trait (VCDR_regressed_left, index variant rs11115974) and educational attainment (index variant rs2731218).

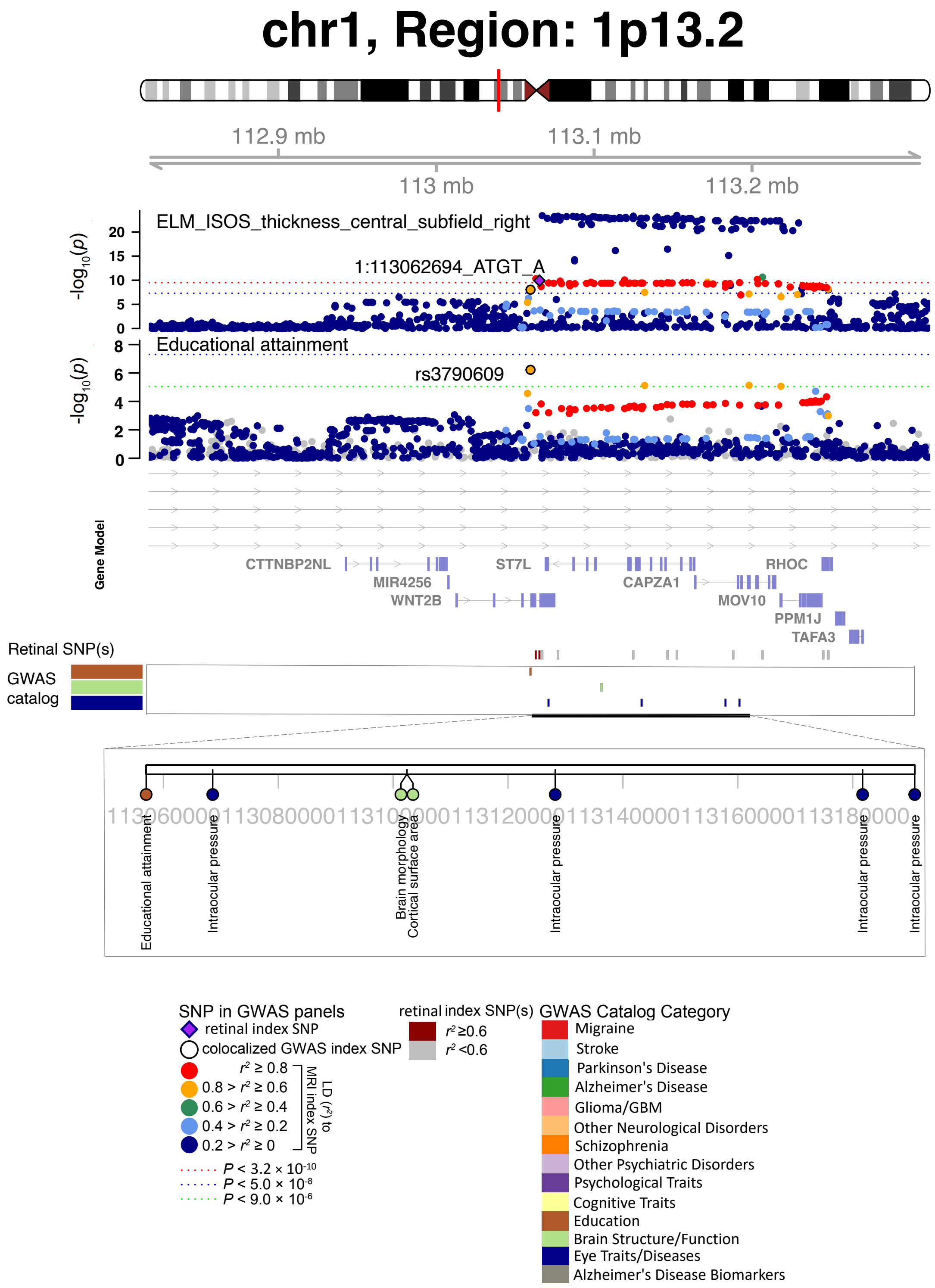


**Fig. S70 Selected genetic loci associated with retinal imaging traits and brain-related complex traits and diseases.** In 1p13.2, we observed the shared association (LD $r^2 \geq 0.6$) between the retinal imaging trait (ELM_ISOS_thickness_central_subfield_right, index variant 1:113062694_ATGT_A) and educational attainment (index variant rs3790609).

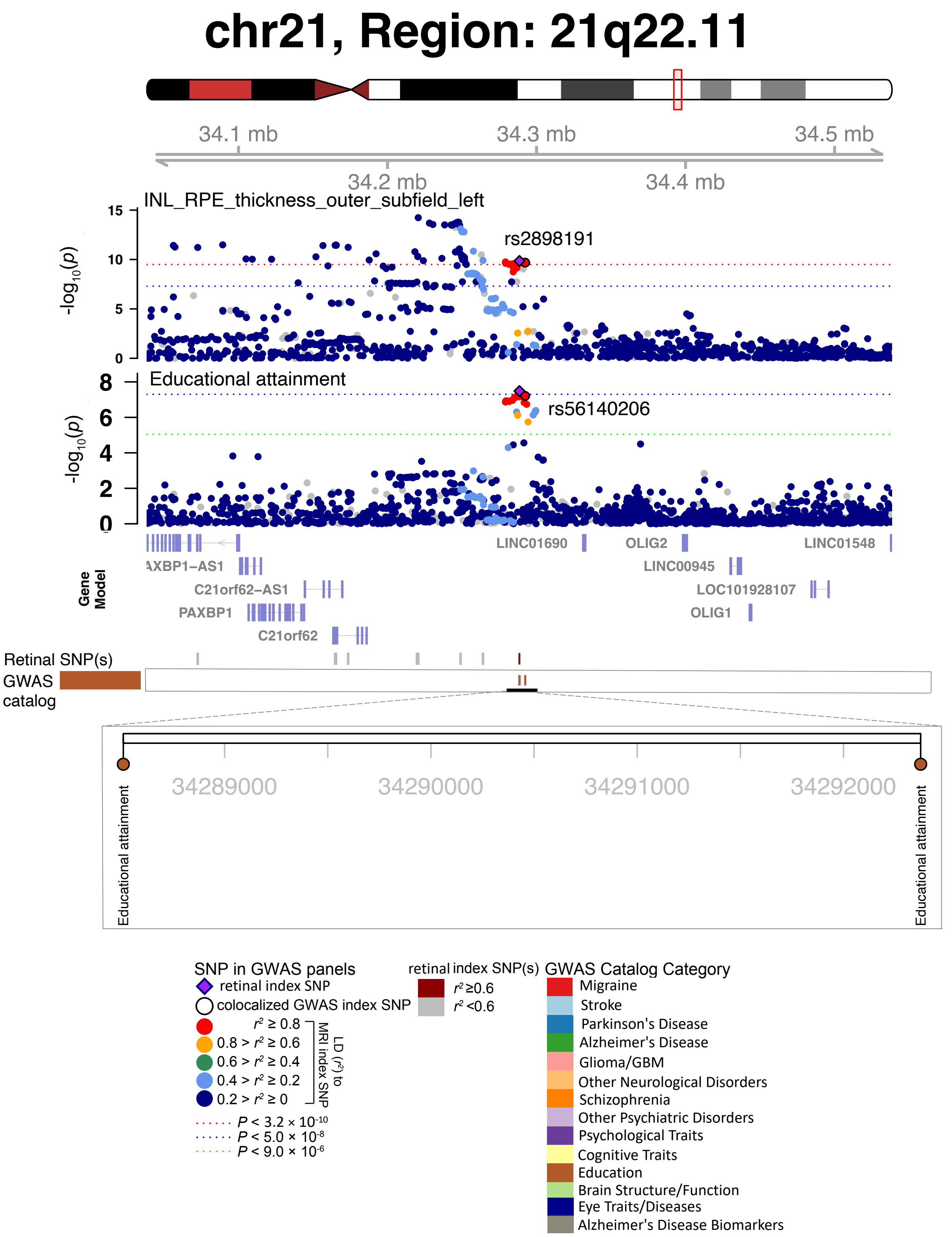


**Fig. S71 Selected genetic loci associated with retinal imaging traits and brain-related complex traits and diseases.** In 21q22.11, we observed the shared association (LD $r^2 \geq 0.6$) between the retinal imaging trait (INL_RPE_thickness_outer_subfield_left, index variant rs2898191) and educational attainment (index variant rs56140206).

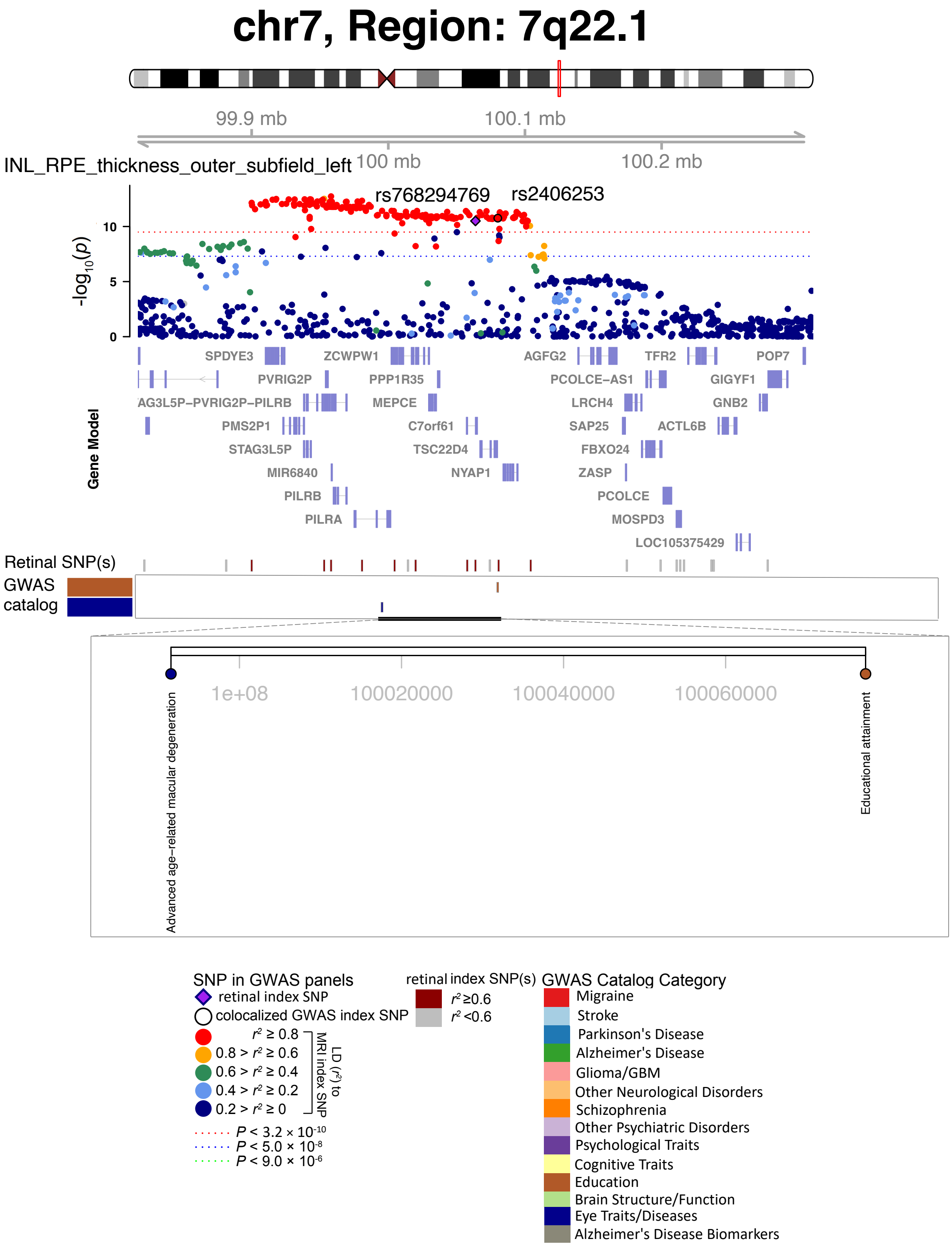


**Fig. S72 Selected genetic loci associated with retinal imaging traits and brain-related complex traits and diseases.** In 7q22.1, we observed the shared association (LD $r^2 \geq 0.6$) between the retinal imaging trait (INL_RPE_thickness_outer_subfield_left, index variant rs768294769) and educational attainment (index variant rs2406253).

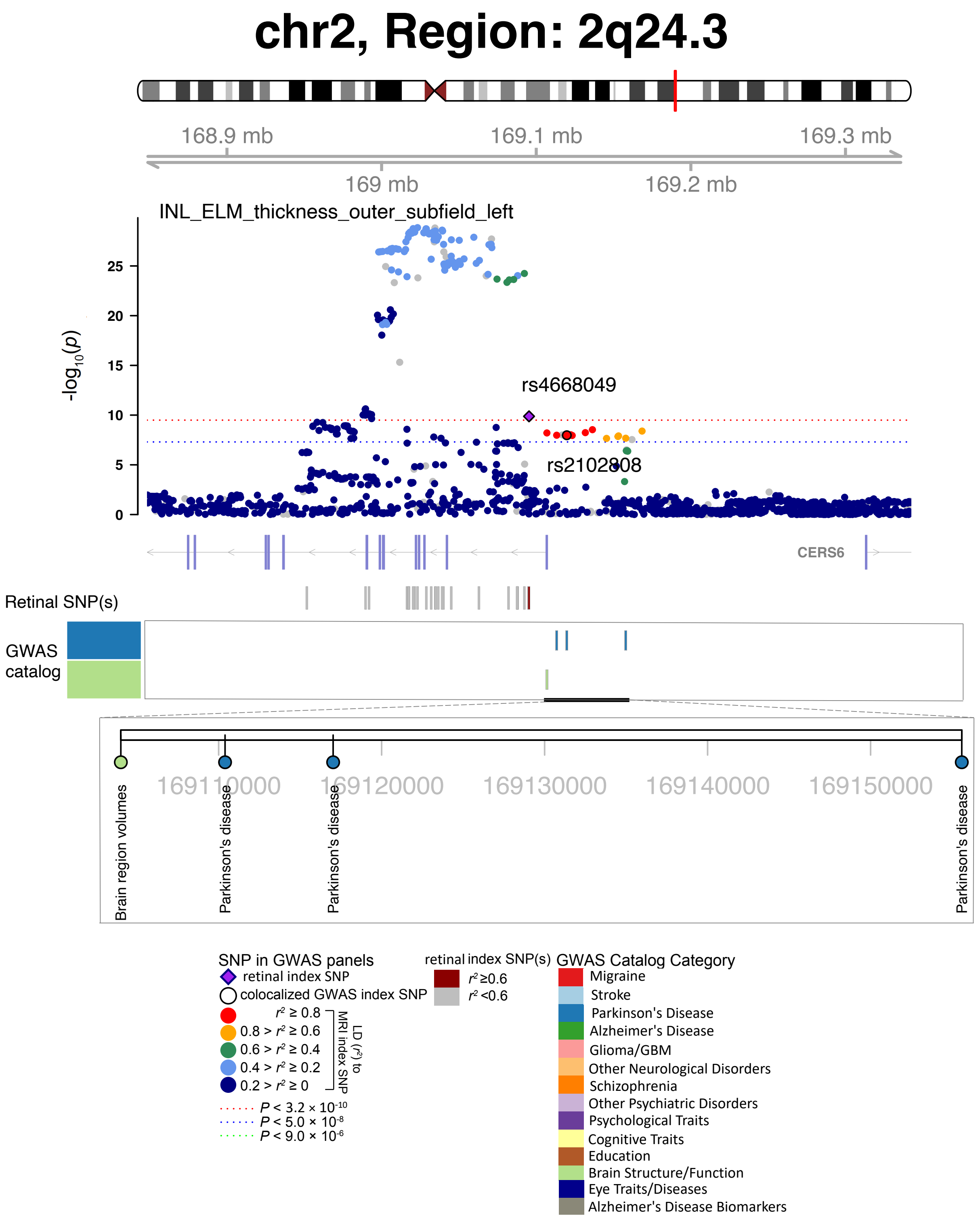


**Fig. S73 Selected genetic loci associated with retinal imaging traits and brain-related complex traits and diseases.** In 2q24.3, we observed the shared association (LD $r^2 \geq 0.6$) between the retinal imaging trait (INL_ELM_thickness_outer_subfield_left, index variant rs4668049) and Parkinson's disease (index variant rs2102808).

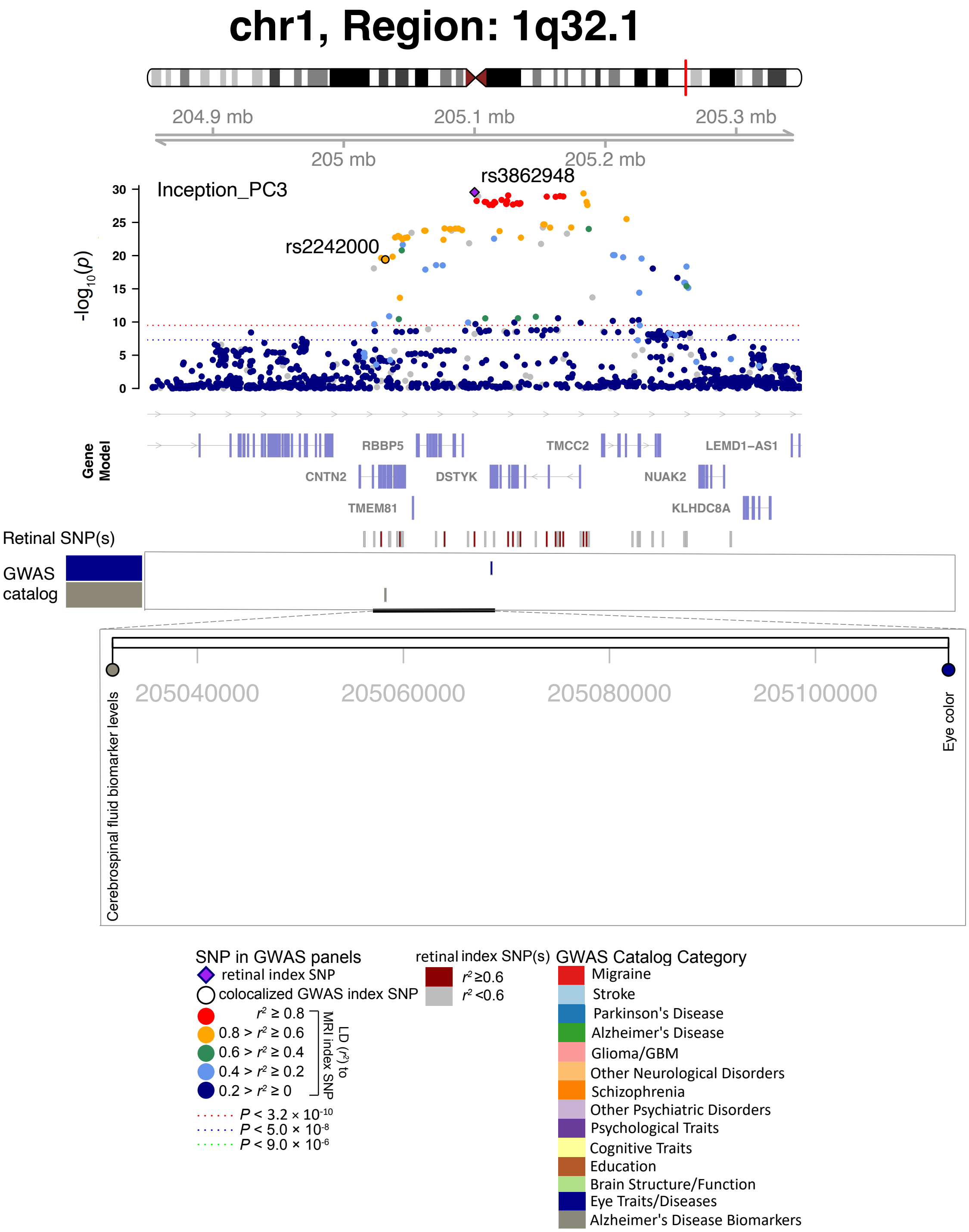


**Fig. S74 Selected genetic loci associated with retinal imaging traits and brain-related complex traits and diseases.** In 1q32.1, we observed the shared association (LD $r^2 \geq 0.6$) between the retinal imaging trait (Inception_PC3, index variant rs3862948) and cerebrospinal fluid biomarker levels (index variant rs2242000).

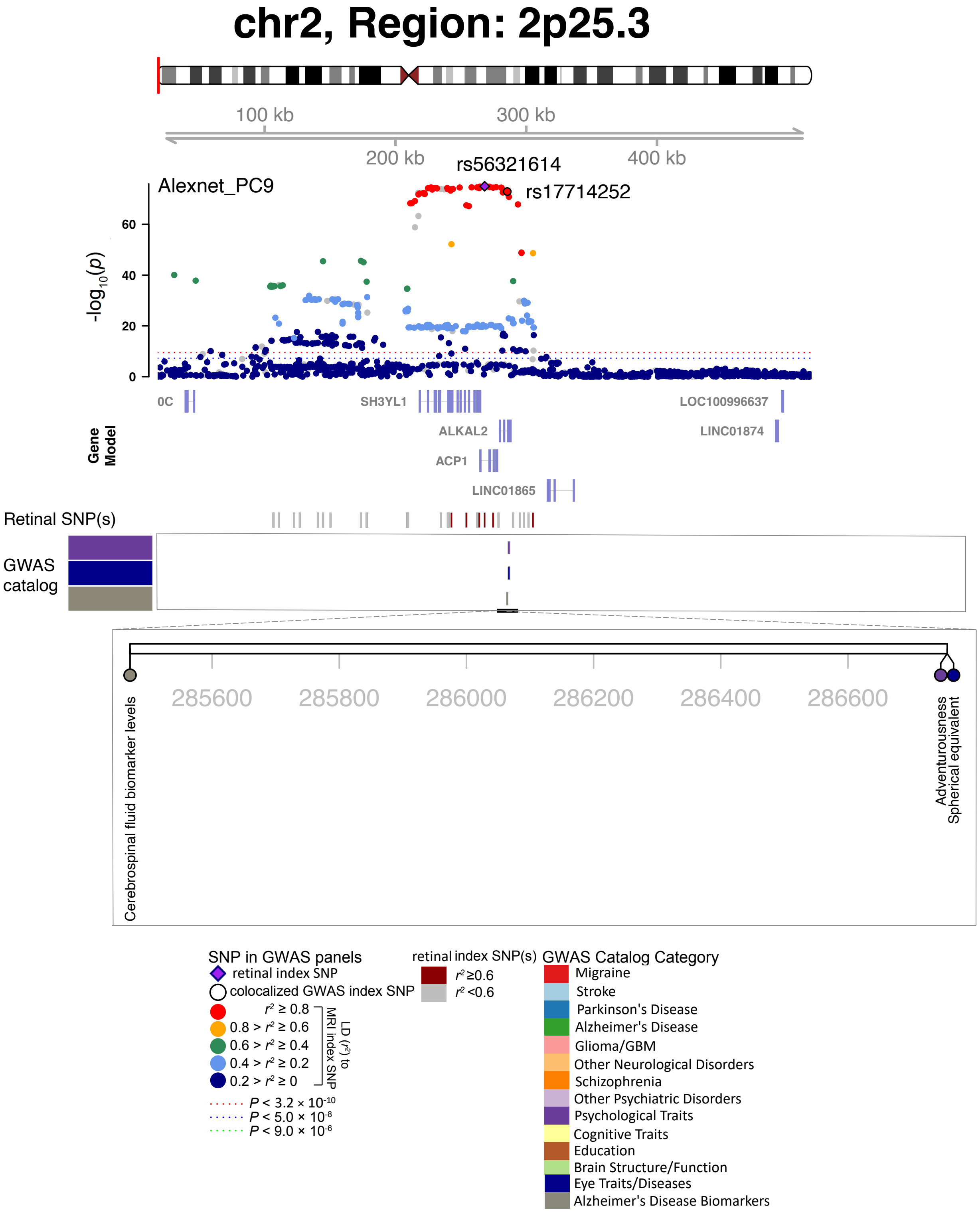


**Fig. S75 Selected genetic loci associated with retinal imaging traits and brain-related complex traits and diseases.** In 2p25.3, we observed the shared association (LD $r^2 \geq 0.6$) between the retinal imaging trait (Alexnet_PC9, index variant rs56321614) and cerebrospinal fluid biomarker levels (index variant rs17714252).

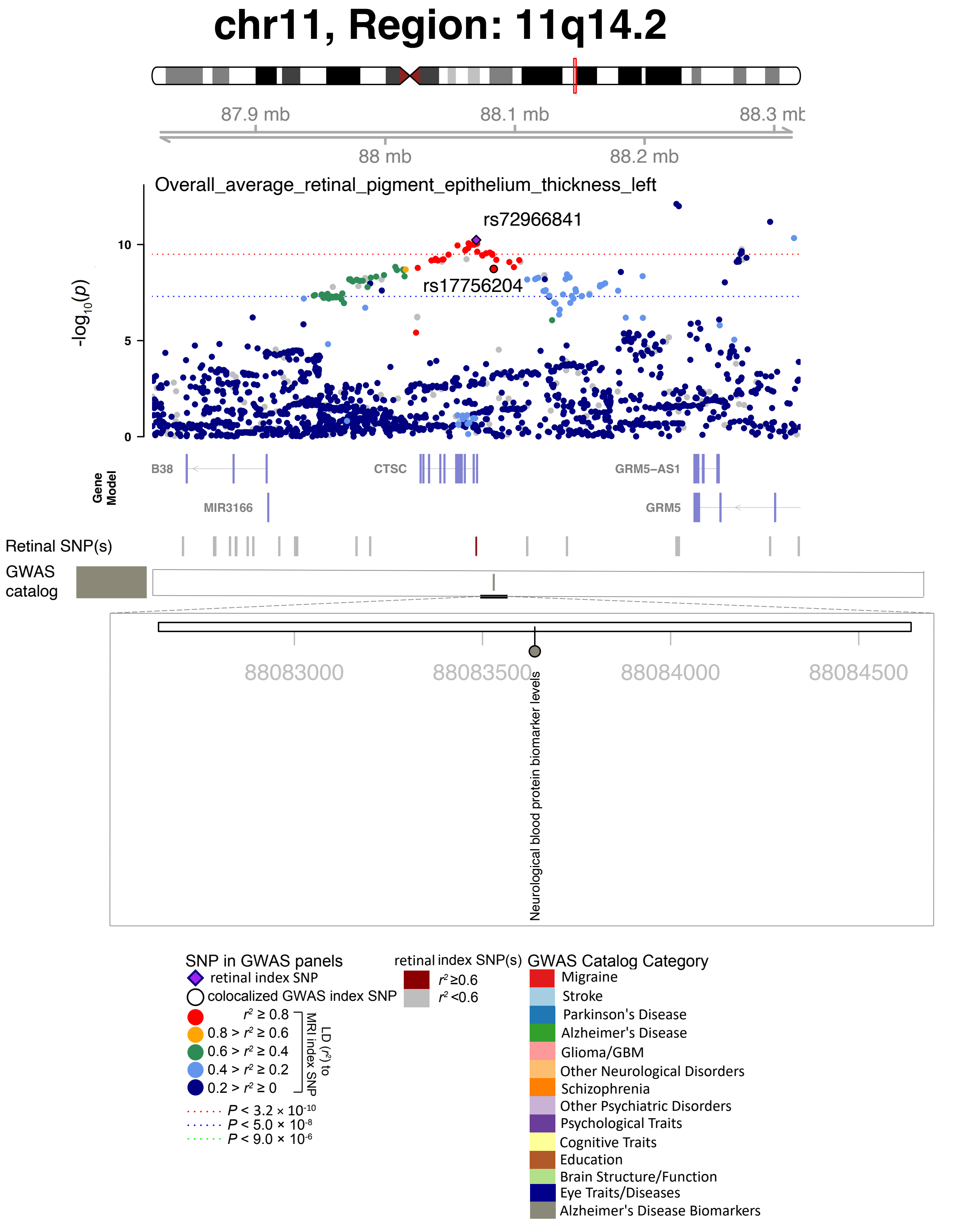


**Fig. S76 Selected genetic loci associated with retinal imaging traits and brain-related complex traits and diseases.** In 11q14.2, we observed the shared association (LD $r^2 \geq 0.6$) between the retinal imaging trait (overall_average_retinal_pigment_epithelium_thickness_left, index variant rs72966841) and neurological blood protein biomarker levels (index variant rs17756204).

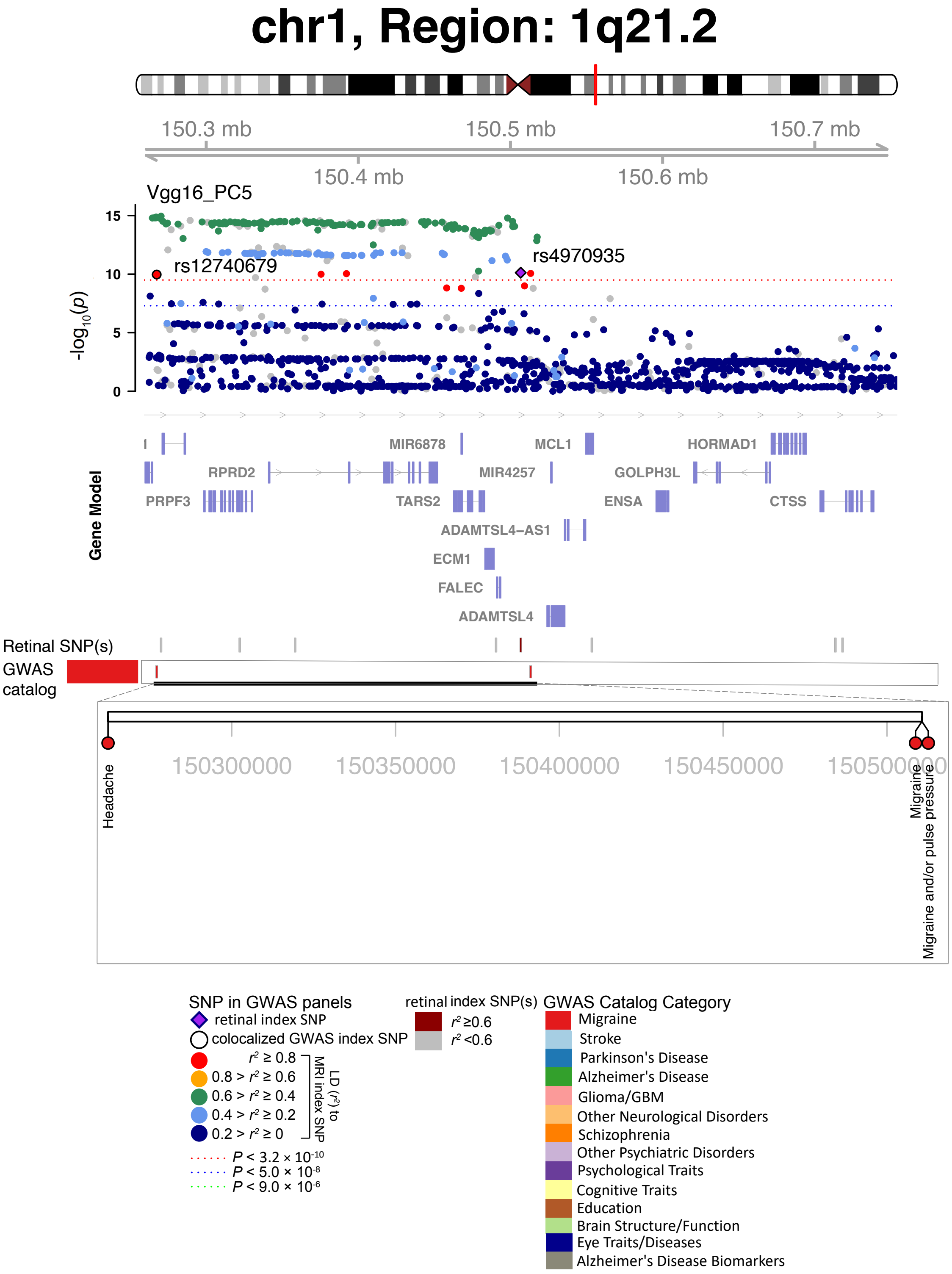


**Fig. S77 Selected genetic loci associated with retinal imaging traits and brain-related complex traits and diseases.** In 1q21.2, we observed the shared association (LD $r^2 \geq 0.6$) between the retinal imaging trait (Vgg16_PC5, index variant rs4970935) and headache (index variant rs12740679).

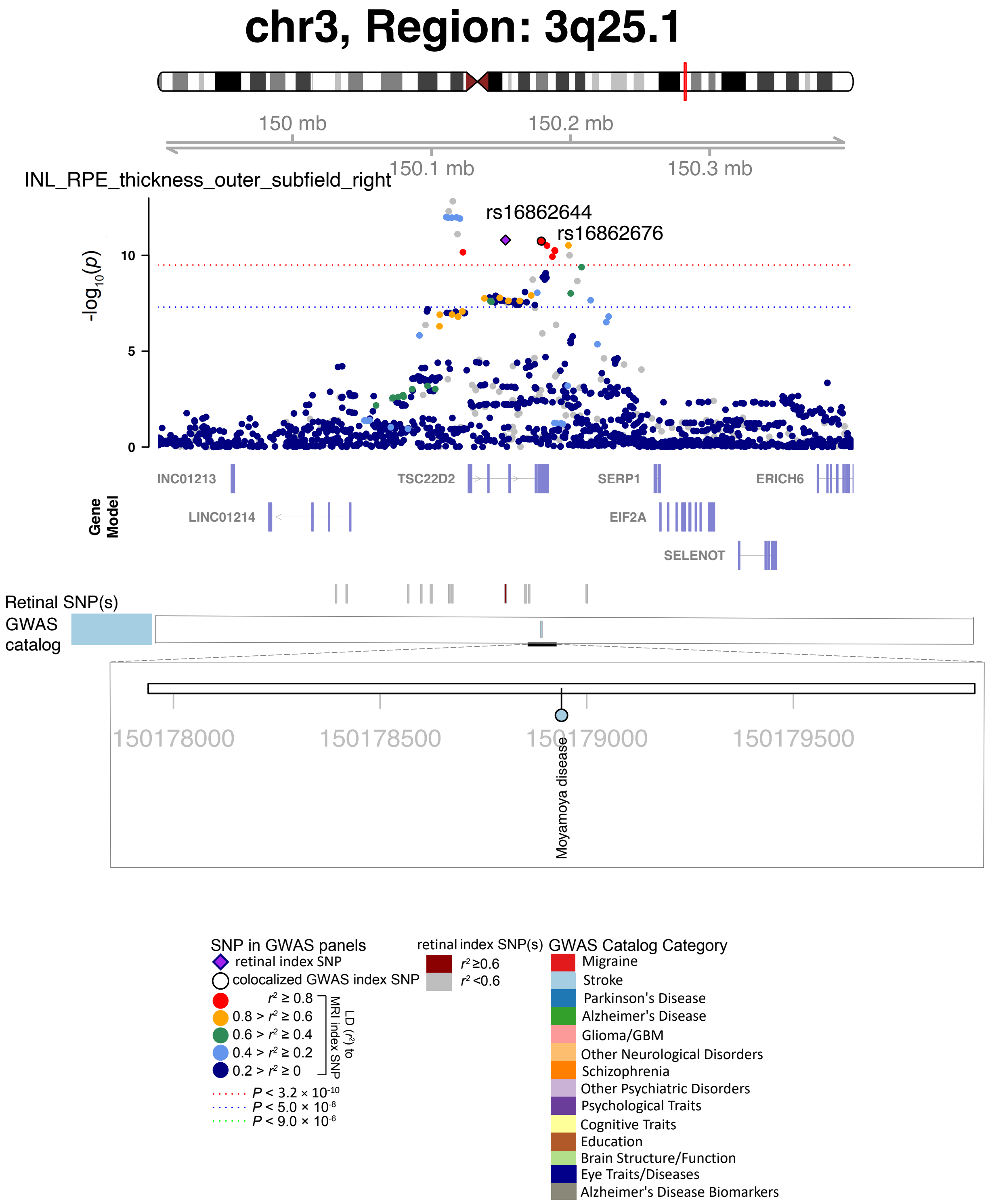


**Fig. S78 Selected genetic loci associated with retinal imaging traits and brain-related complex traits and diseases.** In 3q25.1, we observed the shared association (LD $r^2 \geq 0.6$) between the retinal imaging trait (INL_RPE_thickness_outer_subfield_right, index variant rs16862644) and Moyamoya disease (index variant rs16862676).

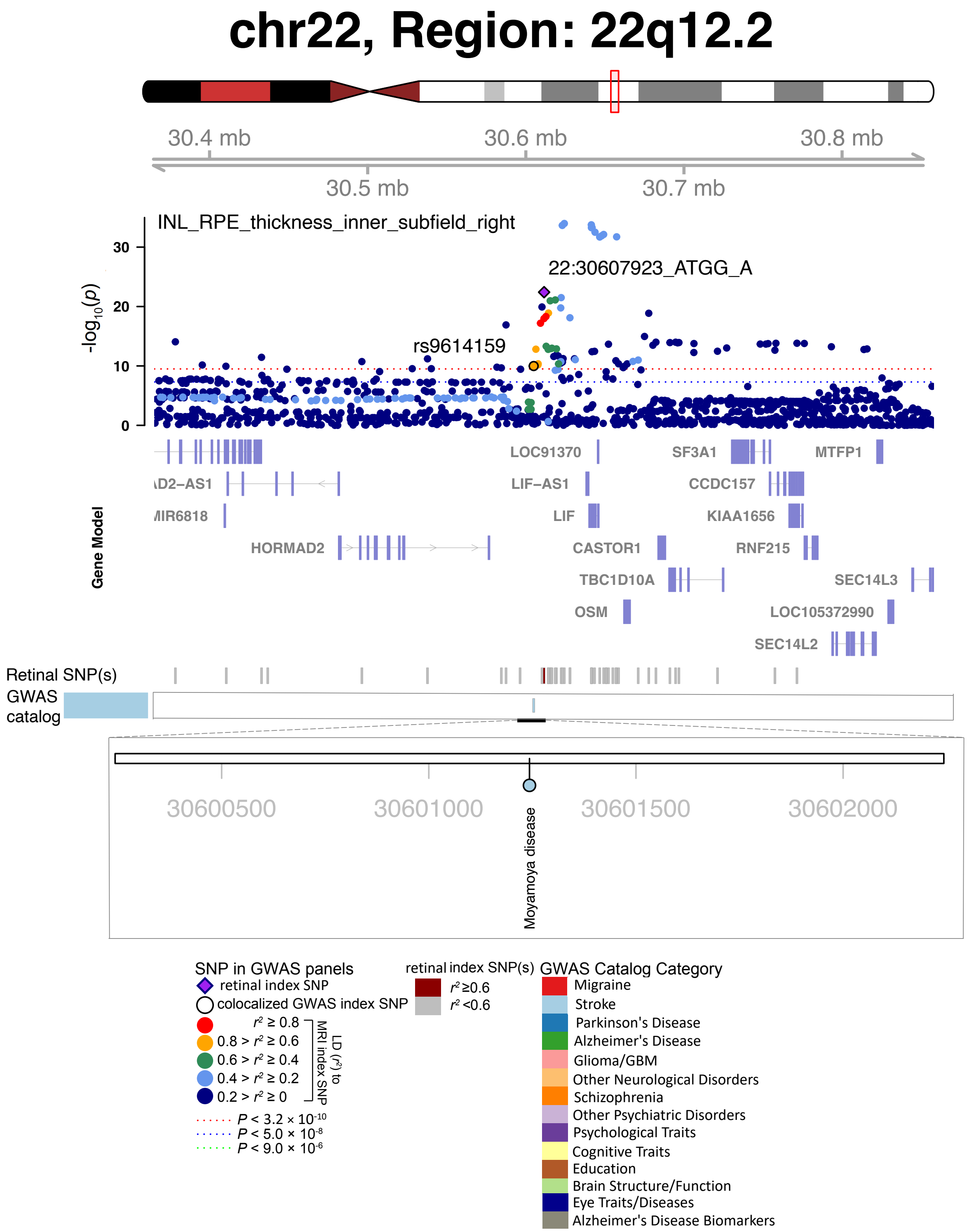


**Fig. S79 Selected genetic loci associated with retinal imaging traits and brain-related complex traits and diseases.** In 22q12.2, we observed the shared association (LD $r^2 \geq 0.6$) between the retinal imaging trait (INL_RPE_thickness_inner_subfield_right, index variant 22:30607923_ATGG_A) and Moyamoya disease (index variant rs9614159).

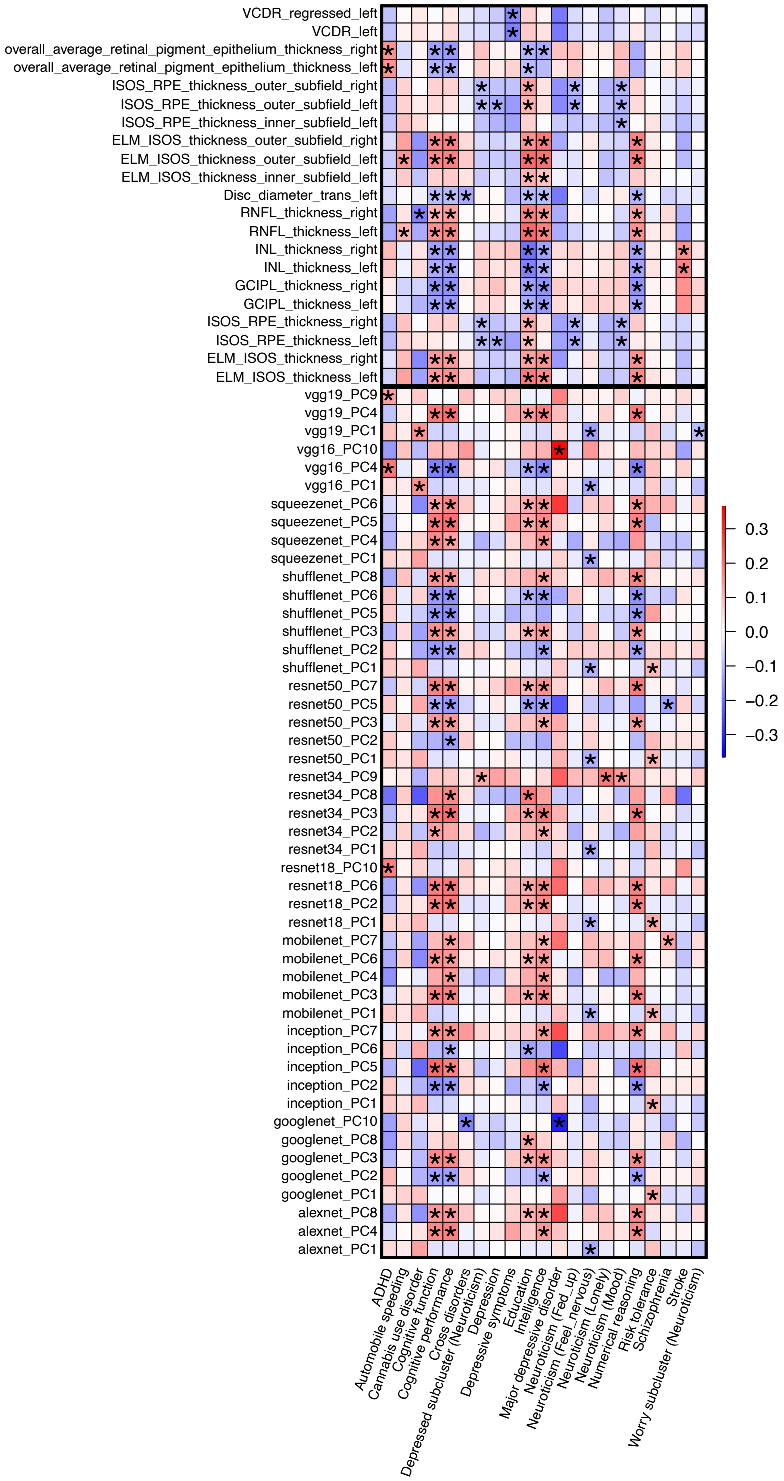


**Fig. S80 Pairwise genetic correlations between retinal imaging traits and brain-related complex traits and diseases.** The asterisks highlight significant associations after controlling the false discovery rate at 5% level. The color represents genetic correlation estimates. The *x*-axis lists the ID of brain-related complex traits and diseases analyzed in previous GWAS, and the *y*-axis provides the name of our retinal imaging traits.

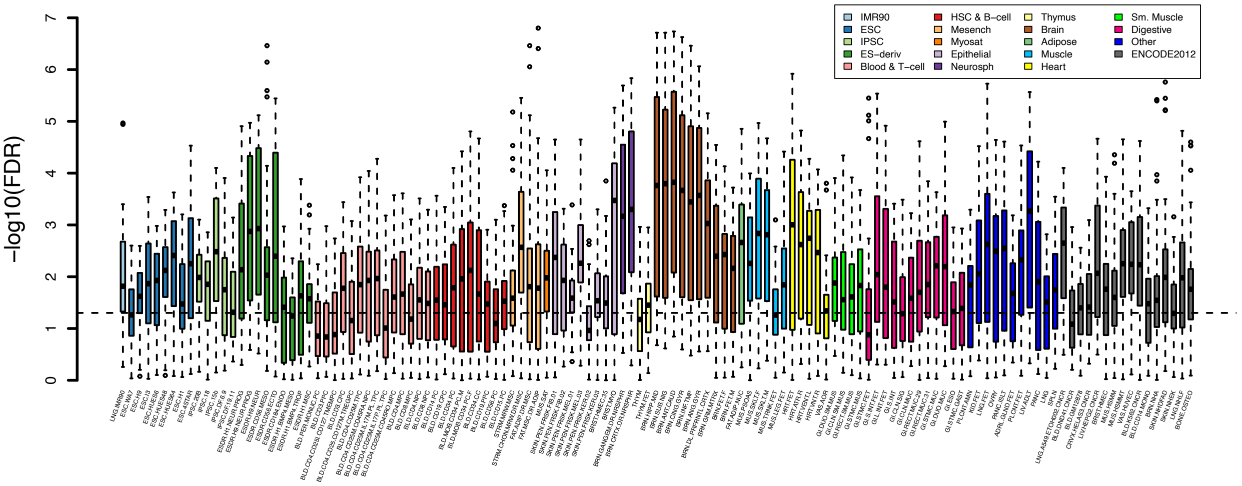


**Fig. S81 Partitioned heritability enrichment analysis of retinal imaging traits for tissue type and cell type-specific regulatory elements.** The dashed lines indicate the significance level after controlling the false discovery rate at 5% level. Different colors represent different cell and tissue groups from the Roadmap Epigenomics Project. We label the names of each regulatory element on the *x*-axis.

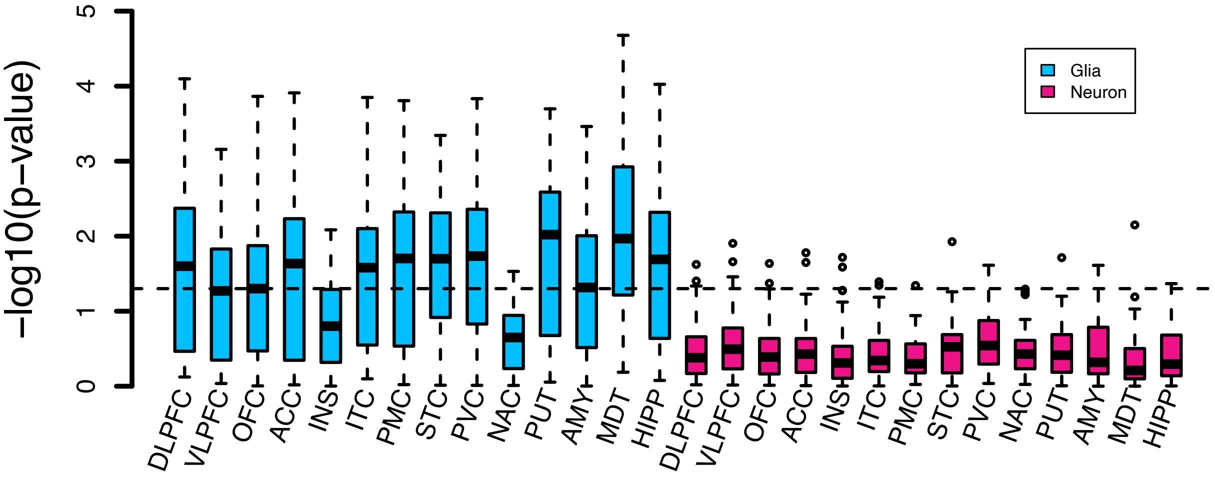


**Fig. S82 Partitioned heritability enrichment analysis of retinal imaging traits in brain cell types.** We show the heritability enrichment of retinal imaging traits in regulatory elements of two brain cell types (neuron and glia) sampled from 14 brain cortical and subcortical regions. DLFPC, dorsolateral prefrontal cortex; VLPFC, ventrolateral prefrontal cortex; OFC, orbitofrontal cortex; ACC, anterior cingulate cortex; INS, insular cortex; ITC, inferior temporal cortex; STC, superior temporal cortex; PMC, primary motor cortex; PVC, primary visual cortex; AMY, amygdala; HIPP, hippocampus; MDT, mediodorsal thalamus; NAC, nucleus accumbens; PUT, putamen. The dashed lines indicate the nominal (0.05) significance level.

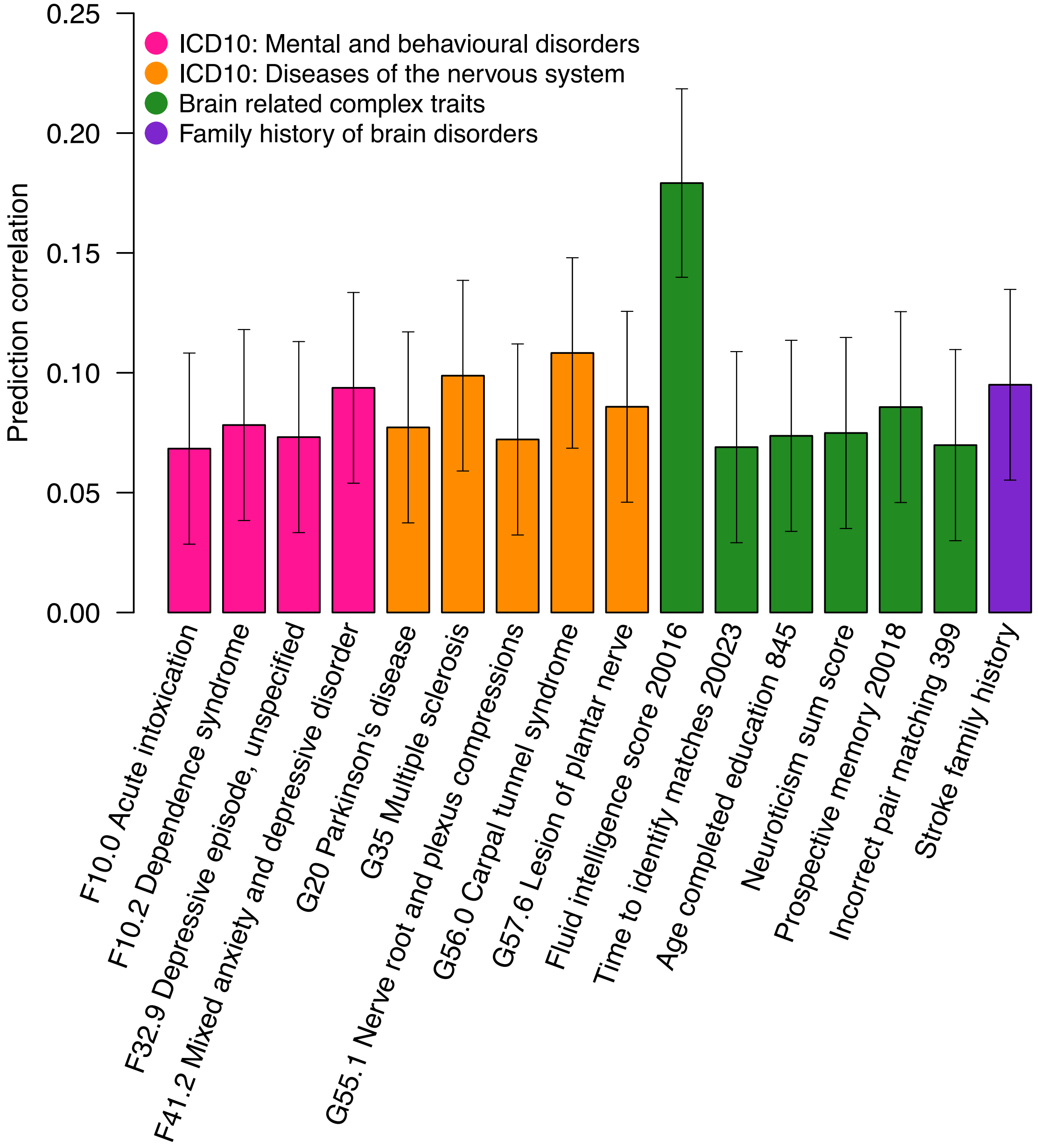


**Fig. S83 Prediction performance of retinal imaging traits for brain-related complex traits and diseases.** Different colors represent categories of complex traits and diseases.

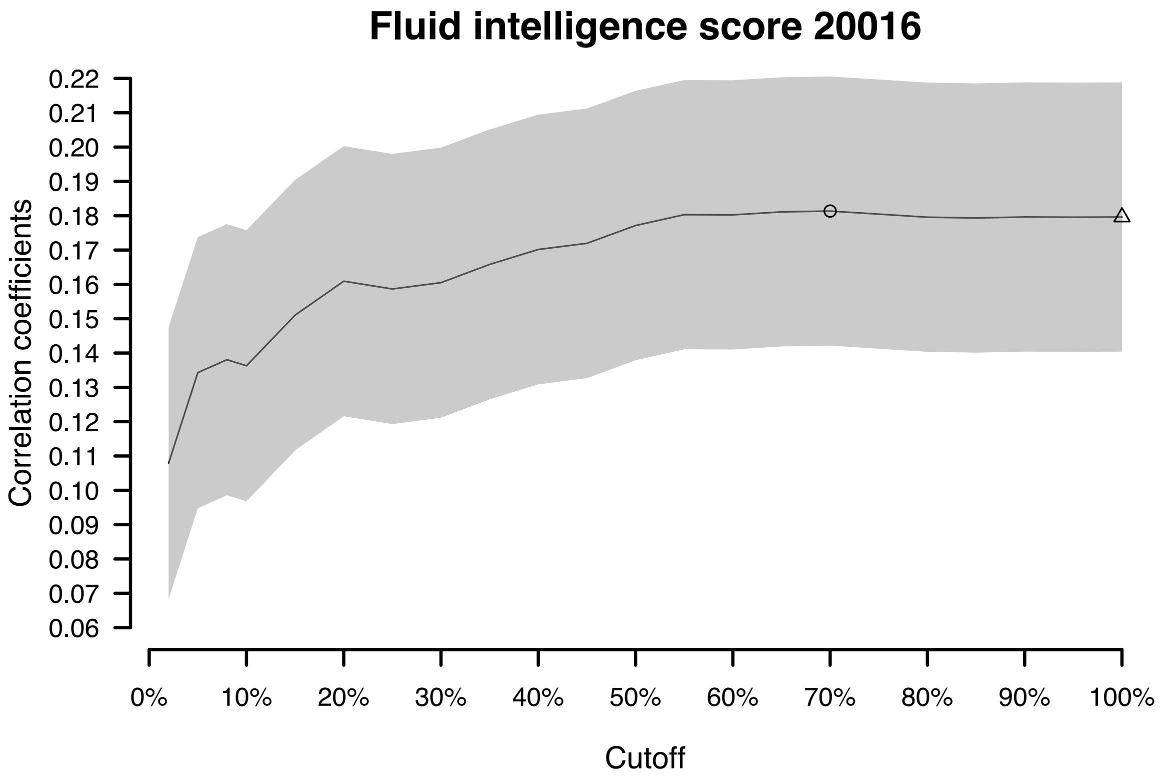


**Fig. S84 Prediction performance of retinal imaging traits for fluid intelligence.** Prediction accuracy of retinal imaging traits on fluid intelligence score (Data field ID 20016) across different percentage thresholds. The prediction accuracy is measured by the correlation between observed and predicted values in the testing data. On the *x*-axis, there are a series of percentage thresholds, suggesting that the top *x* percent of retinal imaging traits were included in the model as predictors. From left to right, there are more predictors included in the model. The circle indicates the threshold with the best prediction accuracy, and the triangle indicates the performance of using all retinal imaging traits.

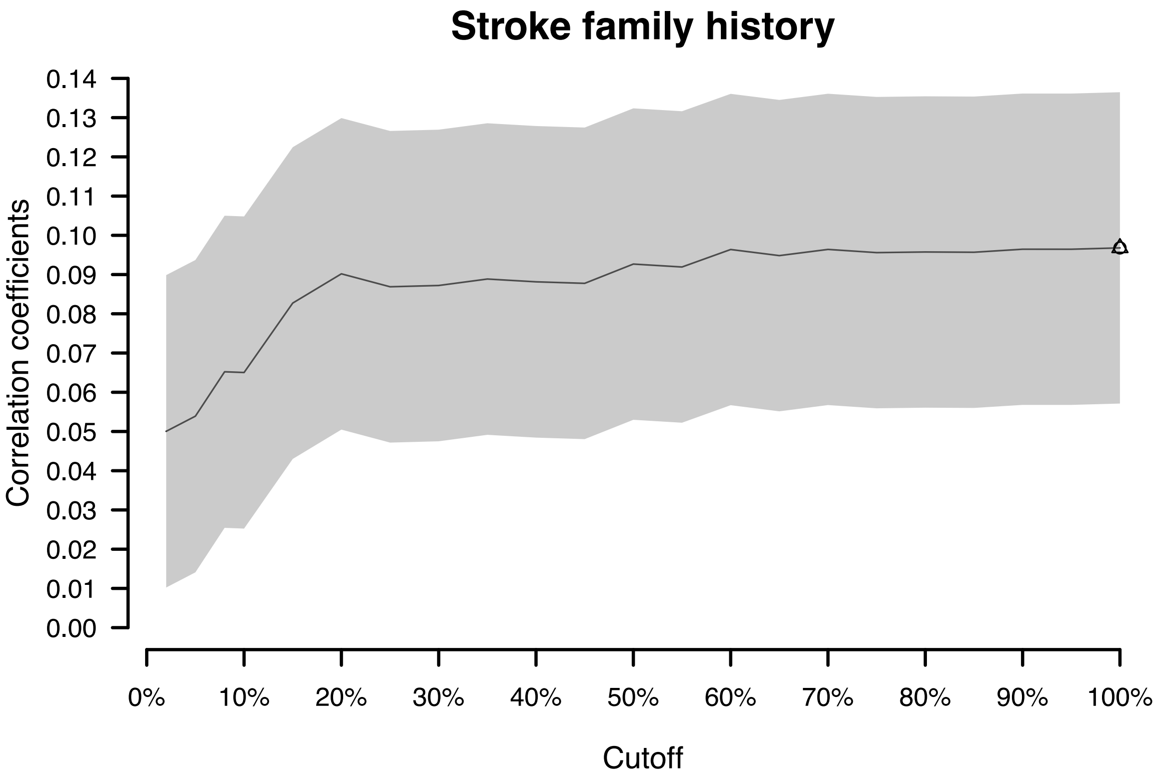


**Fig. S85 Prediction performance of retinal imaging traits for the family history of stroke.** Prediction accuracy of retinal imaging traits on the family history of stroke across different percentage thresholds. The prediction accuracy is measured by the correlation between observed and predicted values in the testing data. On the *x*-axis, there are a series of percentage thresholds, suggesting that the top *x* percent of retinal imaging traits were included in the model as predictors. From left to right, there are more predictors included in the model. The circle indicates the threshold with the best prediction accuracy, and the triangle indicates the performance of using all retinal imaging traits. In this example, the two points are in the same position.

**Legends for Tables S1 to S12** (All tables can be found in a zip file).

**Table S1. ID of brain and retinal imaging traits used in phenotypic association analysis.**

**Table S2. Significant eye-brain phenotypic associations between retinal imaging traits and brain MRI traits.**

**Table S3. SNP-based heritability of 156 retinal imaging traits.**

**Table S4. Independent (LD < 0.1) significant associations for retinal imaging traits.**

**Table S5. Validating significant retinal imaging associations in independent validation GWAS datasets.**

**Table S6. Performance of polygenic risk scores of the 156 retinal imaging traits.**

**Table S7. Genetic overlaps of eye-brain connections.**

**Table S8. Summary of the shared genetic effects between retinal imaging traits and brain phenotypes.**

**Table S9. Genetic correlations between retinal imaging traits and brain-related complex traits and diseases analyzed in previous studies.**

**Table S10. LDSC partitioned heritability enrichment analysis.**

**Table S11. Mendelian randomization analysis between retinal imaging traits and brain diseases.**

**Table S12. Predicting complex traits and diseases using retinal imaging traits.**
